# Development and validation of a prognostic model for COVID-19: a population-based cohort study in Iceland

**DOI:** 10.1101/2021.07.19.21260759

**Authors:** Elias Eythorsson, Valgerdur Bjarnadottir, Hrafnhildur Linnet Runolfsdottir, Dadi Helgason, Ragnar Freyr Ingvarsson, Helgi K Bjornsson, Lovisa Bjork Olafsdóttir, Solveig Bjarnadottir, Arnar Snaer Agustsson, Kristin Oskarsdottir, Hrafn Hliddal Thorvaldsson, Guðrun Kristjansdottir, Aron Hjalti Bjornsson, Arna R Emilsdottir, Brynja Armannsdottir, Olafur Guðlaugsson, Sif Hansdottir, Magnus Gottfredsson, Agnar Bjarnason, Martin I Sigurdsson, Olafur S Indridason, Runolfur Palsson

## Abstract

**Background:** The severity of SARS-CoV-2 infection varies from asymptomatic state to severe respiratory failure and the clinical course is difficult to predict. The aim of the study was to develop a prognostic model to predict the severity of COVID-19 at the time of diagnosis and determine risk factors for severe disease.

**Methods:** All SARS-CoV-2-positive adults in Iceland were prospectively enrolled into a telehealth service at diagnosis. A multivariable proportional-odds logistic regression model was derived from information obtained during the enrollment interview with those diagnosed before May 1, 2020 and validated in those diagnosed between May 1 and December 31, 2020. Outcomes were defined on an ordinal scale; no need for escalation of care during follow-up, need for outpatient visit, hospitalization, and admission to intensive care unit (ICU) or death. Risk factors were summarized as odds ratios (OR) adjusted for confounders identified by a directed acyclic graph.

**Results:** The prognostic model was derived from and validated in 1,625 and 3,131 individuals, respectively. In total, 375 (7.9%) only required outpatient visits, 188 (4.0%) were hospitalized and 50 (1.1%) were either admitted to ICU or died due to complications of COVID-19. The model included age, sex, body mass index (BMI), current smoking, underlying conditions, and symptoms and clinical severity score at enrollment. Discrimination and calibration were excellent for outpatient visit or worse (C-statistic 0.75, calibration intercept 0.04 and slope 0.93) and hospitalization or worse (C-statistic 0.81, calibration intercept 0.16 and slope 1.03). Age was the strongest risk factor for adverse outcomes with OR of 75-compared to 45-year-olds, ranging from 5.29-17.3. Higher BMI consistently increased the risk and chronic obstructive pulmonary disease and chronic kidney disease correlated with worse outcomes.

**Conclusion:** Our prognostic model can accurately predict the outcome of SARS-CoV-2 infection using information that is available at the time of diagnosis.

## Introduction

Coronavirus disease 2019 (COVID-19), caused by the severe acute respiratory syndrome-coronavirus-2 (SARS-CoV-2), was first described in Wuhan in December 2019 and was declared a pandemic on March 11, 2020.[1] The severity of COVID-19 ranges from asymptomatic infection to severe respiratory failure and death. While early reports suggested that most infections were severe, later studies found that 81% of those who were symptomatic had mild disease, 14% had severe disease, and 5% developed critical illness.[2] The infection fatality rate has been estimated to be between 0.26-0.66%.[3, 4]

From the first diagnosed case of COVID-19 in Iceland on February 27, 2020 until December 31, 2020, a total of 6,126 persons tested SARS-CoV-2-positive by quantitative reverse-transcriptase polymerase chain reaction (qPCR). Broad access to qPCR testing was introduced early in the pandemic for both symptomatic and asymptomatic persons.[5] All individuals who tested positive were enrolled into a telehealth service provided by the COVID Outpatient Clinic of Landspitali–The National University Hospital of Iceland (LUH).[6] The resulting cohort of SARS-CoV-2-positive cases was prospectively followed from the date of diagnosis. A national seroprevalence study found that a large proportion (56%) of seropositive individuals had been identified by qPCR testing during the first-wave of the epidemic in Iceland.[7]

The relationship between individual baseline characteristics and subsequent disease severity is an important consideration for the allocation of public health resources during the pandemic. Male sex, advanced age[8] and obesity[9, 10] have been associated with increased disease severity and diabetes, cardiovascular and cerebrovascular diseases, chronic kidney disease, cancer and preexisting lung disease have been correlated with poorer outcomes.[11–13] Moreover, dyspnea, fever, and gastrointestinal symptoms have been associated with a more severe disease course. However, many studies to date include a highly selected group of patients with COVID-19 who require hospitalization and therefore do not provide actionable prognostic predictions for risk stratification of patients early in the course of their illness.

While extensive testing is desirable for contact tracing and isolation of infected people, large numbers of cases can easily overwhelm the capacity of the healthcare system for provision of clinical care. Early risk stratification offers opportunities for triaging SARS-CoV-2-positive persons to appropriate levels of monitoring and intervention. Additionally, knowledge of risk factors for severe disease can assist in prioritising individuals for vaccination. The aim of this study was twofold. Firstly, to develop and validate a multivariable model to predict the prognosis of SARS-CoV-2-positive adults at the time of diagnosis. Secondly, to determine which baseline characteristics are risk factors for severe COVID-19.

## Methods

### Ethical approval

The study was approved by the National Bioethics Committee of Iceland (VSN 20-078).

### Study population

The study population included all individuals who tested positive for SARS-CoV-2 by qPCR in Iceland between February 27 and December 31, 2020. Three national testing programs were implemented during the study period; targeted testing based on clinical suspicion (from February 1), open invitation population screening (from March 13) and mandatory screening at the border (from June 15). Those who tested positive at border screening underwent antibody testing, a repeat qPCR test of a nasopharyngeal swab and were assessed regarding symptoms and exposure. Active infections were diagnosed based on these data. All persons who were SARS-CoV-2-positive and considered to have an active infection were enrolled into the telehealth service of the LUH COVID Outpatient Clinic until uneventful termination of telehealth follow-up care, hospital discharge or death from COVID-19.

### The COVID-19 Outpatient Clinic

The COVID-19 Outpatient Clinic coordinated the outpatient care of all SARS-CoV-2-positive persons in Iceland as described previously.[6] Data regarding underlying conditions, medication use, clinical symptoms and severity of the infection were prospectively recorded using a standardized data entry form starting on March 17. During each interview, the presence of 19 specific symptoms was documented. Patients were evaluated and assigned a clinical severity score by the interviewing physician or nurse, based on the combination of their clinical judgment and patients’ symptoms. The clinical severity scores were generated by infectious disease consultants at LUH at the beginning of the pandemic: low severity, defined as having mild or no symptoms; moderate severity, defined as mild dyspnea, cough or fever for less than five days; and high severity, defined as severe dyspnea, worsening cough and high or persistent fever for five days or longer. Patients with alarming symptoms were transported to the COVID Outpatient Clinic for in-person evaluation. Patients were discharged from the telehealth service when at least 14 days had passed from qPCR diagnosis and seven days had passed from resolution of symptoms.

### Data sources

In addition to the prospectively collected information that was obtained from telehealth interviews, data were obtained from several population-based registries in Iceland. All International Classification of Diseases, 10th Revision (ICD-10) diagnostic codes recorded ≥14 days prior to the persons’ first positive qPCR for SARS-CoV-2 were obtained from three registries: LUH patient registry (from 2009), the Register of Primary Health Care Contacts (from 2004) and the Register of Contacts with Medical Specialists in Private Practice (from 2010). Data on all filled drug prescriptions were retrieved for each individual from the Prescription Medicines Register for the period ranging from 395 days (13 months) to 14 days before the first positive qPCR for SARS-CoV-2. Finally, all measurements of serum creatinine between January 2010 and until 14 days before the individual’s first positive qPCR were extracted from a centralized laboratory database. Estimated glomerular filtration rate (eGFR) was calculated using the Chronic Kidney Disease Epidemiology Collaboration (CKD-EPI) equation.[14]

### Prognostic model development

A prognostic model was developed to predict the clinical outcome of SARS-CoV-2-positive adults who were 18 years of age or older at first contact with the healthcare system following confirmed diagnosis. The model was derived using the cohort of SARS-CoV-2-positive persons who were diagnosed between February 27 and April 30. Persons without an Icelandic national identification number, those who were not considered to have an active infection and those living in a nursing home or who were admitted to hospital at the time of diagnosis were excluded (Figure 1). A formal sample size calculation was not performed because all available data were used. A proportional-odds logistic model was used and outcomes were defined on an ordinal scale, ranging from the least to the most severe; (1) absence of clinical deterioration requiring in-person evaluation or hospitalization for the duration of telehealth care; (2) clinical deterioration requiring in-person evaluation at the LUH COVID-19 Outpatient Clinic, but not subsequent hospitalization; (3) hospitalization; and (4) admission to intensive care unit (ICU) or death. Predictor variables were derived from the prospectively recorded data obtained during the enrollment interview into the telehealth service. Age was modelled as a non-linear variable using a restricted cubic spline with knots placed at the 0.05, 0.35, 0.65 and 0.95 percentiles. Body mass index (BMI) was included as a linear variable and all other predictors were dichotomous. All decisions regarding predictors and model development were made prior to examining the outcome data and were based on the prior literature and clinical expertise of the study authors. No statistical selection procedures or model comparisons were employed. The included predictor variables are shown in Table 1. Missing data were imputed 100 times using multiple imputation with chained equations (MICE) and predictive mean matching using the *aregImpute* function.[15]

**Figure 1.**
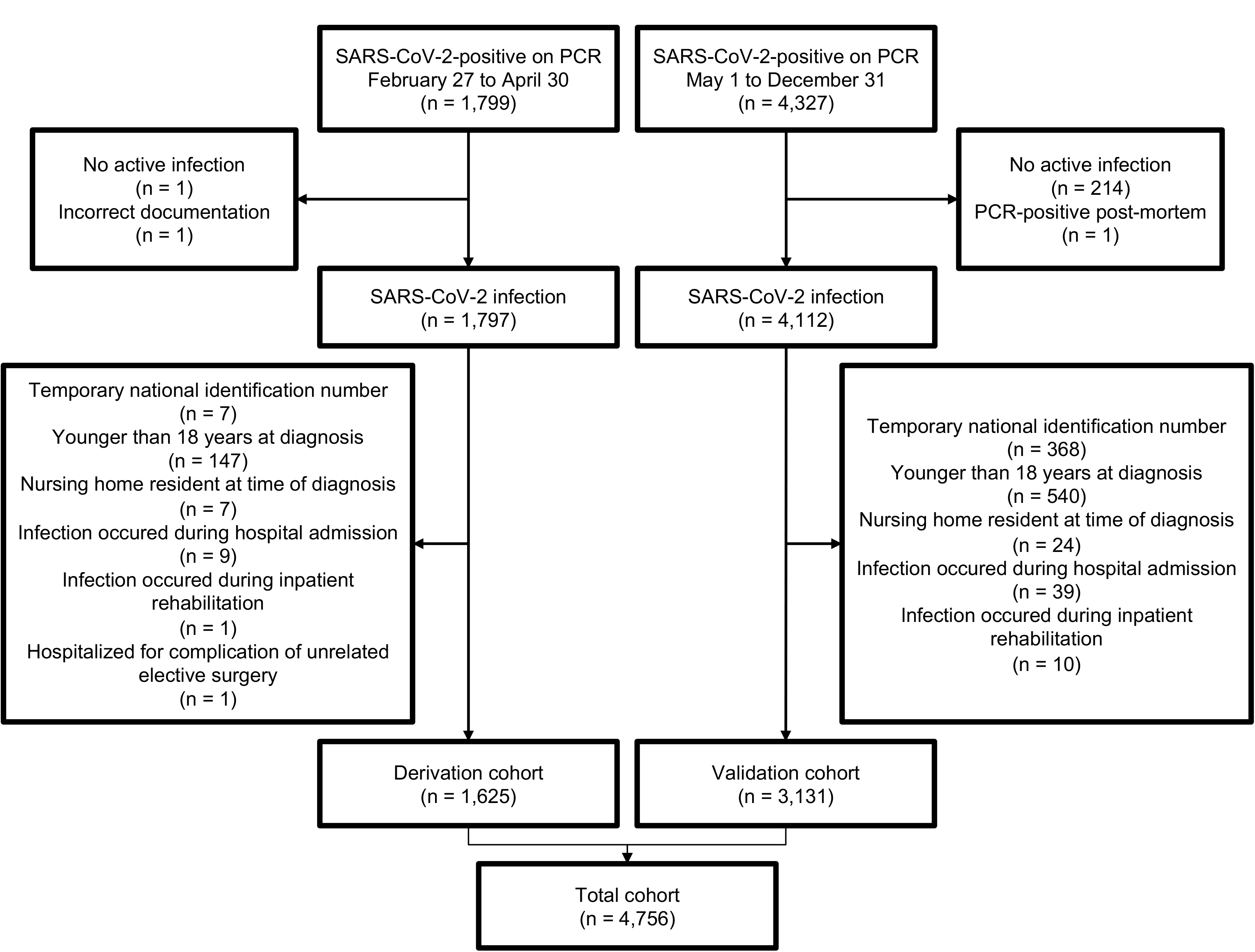
Flow diagram of the study.

**Table 1.** Variables included in the prognostic model are shown for the derivation and validation cohorts. Continuous variables are summarised as medians and interquartile ranges (IQR). The number of cases behind each categorical variable are presented along with the proportion. For each of the variables, the number and proportion of cases with missing data are displayed within parentheses.

|  | Median and interquartile range for continuous<br>or *n* and % for categorical<br>(*n* missing and %) |  |  |
| --- | --- | --- | --- |
| Predictor | Derivation cohort<br>*n* = 1,625 | Validation cohort<br>*n* = 3,131 | p-<br>value |
| Age | 45, 31-56<br>(0, 0%) | 37, 27-53<br>(0, 0%) | <0.001 |
| Sex = Male | 799, 49.2%<br>(0, 0%) | 1,656, 52.9%<br>(0, 0%) | 0.016 |
| Body mass index | 26.2, 23.4-30.1<br>(1,115, 68.6%) | 25.9, 23.1-29.4<br>(326, 10.4%) | 0.075 |
| Current smoking | 84, 6.1%<br>(246, 15.1%) | 311, 10.5%<br>(158, 5.0%) | <0.001 |
| Prior smoking | 322, 41.1%<br>(841, 51.8%) | 931, 44.8%<br>(1,055, 33.7%) | 0.076 |
| Diabetes | 49, 3.0%<br>(11, 0.7%) | 87, 2.8%<br>(40, 1.3%) | 0.735 |
| Hypertension | 229, 14.2%<br>(11, 0.7%) | 340, 11.0%<br>(40, 1.3%) | 0.002 |
| Heart disease | 121, 7.5%<br>(11, 0.7%) | 161, 5.2%<br>(40, 1.3%) | 0.002 |
| Chronic kidney disease | 12, 0.7%<br>(11, 0.7%) | 13, 0.4%<br>(40, 1.3%) | 0.217 |
| Pulmonary disease | 127, 7.9%<br>(11, 0.7%) | 118, 3.8%<br>(40, 1.3%) | <0.001 |
| Cancer | 60, 3.7%<br>(11, 0.7%) | 54, 1.7%<br>(40, 1.3%) | <0.001 |
| Flu-like symptoms | 1,446, 90.9%<br>(35, 2.2%) | 2,335, 75.5%<br>(40, 1.3%) | <0.001 |
| Upper respiratory symptoms | 1,135, 75.4%<br>(119, 7.3%) | 1,592, 51.5%<br>(40, 1.3%) | <0.001 |
| Lower respiratory symptoms | 649, 44.6%<br>(170, 10.5%) | 557, 18.0%<br>(40, 1.3%) | <0.001 |
| Gastrointestinal symptoms | 505, 35.1%<br>(187, 11.5%) | 513, 16.6%<br>(40, 1.3%) | <0.001 |
| Clinical score = moderate or high severity | 293, 18.4%<br>(36, 2.2%) | 177, 6.2%<br>(270, 8.6%) | <0.001 |
| Telehealth only | 1,363, 83.9%<br>(0, 0%) | 2,780, 88.8%<br>(0, 0%) | <0.001 |
| Outpatient visit | 162, 10.0%<br>(0, 0%) | 213, 6.8%<br>(0, 0%) |  |
| Hospitalization | 71, 4.4%<br>(0, 0%) | 117, 3.7%<br>(0, 0%) |  |
| Intensive care unit admission<br>or death | 29, 1.8%<br>(0, 0%) | 21, 0.7%<br>(0, 0%) |  |

The prognostic model was validated in the cohort diagnosed between May 1 and December 31. Multiple imputation was performed separately for the derivation and validation cohorts. Confidence intervals were obtained by 2000 bootstrap resamplings of a randomly selected imputed dataset.[16] Model discrimination was quantified using the C-statistic. Calibration was assessed by visual examination of calibration plots and via several calibration indices. The sensitivity, specificity, positive predictive value (PPV) and negative predictive value (NPV) of the model in the validation cohort were reported for each outcome over a range of probabilistic thresholds. The potential impact of implementing the model in the COVID Outpatient Clinic was explored by evaluating the number of interviews that would have been prevented if those who had a predicted risk of requiring an outpatient visit or worse below a given threshold, received only two interviews (at enrollment and discharge) instead of the observed number of interviews. In addition, decision curve analysis was performed to quantify the net benefit of the prognostic model over a range of plausible probability thresholds for use in two clinical scenarios: (1) to aid in determining whether an individual‘s risk of clinical deterioration requiring an outpatient visit or worse is sufficiently small to omit from the telehealth service; and (2) to aid in deciding whether an individual’s risk of hospitalization or worse is sufficiently large to recommend more intensive follow-up or therapeutic intervention. The model validation and decision curve analysis are described in more detail in the Supplementary Methods.

The development and validation of the prognostic model conformed with the Transparent Reporting of a Multivariable Prediction Model for Individual Prognosis or Diagnosis (TRIPOD) guidelines.[17] Although the model derivation and validation are published together, the model was derived before the validation data were collected. Only two changes were made to the model following the collection of validation data (but prior to validating the model): CKD was dropped as a predictor variable, and moderate and high clinical severity scores were combined, in both instances due to few patients having these predictors in the validation cohort.

### Risk factor analysis

The relationship between specific baseline characteristics and severity of COVID-19 was explored using the entire cohort of SARS-CoV-2-positive adults after applying the same exclusion criteria as above (Figure 1). Underlying conditions (diabetes, hypertension, ischemic heart disease [IHD], chronic obstructive pulmonary disease [COPD] and cancer) were defined based on the presence of a compatible ICD-10 diagnosis codes in the population-based registries and a prescription for medication consistent with the condition that was filled during the preceding 13 months. Chronic kidney disease was defined as eGFR <60 mL/min/1.73 m^2^ on at least two separate occasions ≥90 days apart. Determinations of eGFR obtained during an episode of acute kidney injury were excluded from the assessment of CKD.[18] Height, weight and past and current smoking history were retrieved from the standardized data entry form of the COVID-19 Outpatient Clinic. The precise definitions of all included variables are described in Supplementary Table 1. Missing data for BMI and past and current smoking history were multiply imputed.

A directed acyclic graph (DAG) was created to describe the hypothesized relationships between the exposure variables and severity of COVID-19 and was used to define the minimally sufficient adjustment set needed to estimate the association of each variable with the outcome (Supplementary Figure 1).[19] Logistic regression models were fitted using dichotomization of the ordinal outcomes of the prognostic model. The relationship between the variables and the outcomes were reported as univariate and adjusted odds ratios (OR) with 95% confidence intervals (CI). In the case of continuous variables, the OR between informative cut-offs were reported as chosen by the study authors.

All statistics were performed in R version 3.6.3.[20] using the *tidyverse* package for data manipulation. The *cowplot* package[21] was used to create multipanel figures and *tableone*[22] was used to create summary tables. All statistical code is available at https://osf.io/t2bp8/.

## Results

### Study population

Of the 175,243 individuals who were tested for SARS-CoV-2 using qPCR in Iceland during the study period, 6,126 were positive. After applying exclusion criteria, 1,625 individuals were included in the derivation cohort and 3,131 in the validation cohort (Figure 1). Compared to the derivation cohort, the median age of the validation cohort was younger (37 years compared to 45 years, p <0.001), the proportion of males higher (53% compared to 49%, p = 0.016), and the proportion of persons reporting each of the prospectively recorded underlying conditions lower, except for diabetes (Table 1). In total, 4,143 (87%) patients never required in-person visit for evaluation during telehealth follow-up, 375 (7.9%) only required outpatient visits, 188 (4.0%) required admission to hospital but no further escalation of care and 50 (1.1%) either required admission to ICU or died due to complications of COVID-19. The median time from the first positive qPCR to telehealth enrollment was 0 days (IQR, 0-1, range 0-4), and the median follow-up time was 15 days (IQR, 14-16, range, 7-67). No patient was lost to follow-up.

### Prognostic model performance

The internal model calibration and discrimination for the derivation cohort are shown in Supplementary Figure 2 and Supplementary Table 2. On model validation, the discrimination was consistent for the three clinical outcomes with point estimates of the C-statistic ranging from 0.79 to 0.85 (Table 2). Visual examination of calibration plots and calibration metrics were adequate for outpatient visit or worse (calibration intercept, 0.03 [95%CI, -0.10 to 0.14], calibration slope 0.94 [95%CI, 0.81 to 1.07]) and hospitalization or worse (calibration intercept, 0.15 [95%CI, -0.03 to 0.33], calibration slope, 1.04 [95%CI, 0.91 to 1.17]), but not for ICU admission or death (calibration intercept, -0.6 [95%CI, -1.2 to -0.28], calibration slope, 0.77 [95%CI, 0.61 to 0.97]) (Table 2, Figure 2).

**Figure 2.**
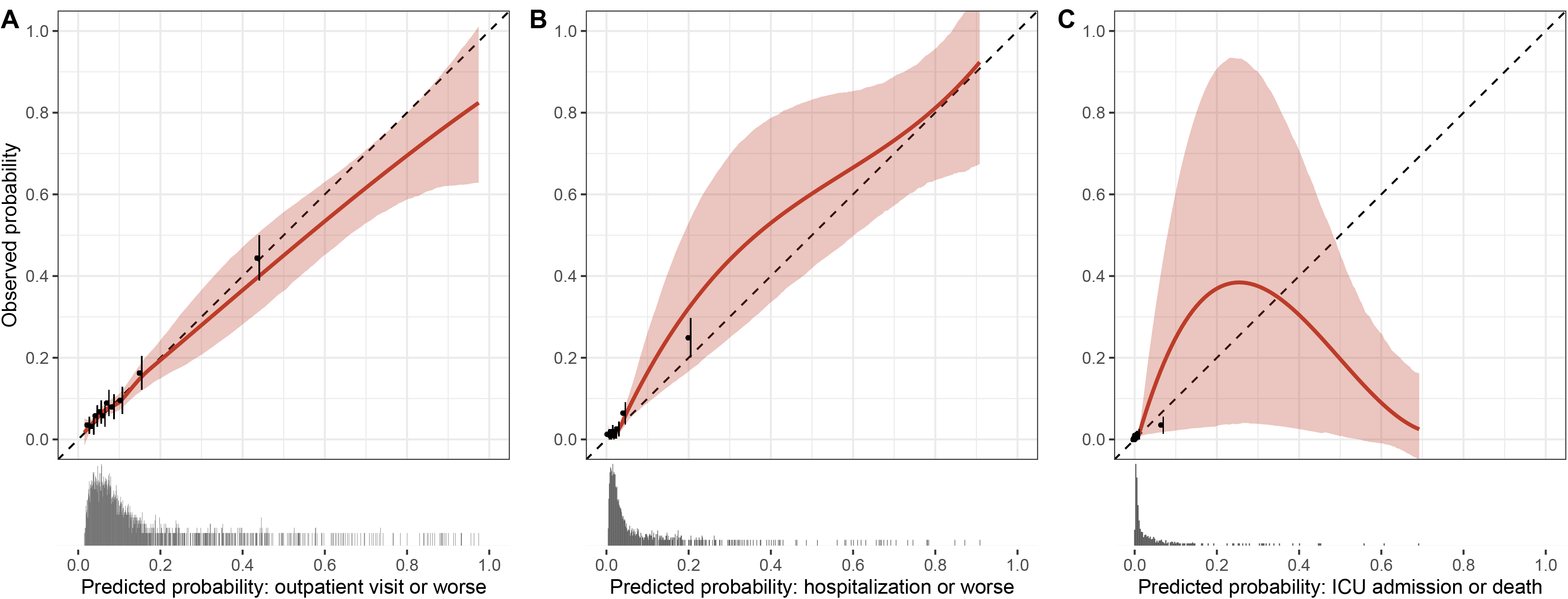
Calibration curves of the prognostic model using the validation cohort illustrate the relationship between the observed and predicted probability of outpatient visit or worse (Panel A), hospitalization or worse (Panel B) and admission to intensive care unit or death (Panel C). The sample distribution of predicted probabilities is shown with marginal histograms. The sample is divided into 10 equally large groups and the aggregate relationship between observed and predicted probability is depicted as a black dot and pointrange. The weighted scatterplot smoothing (LOWESS) relationship between the observed and predicted probabilities is shown as a red line with the shaded area representing 95% confidence intervals based on bootstrap resampling. These are compared to the dashed black line, reflecting a perfect relationship between observed and predicted probabilities.

**Table 2.**
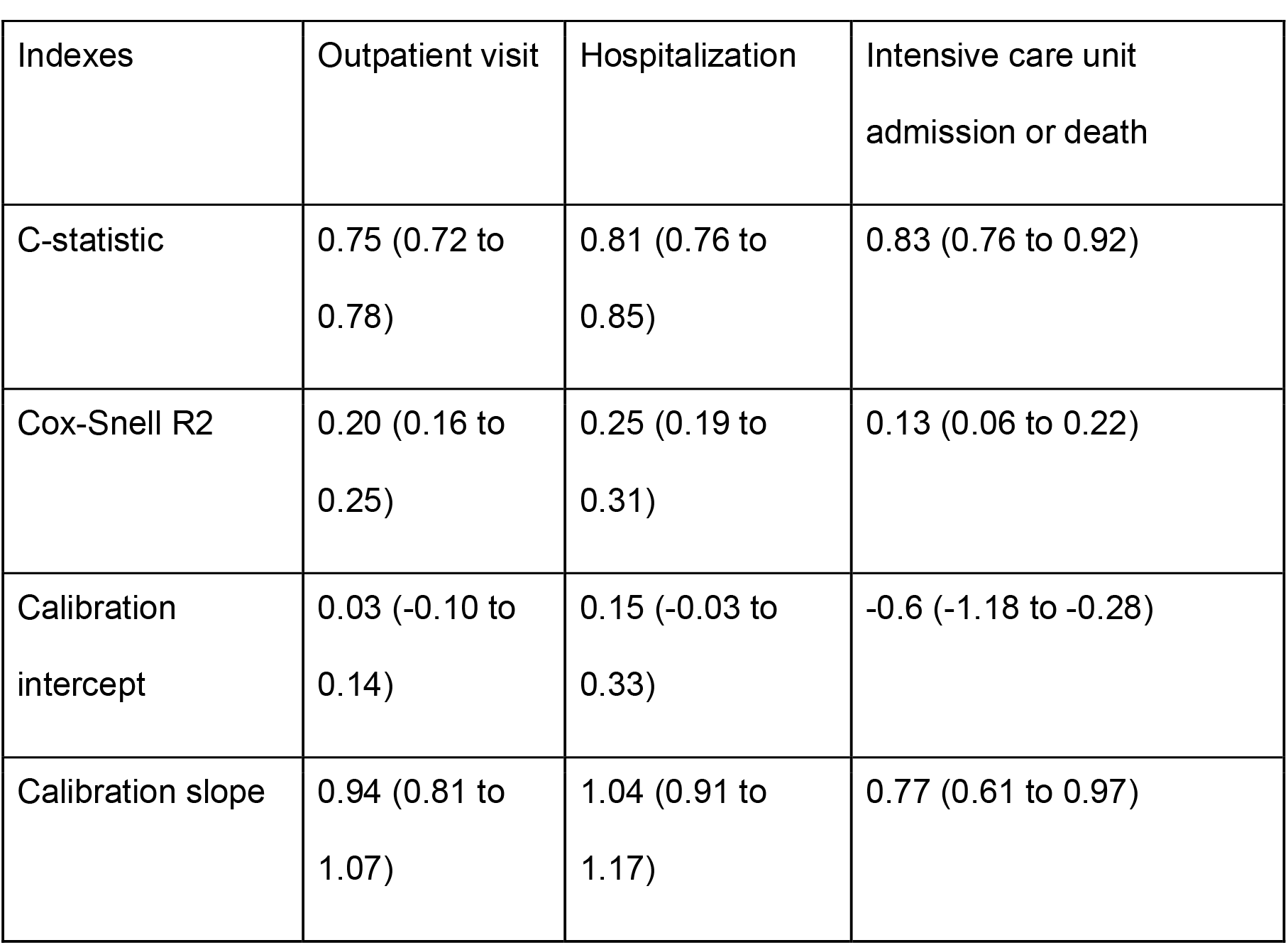
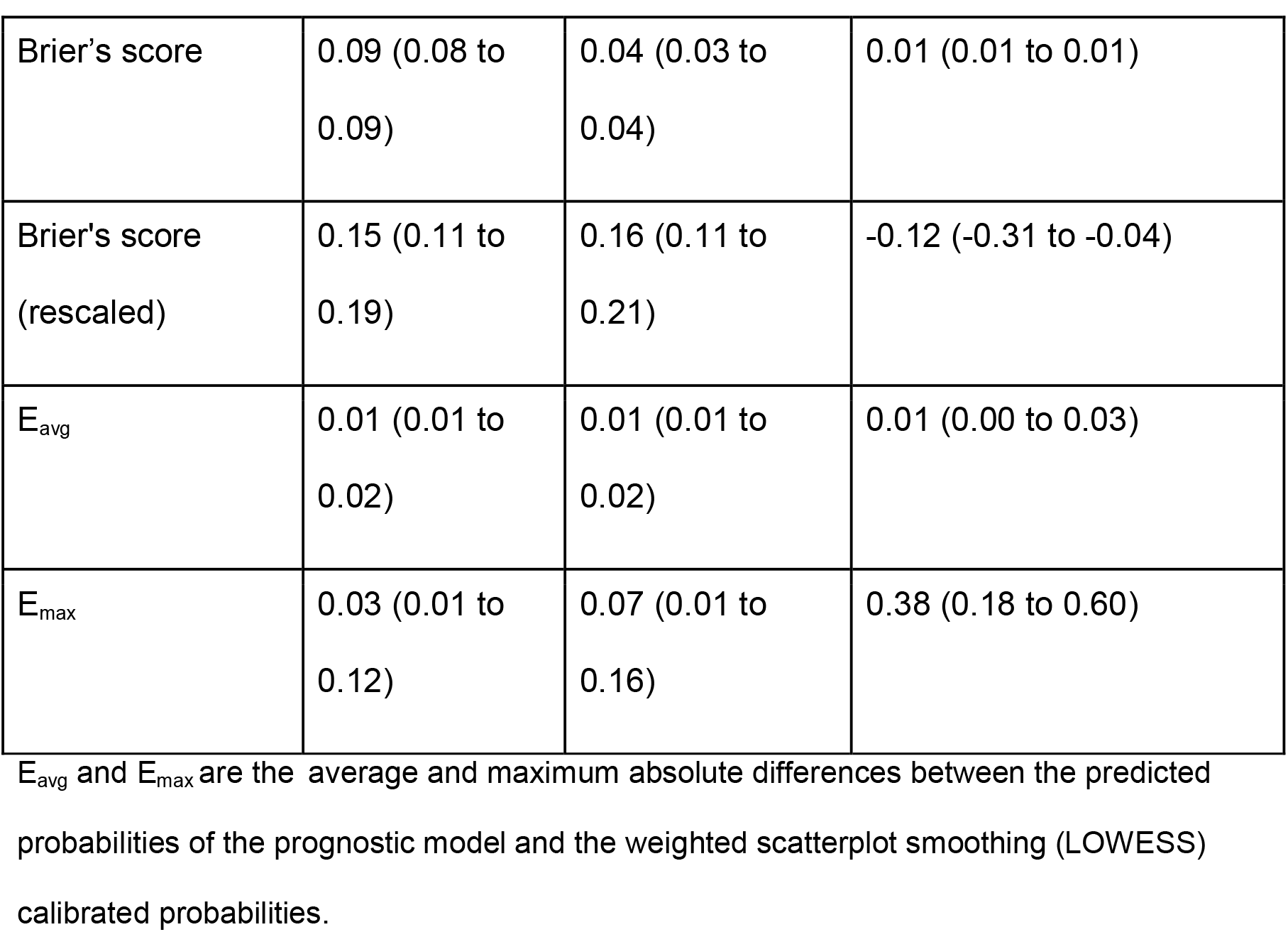
Calibration and discrimination indices of the prognostic model for the validation cohort for each of the outcomes. The 95% bootstrapped confidence intervals are presented within parentheses.

| Indexes | Outpatient visit | Hospitalization | Intensive care unit admission or death |
| --- | --- | --- | --- |
| C-statistic | 0.75 (0.72 to 0.78) | 0.81 (0.76 to 0.85) | 0.83 (0.76 to 0.92) |
| Cox-Snell R2 | 0.20 (0.16 to 0.25) | 0.25 (0.19 to 0.31) | 0.13 (0.06 to 0.22) |
| Calibration intercept | 0.03 (-0.10 to 0.14) | 0.15 (-0.03 to 0.33) | -0.6 (-1.18 to -0.28) |
| Calibration slope | 0.94 (0.81 to 1.07) | 1.04 (0.91 to 1.17) | 0.77 (0.61 to 0.97) |
| Brier's score | 0.09 (0.08 to 0.09) | 0.04 (0.03 to 0.04) | 0.01 (0.01 to 0.01) |
| Brier's score (rescaled) | 0.15 (0.11 to 0.19) | 0.16 (0.11 to 0.21) | -0.12 (-0.31 to -0.04) |
| E<sub>avg</sub> | 0.01 (0.01 to 0.02) | 0.01 (0.01 to 0.02) | 0.01 (0.00 to 0.03) |
| E<sub>max</sub> | 0.03 (0.01 to 0.12) | 0.07 (0.01 to 0.16) | 0.38 (0.18 to 0.60) |

E<sub>avg</sub> and E<sub>max</sub> are the average and maximum absolute differences between the predicted probabilities of the prognostic model and the weighted scatterplot smoothing (LOWESS) calibrated probabilities.

The lower 95% confidence limit for the NPV of requiring an outpatient visit or worse was maximized at a threshold of 4.1% predicted risk. At this threshold, the sensitivity of the need for outpatient visit or worse was 94.9% (95%CI, 92.0 to 96.7), specificity was 20.2% (95%CI, 18.7 to 21.7), PPV was 13.0% (95%CI, 11.8 to 14.4) and NPV was 96.9% (95%CI, 95.1 to 98.0). In the validation cohort, 585 (18.7%) patients had this level of predicted risk or lower. Using a threshold of 10% predicted risk, the sensitivity was 61.5% (95%CI, 56.4 to 66.4), specificity was 76.3% (95%CI, 74.7 to 77.8), PPV was 24.7% (95%CI, 21.9 to 27.7) and NPV was 94.0% (95%CI, 92.9 to 94.9). This degree of predicted risk or lower was observed for 2,268 (72.4%) persons in the validation cohort. The performance of the model as a function of continuous predicted risk is shown in Supplementary Figure 3.

### Prognostic model usage

The possible impact of implementing the prognostic model in the clinical pathway of the telehealth service provided by the COVID Outpatient Clinic is shown in Figure 3. If individuals in the validation cohort who had ≤4.1% predicted risk of outpatient visit or worse were triaged during the enrollment interview to receive only one additional telehealth interview seven days later, the prognostic model would have averted 2,643 interviews (13.4% of all interviews) with 565 persons (18.0% of all persons). Of those, 5.2% (11 out of a total of 213 in the validation cohort) would later require an outpatient visit for evaluation, 3.4% (four out of 117) would be admitted to hospital and none would die or require admission to ICU. For the decision to omit from the telehealth service, decision curve analysis revealed that the net benefit of the prognostic model compared with the strategy of enrolling all patients ranged from 0.03 to 0.33 over a reasonable span of low-risk thresholds (4% to 10%) (Panel A of Supplementary Figure 4). Similarly, for the decision to recommend more rigorous management, the net benefit of the model compared with a strategy of no enrollment into close follow-up ranged from 0.34 to 0.49 over a reasonable scope of high-risk thresholds (5% to 10%) (Panel B of Supplementary Figure 4).

**Figure 3.**
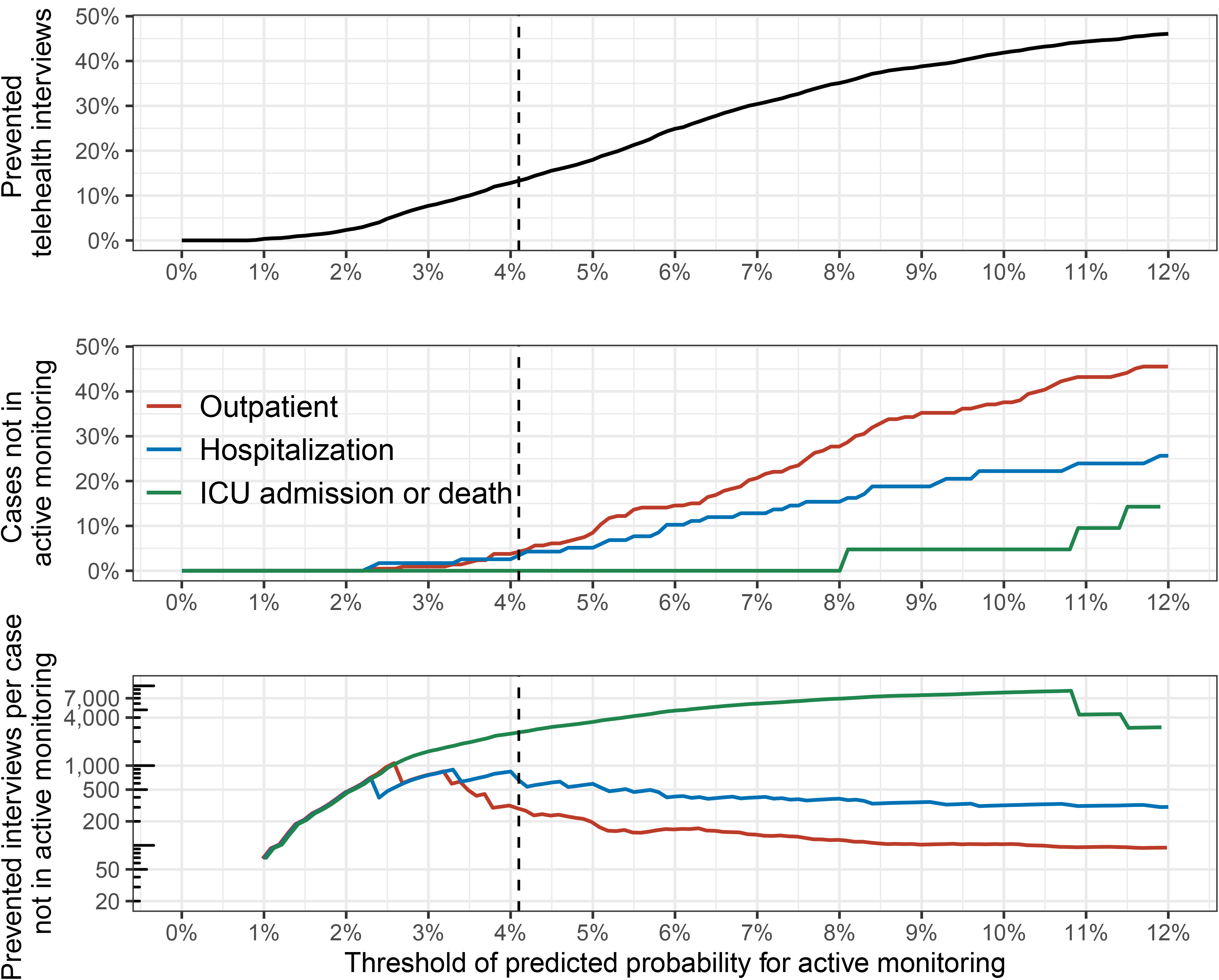
Potential benefits and harms of implementing the prognostic model to triage into a telehealth service, using different thresholds for the predicted probability of the need for an outpatient visit or worse. The top panel illustrates the proportion of telehealth interviews that would have been avoided if those who had a predicted probability below the given threshold received only two (enrollment and discharge) interviews. The middle panel depicts the proportion of individuals who required escalation of care that would have received fewer interviews using the corresponding threshold. The bottom panel illustrates the ratio between these potential benefits and harms. The threshold at which the lower confidence limit of the negative predictive value of outpatient visit or worse is maximized (4.1%) is shown as a vertical dotted line.

### Risk factor analysis

The results of the risk factor analysis are shown in Table 3. Age was the strongest risk factor for experiencing each of the study outcomes with point estimates of odds ratios for persons 75 years of age compared with 45 years of age ranging from 5.29 to 17.3 (Supplementary Figure 5). Male sex decreased the risk of outpatient visit or worse (OR, 0.68; 95%CI, 0.57 to 0.81), but did not significantly increase the risk of hospitalization or worse (OR, 1.14; 95%CI 0.87 to 1.48) or admission to ICU or death (OR, 1.68; 95%CI 0.94 to 2.99). Higher BMI consistently increased the risk of the study outcomes with point estimates of the OR between individuals with BMI of 35 kg/m^2^ compared with 25 kg/m^2^ ranging from 1.98 to 5.02 (Supplementary Figure 6). No significant interaction was found between age and BMI (Supplementary Figure 7). The effects of comorbidities were generally modest (point estimates of the OR ranging from 0.80 to 2.94) with the exception of the OR of COPD for admission to ICU or death, which was 5.33 (95% CI, 2.11 to 13.5). Current smoking appeared to be associated with a lower likelihood of experiencing any of the outcomes in both univariate and adjusted analysis.

**Table 3.**
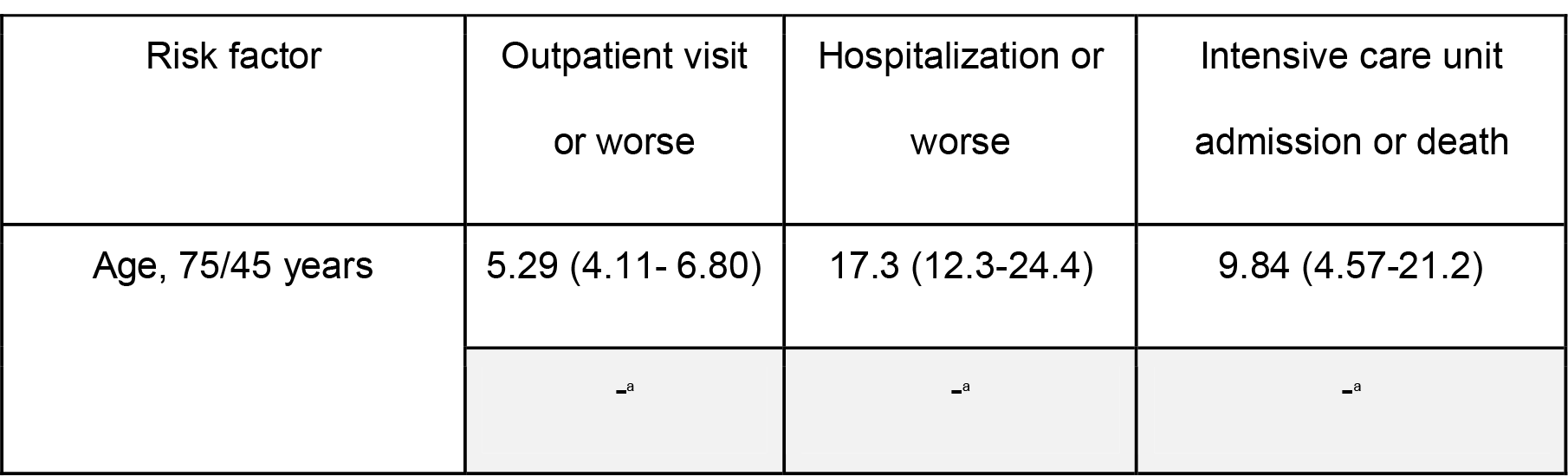
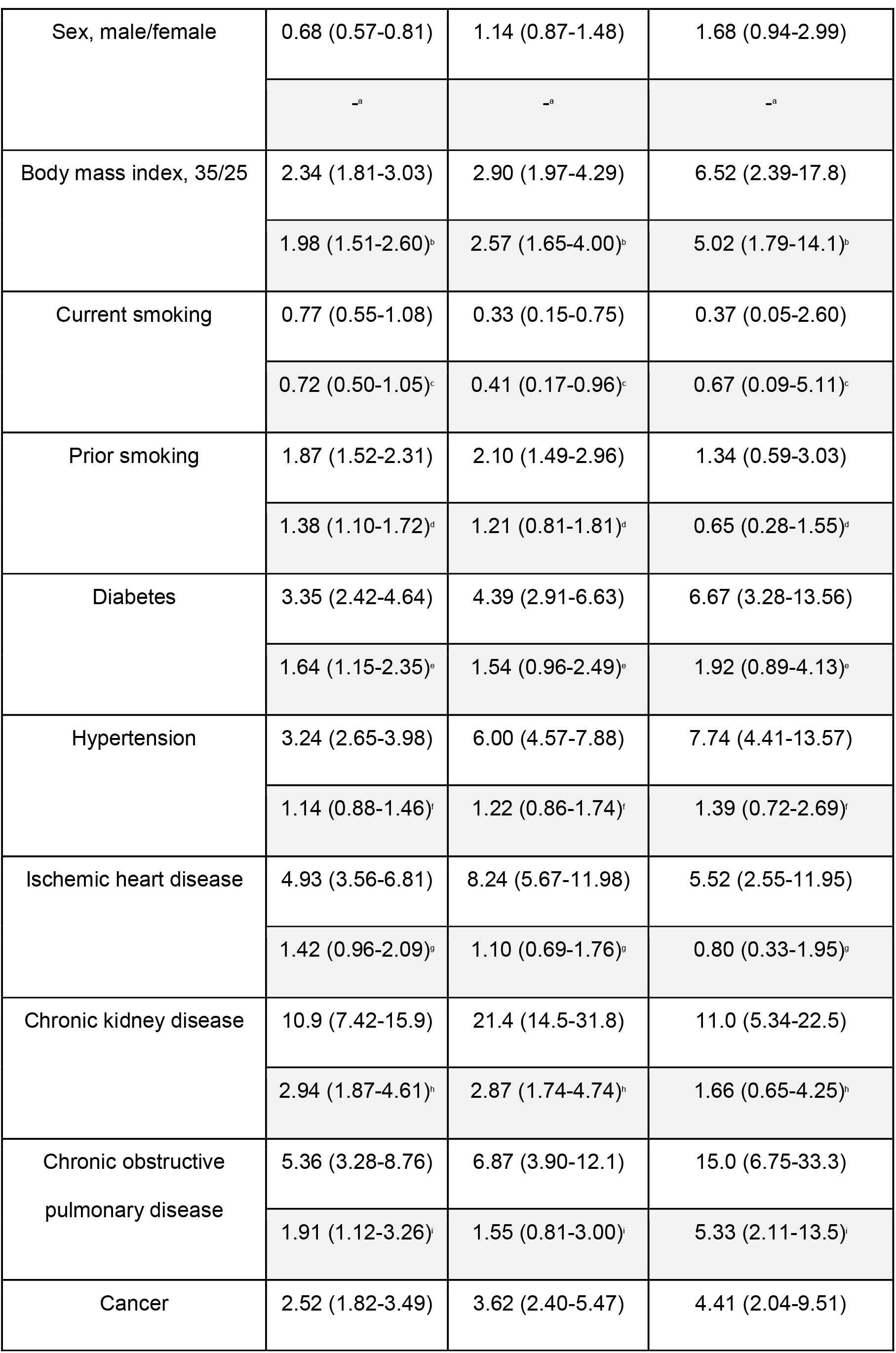
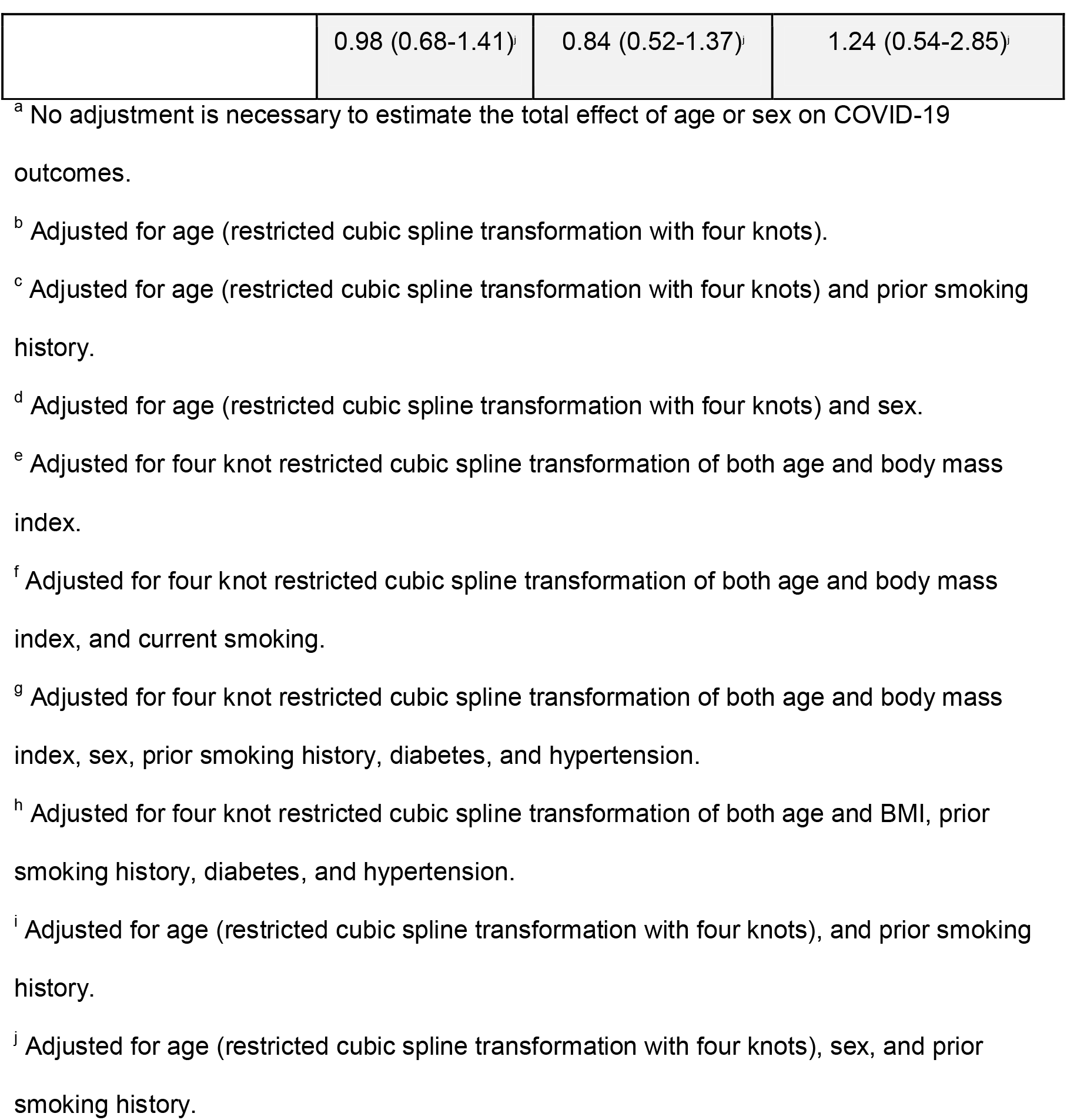
Univariate and adjusted odds ratios (OR) of adverse outcomes of COVID-19 for each risk factor and outcome measure. The hypothesized relationship between the risk factors and outcomes were stated as a directed acyclic graph (Supplementary Figure 1). Odds ratios were adjusted for the minimally sufficient set of confounders needed to estimate the total effect of the risk factor on the outcome given this hypothesized relationship. Adjusted odds ratios are shown in the shaded rows. The 95% confidence interval of the odds ratios are displayed within parentheses.

## Discussion

In this study, a prognostic model was derived and validated in prospective population-based cohorts of SARS-CoV-2-positive adults. We demonstrate how the model could be implemented in the context of a telehealth service that obtains all the necessary information for prognostication through interviews and triages patients to the appropriate level of care. The results of risk factor analysis are in line with previous reports of conditions associated with increased risk of adverse outcomes.

The COVID-19 pandemic has strained and, in many cases, overwhelmed national healthcare systems due to the large number of seriously ill patients requiring advanced medical care. High rates of virologic testing and isolation of positive individuals can assist in curtailing disease spread, but increases the number of identified cases. While SARS-CoV-2 infection can lead to severe disease and death, most cases are mild or asymptomatic. Predicting who will subsequently require comprehensive care and, equally importantly, identifying those who will only need minimal or no further follow-up is challenging. However, potential solutions do exist, including optimization of healthcare resources allocation and reduction of the overall healthcare burden using early interventions. Risk stratifying patients in a pandemic setting with soaring case counts requires efficient and robust methods that should ideally be implemented without the need for close contact or extensive clinical testing. Limited evidence supporting such stratification is currently available.

The prognostic model in our study was derived and validated in large community-based cohorts of adults. Both cohorts are considerably larger than most COVID-19 cohorts used for previously developed models, in which the median size of the derivation cohorts was 338 and the median size of the validation cohorts was 189.[23] On validation, the model was found to have good ability to discriminate between persons who either will or will not experience each of the three outcomes, as reflected by high C-statistics. The agreement between observed and predicted probabilities of the outcomes was also excellent with respect to the need for an outpatient visit or worse, and hospitalization or worse, for several different calibration metrics and by visual examination of the calibration plots (Table 2, Figure 2). Most notably, the E_avg_ and E_max_ values (average and maximum absolute differences between the predicted probabilities and the LOWESS calibrated probabilities) were 0.9% and 3.2% for outpatient visit or worse and 1.1% and 6.8% for hospitalization or worse, respectively. In our validation cohort, the calibration of the need for admission to ICU or death was poor, as is demonstrated by visual examination of the calibration plot and an E_max_ of 38%, among other metrics. This could be explained, at least in part, by therapeutic advances made in the inpatient treatment of COVID-19 which primarily benefitted patients in the validation cohort.[24, 25]

In addition to demonstrating good discrimination and calibration of the prognostic model, we report the clinical utility of the model with a case study of the telehealth service provided by the LUH COVID-19 Outpatient Clinic and with decision curve analysis. We show that by using a threshold of 4-10% predicted risk for outpatient visit or worse to triage into the telehealth service, a relatively large proportion of interviews could have been avoided without missing many individuals who needed more advanced care. Decision curve analysis similarly showed that the model led to increased net benefit when deciding whether follow-up of patients is needed or not and determining which cases will require even closer surveillance. The optimal thresholds at which to use the prognostic model for resource allocation will vary based on local factors, such as the number of patients that are diagnosed in relation to the capacity of the healthcare infrastructure and the costs and benefits of the treatments or monitoring strategies being considered.

A strong practical advantage of this prognostic model is the ability to estimate the risk of several outcomes using predictors that can be collected via telephone interview at the time of diagnosis, as opposed to conventional clinical assessment that includes physical examination, laboratory testing and imaging studies. The model was derived and validated in the setting of easy access to diagnostic testing for SARS-CoV-2 and included all nonhospitalized adults who tested positive in Iceland. The risk of selection bias is therefore minimal. Furthermore, data were invariably obtained by trained healthcare providers within 24 hours of the individual’s first positive qPCR using a standardized questionnaire, thereby minimizing recall bias. A systematic review by Wynants et al[23] compared 107 prognostic models for predicting the need for interventions, progression to severe disease, and mortality from COVID-19. The authors did not recommend any of these models for current use due to risk of bias. A common cause of bias was that many patients were excluded because they had not developed an outcome by the end of the study period, i.e. had not recovered, been discharged from hospital, or died. In our study, follow-up of all cases was complete with a median duration of 15 days and up to 64 days for patients with persistent symptoms. Finally, the derivation, validation, and reporting of the prognostic model adhered to the TRIPOD guidelines. All of the above support the generalizability of our model to other settings.

In the risk factor analysis that was based on the entire cohort of 4,756 SARS-CoV-2-positive persons, age was most important. For hospitalization and ICU admission or death as outcome measures, the OR for this age difference was an order of magnitude higher than for all other risk factors. In the absence of prognostic models, age should guide decision-making regarding prioritization for vaccination and patient care. While male sex has consistently been linked to worse outcome in previous studies, our study did not support this observation. On the contrary, female sex was associated with an increased need for any medical intervention, though the point estimate of the odds ratio of worse outcome for males increased with greater severity. A robust association was found between increasing BMI and the more severe outcomes, which is consistent with prior research.[26] The point estimates of the effect sizes of comorbid conditions were generally modest and few were significantly associated with adverse outcome after adjusting for the minimally sufficient set of confounders. Although diabetes was linked to the need for any intervention, it was not associated wirh the more severe outcomes. This suggests that other factors than impaired glucose metabolism may mediate the increased risk among patients with high BMI, such as reduced lung function and prothrombotic state related to obesity.[10] Smoking was not associated with worse outcomes which is in line with previous studies, including meta-analyses.[27] It should be noted that only 7% of Icelanders are daily smokers[28], as was reflected in our cohort of SARS-CoV-2-positive persons. However, a diagnosis of COPD, which is usually associated with smoking, correlated with adverse outcomes. Finally, CKD was associated with more severe outcomes which is consistent with prior studies.[29]

While the strength of this study resides in the population-based approach, there are possible limitations that may decrease the generalizability of our findings. The clinical threshold at which an outpatient visit is considered to be indicated will likely vary between cultures and healthcare systems. This variation in clinical thresholds also exists for hospitalizations and ICU admissions, but is probably considerably less than for outpatient evaluation. In addition, the clinical severity score was loosely defined and was largely based on the clinical impression of the physician conducting the enrollment interview. Inter-rater variability was not quantified and we are therefore unable to speculate how this predictor will generalize to other settings. As the need for in-person evaluation at the COVID Outpatient Clinic was also determined by the same healthcare professionals during telephone interviews, it is perhaps not surprising that the clinical severity score was predictive. However, the clinical severity score during the enrollment interview also predicted the need for hospital admission, ICU care and death which downplays the possibility. Furthermore, a study examining the humoral response to SARS-CoV-2 in an overlapping cohort found a strong correlation between the clinical severity score and higher antibody levels, supporting the validity of the clinical severity score.[7]

## Conclusion

Accurate determination of risk factors for adverse outcomes in COVID-19 has broad implications for public health interventions, such as characterizing of populations at risk and orchestrating healthcare delivery. Based on the multivariable prognostic model generated by this study, it is possible to predict the risk of severe disease based on symptoms at diagnosis, comorbidities and demographic factors. These variables can be sampled by telephone interview at the time of diagnosis of COVID-19. This information may be valuable for risk stratification of patients at the time of diagnosis and prioritising health care resources.

## Declarations

### Ethics approval and consent to participate

The study was approved by the National Bioethics Committee of Iceland (VSN 20-078).

### Consent for publication

Not applicable.

### Availability of data and materials

The data that support the findings of this study are available from the corresponding author on reasonable request.

### Authors’ contributions

EE, VB, HLR, MIS, OSI, and RP developed the concept of the study. EE, VB, HLR, DH, HKB, LBO, SB, ASA, KO, HHT, and GK participated in aquiring data. EE, LBO, MIS, and RP were responsible for data curation. EE developed the methodology. EE, OSI, and RP analysed the data. MIS, OSI, and RP provided supervision. EE, VB, AB and OSI wrote the original draft of the manuscript. All authors reviewed and edited the manuscript. The corresponding author attests that all listed authors meet authorship criteria and that no others meeting the criteria have been omitted. RP is guarantor for the study.

## Supporting information

TRIPOD checklist

Statistical code and output

## Data Availability

https://osf.io/t2bp8/

## Acknowledgements

Not applicable.

