## Supplemental files for "Development and validation of a prognostic model for COVID-19: a population-based cohort study in Iceland"

**Supplementary Methods**

***Model discrimination and calibration indices***

Model discrimination was quantified using the C-statistic (the probability that a randomly selected patient who did experience the outcome has a higher predicted probability than a randomly selected patient who did not). Calibration was assessed by visual examination of calibration plots and by using several calibration indices: calibration intercept (assessment of mean risk with a target value of 0, such that negative values represent systematic overestimation of risk and positive values systematic underestimation of risk), calibration slope (assessment of estimated risk with a target value of 1, such that values <1 represent estimated risks that are too extreme [too high for high-risk patients and too low for low-risk patients] and values >1 represent estimated risks that are too conservative [too low for high-risk patients and too high for low-risk patients]), Brier score (the mean squared difference between the predicted probability of the outcome for each patient and his/her observed outcome, which ranges from 0 to 1, such that 0 indicates perfect accuracy and 1 perfect inaccuracy), rescaled Brier score (Brier score rescaled by the prevalence of the outcome to range between 0 and 1, such that 0 represents prediction accuracy that is no better than a random guess and 1 represents perfect accuracy), E_avg_ (the mean difference between the predicted probability of the outcome and the weighted scatterplot smoothing [LOWESS] calibrated probability) and E_max_ (the maximum absolute difference between the predicted probability of the outcome and the LOWESS calibrated probability).

***Decision curve analysis***

To aid in determining whether an individual’s risk of clinical deterioration requiring an outpatient visit or worse is sufficiently small to omit further telehealth follow-up, the net benefit of the prognostic model at a low-risk threshold L is defined as ${TNR}_{L}-\frac{p}{1-p}*\frac{1-L}{L}*{FNR}_{L}$ where *TNR_L_* is the true negative rate (the rate of patients classified as low-risk who did not require an outpatient visit or worse)*, p* is the proportion of patients who required an outpatient visit or worse, and *FNR_L_* is the false negative rate (the rate of patients classified as low risk who did require an outpatient visit or worse). This can alternatively be stated as the net increase in the proportion of low-risk individuals who avoided unnecessary follow-up using the prognostic model compared with a strategy of enrolling all patients into the telehealth service. To aid in determining whether an individual’s risk of hospital admission or worse is sufficiently large for enrollment into more rigorous clinical care, the net benefit was defined as ${TPR}_{H}-\frac{1-p}{p}*\frac{H}{1-H}*{FPR}_{H}$for a high-risk threshold *H,* where *TPR* is the true positive rate (the rate of patients classified as high risk among those who were hospitalized or died), *p* is the proportion of patients who were hospitalized or died and *FPR* is the false positive rate (the rate of patients classified as high risk among those who did not require hospitalizationand and did not die), which can alternatively be stated as the net increase in the proportion of patients who would be appropriately enrolled into more comprehensive management compared with a strategy of not offering increased follow-up to any patient.

**Supplementary Tables**

**Supplementary Table 1.** The definition of each variable used in the prognostic model, multiple imputation procedure or risk factor analysis.

| **Predictor** | **Variable description** | **Variable use** |
| --- | --- | --- |
| Age | Age of the person in years at the time of the first positive qPCR test. In the prognostic and risk factor analysis, age was modelled with a restricted cubic spline with knots placed at the 0.05, 0.35, 0.65 and 0.95 percentiles. For the multiple imputation procedure, a restricted cubic spline with six knots at the 0.05, 0.23, 0.41, 0.59, 0.77 and 0.95 percentiles was used. | Prognostic model, multiple imputation for prognostic model, risk factor analysis |
| Sex | Sex of the person as recorded in the Icelandic population register. Female = 0, Male = 1. | Prognostic model, multiple imputation for prognostic model, risk factor analysis |
| Body mass index (BMI) | Weight (kg) divided by height squared (cm) as reported during the enrollment interview. The prognostic model included BMI as a linear predictor. The risk factor analysis included BMI as a nonlinear variable using a restricted cubic spline with four knots placed at the 0.05, 0.35, 0.65 and 0.95 percentiles and the multiple imputation procedure included BMI using a restricted cubic spline with six knots placed at the 0.05, 0.23, 0.41, 0.59, 0.77 and 0.95 percentiles. | Prognostic model, multiple imputation for prognostic model, risk factor analysis |
| Hypertension,  prospective | History of hypertension reported during the enrollment interview. Absent = 0,  Present = 1. | Prognostic model, multiple imputation for prognostic model, risk factor analysis |
| Diabetes,  prospective | History of diabetes reported during the enrollment interview. Absent = 0,  Present = 1. | Prognostic model, multiple imputation for prognostic model, risk factor analysis |
| Heart disease | History of any heart disease reported during the enrollment interview. Absent = 0,  Present = 1. | Prognostic model, Multiple imputation for prognostic model, risk factor analysis |
| Pulmonary disease,  prospective | History of any pulmonary disease reported during the enrollment interview. Absent = 0,  Present = 1. | Prognostic model, multiple imputation for prognostic model, risk factor analysis |
| Current or prior malignancy,  prospective | History of current or prior malignancy reported during the enrollment interview. Absent = 0,  Present = 1. | Prognostic model, multiple imputation for prognostic model, risk factor analysis |
| Current smoking | Current cigarette smoking reported during the enrollment interview. Absent = 0,  Present = 1. | Prognostic model, multiple imputation for prognostic model, risk factor analysis |
| Prior smoking | History of prior cigarette smoking reported during the enrollment interview. Absent = 0,  Present = 1. | Risk factor analysis |
| Flu-like symptoms | Presence of any one of the following symptoms reported during the enrollment interview: fever (≥38°C), chills or rigors, non-productive cough, headache, lethargy, myalgia or anorexia. Absent = 0,  Present = 1.v | Prognostic model, multiple imputation for prognostic model |
| Upper respiratory symptoms | Presence of any one of the following symptoms reported during the enrollment interview: rhinorrhea, sore throat, dysosmia or dysgeusia. Absent = 0,  Present = 1. | Prognostic model, multiple imputation for prognostic model |
| Lower respiratory symptoms | Presence of any one of the following symptoms reported during the enrollment interview: productive cough, shortness of breath or dyspnea on exertion or at rest. Absent = 0,  Present = 1. | Prognostic model, multiple imputation for prognostic model |
| Gastrointestinal symptoms | Presence of any one of the following symptoms reported during the enrollment interview: nausea, vomiting, abdominal pain or diarrhea. Absent = 0,  Present = 1. | Prognostic model, multiple imputation for prognostic model |
| Clinical severity score | The clinical severity of the current illness as judged by the nurse or physician conducting the enrollment interview, loosely defined as: low severity (mild symptoms), moderate severity (mild dyspnea, cough or fever less than five days) and high severity (severe dyspnea, worsening cough and high or persistent fever for five days or longer).  Mild = 0, moderate or high = 1. | Prognostic model, multiple imputation for prognostic model |
| Hypertension, registry-based | ICD-10 diagnosis code I10-I16 specifying hypertensive disease or subgroups recorded in Landspitali–The National University Hospital of Iceland‘s patient registry in 2009-2020, the Register of Primary Health Care Contacts in 2004-2020 or the Register of Contacts with Medical Specialists in Private Practice in 2010-2020 **AND** a filled prescription for a medication assigned ATC code C02 (antihypertensive drugs), C07 (beta blocking agents), C08 (calcium channel blockers), or C09 (drugs acting on the renin-angiotensin system), recorded in the Prescription Medicines Register  between 395 days and 14 days before the the person´s first positive qPCR test. Absent = 0,  Present = 1. | Multiple imputation for prognostic model, risk factor analysis |
| Diabetes, registry-based | ICD-10 diagnosis code E08-13 specifying diabetes mellitus or subgroups recorded in Landspitali–The National University Hospital of Iceland‘s patient registry in 2009-2020, the Register of Primary Health Care Contacts in 2004-2020 or the Register of Contacts with Medical Specialists in Private Practice in 2010-2020 **AND** a filled prescription for a medication assigned ATC code A10 and subgroups (antidiabetic drugs) recorded in the Prescription Medicines Register between 395 days and 14 days before the person´s first positive qPCR test. Absent = 0,  Present = 1. | Multiple imputation for prognostic model, risk factor analysis |
| Heart disease, registry-based | ICD-10 diagnosis codeI20-I25 specifying ischemic heart disease, I50 specifying heart Failure, I34-I37 specifying nonrheumatic valve diseases, or I44-I49 specifying arrhythmias or subgroups recorded in Landspitali–The National University Hospital of Iceland‘s patient registry in 2009-2020, the Register of Primary Health Care Contacts in 2004-2020 or the Register of Contacts with Medical Specialists in Private Practice in 2010-2020 **AND** a filled prescription for a medication assigned ATC code B01AC (platelet aggregation inhibitors excl. heparin), C10 (lipid modifying agents), C01DA (organic nitrates), C03 (diuretics), C07 (beta blocking agents) or C09 (drugs acting on the renin-angiotensin system) recorded in the Prescription Medicines Register between 395 days and 14 days before the person´s first positive qPCR test. Absent = 0,  Present = 1. | Multiple imputation for prognostic model |
| Ischemic heart disease, registry-based | ICD-10 diagnosis code of I20-I25 specifyig ischemic heart disease or subgroups recorded in Landspitali–The National University Hospital of Iceland‘s patient registry in 2009-2020, the Register of Primary Health Care Contacts in 2004-2020 or the Register of Contacts with Medical Specialists in Private Practice in 2010-2020 **AND** a filled prescription for a medication assigned ATC code B01AC (platelet aggregation inhibitors excl. heparin), C10 (lipid modifying agents), or C01DA (organic nitrates) recorded in the Prescription Medicines Register between 395 days and 14 days before the patients’ first positive qPCR test. Absent = 0,  Present = 1. | Risk factor analysis |
| Chronic kidney disease, registry-based | Chronic kidney disease defined as the presence of two separate eGFR determinations <60 mL/min/1.73 m^2^ that were obtained at least 90 days apart, and did not occur during an episode of acute kidney injury.  The eGFR was calculated from serum creatinine values obtained from the Landspitali–The National University Hospital of Iceland‘s patient registry in 2009-2020 and the Register of Primary Health Care Contacts in 2004-2020, using the Chronic Kidney Disease Epidemiology Collaboration equation. Absent = 0,  Present = 1. | Multiple imputation for prognostic model, risk factor analysis |
| Pulmonary disease, registry-based | ICD-10 diagnosis code J40-J47 specifying chronic lower respiratory diseases or subgroups, J60-J70 specifying lung diseases due to external agents or subgroups, or J80-J84 specifying other respiratory diseases principally affecting the interstitium or subgroups recorded in Landspitali–The National University Hospital of Iceland‘s patient registry in 2009-2020, the Register of Primary Health Care Contacts in 2004-2020 or the Register of Contacts with Medical Specialists in Private Practice in 2010-2020 **AND** a filled prescription for medication assigned ATC code R03 (drugs for obstructive airway diseases) recorded in the Prescription Medicines Register between 395 days and 14 days before the person´s first positive qPCR test. Absent = 0,  Present = 1. | Multiple imputation for prognostic model |
| Chronic obstructive pulmonary disease, registry-based | ICD-10 diagnosis code J41-J44 specifying chronic lower respiratory diseases or subgroups recorded in Landspitali–The National University Hospital of Iceland‘s patient registry in 2009-2020, the Register of Primary Health Care Contacts in 2004-2020 or the Register of Contacts with Medical Specialists in Private Practice in 2010-2020 **AND** a filled prescription for a medication with ATC code R03 (drugs for obstructive airway diseases) recorded in the Drug Prescription Medicines Register  between 395 days and 14 days before the patients’ first positive qPCR test. Absent = 0,  Present = 1. | Risk factor analysis |
| Current or prior malignancy, registry-based | ICD-10 diagnosis code C00-C96 specifying neoplasms or subgroups recorded in Landspitali–The National University Hospital of Iceland‘s patient registry in 2009-2020, the Register of Primary Health Care Contacts in 2004-2020 or the Register of Contacts with Medical Specialists in Private Practice in 2010-2020. Absent = 0,  Present = 1. | Multiple imputation for prognostic model, risk factor analysis |
| Time from first case | Elapsed time in days from the day before the first diagnosed case of SARS-CoV-2 infection in Iceland (February 26, 2020) to the date of the person´s first positive qPCR test. This was modelled with a restricted cubic spline with six knots at the 0.05, 0.23, 0.41, 0.59, 0.77 and 0.95 percentiles. | Multiple imputation for prognostic model |
| Time from symptoms to diagnosis | Elapsed time in days from date of symptom onset as reported during the enrollment interview to the date of the person´s first positive qPCR test. This was modelled with a restricted cubic spline with six knots at the 0.05, 0.23, 0.41, 0.59, 0.77 and 0.95 percentiles. | Multiple imputation for prognostic model |
| Follow-up time | Elapsed time in days from the telehealth service enrollment interview to termination of telehealth follow-up or death. This was modelled with a restricted cubic spline with six knots at the 0.05, 0.23, 0.41, 0.59, 0.77 and 0.95 percentiles. | Multiple imputation for prognostic model |
| Number of primary care visits during the preceding two years | The number of visits to primary care services recorded in the Register of Primary Health Care Contacts between 730 days and 14 days before the person’s first positive qPCR test. This was modelled with a restricted cubic spline with six knots at the 0.05, 0.23, 0.41, 0.59, 0.77 and 0.95 percentiles. | Multiple imputation for prognostic model |
| Prior hospitalization | Whether or not the patient had been admitted to Landspitali–The National University Hospital of Iceland between 730 days and 14 days before the first positive qPCR test. Absent = 0,  Present = 1. | Multiple imputation for prognostic model |
| Number of filled prescriptions during the preceding 395 days (13 months) | The number of filled prescriptions n the Prescription Medicines Register between 395 days and14 days before a patients’s first positive qPCR test. This was modelled with a restricted cubic spline with six knots at the 0.05, 0.23, 0.41, 0.59, 0.77 and 0.95 percentiles. | Multiple imputation for prognostic model |
| Residence in Capital Region | The residence of the patient at the time of his/her first positive qPCR test. Equal to one if the patient resides within the Capital Region, and zero otherwise. | Multiple imputation for prognostic model |

**Supplementary Table 2.** Calibration and discrimination indices of the prognostic model for the derivation cohort, shown for each of the outcomes. The 95% bootstrapped confidence intervals are presented within parentheses.

| **Indices** | **Outpatient** | **Hospitalization** | **Intensive care unit admission or death** |
| --- | --- | --- | --- |
| C-statistic | 0.79 (0.77 to 0.83) | 0.84 (0.78 to 0.89) | 0.85 (0.78 to 0.89) |
| Cox-Snell R2 | 0.27 (0.23 to 0.33) | 0.30 (0.21 to 0.38) | 0.20 (0.12 to 0.28) |
| Calibration intercept | -0.01 (-0.16 to 0.16) | -0.01 (-0.24 to 0.15) | 0 (-0.43 to 0.31) |
| Calibration slope | 0.95 (0.85 to 1.11) | 1.09 (0.91 to 1.31) | 0.95 (0.76 to 1.17) |
| Brier score | 0.11 (0.10 to 0.12) | 0.05 (0.04 to 0.05) | 0.02 (0.01 to 0.02) |
| Brier score (rescaled) | 0.20 (0.16 to 0.26) | 0.19 (0.13 to 0.26) | 0.05 (-0.03 to 0.12) |
| E_avg_ | 0.01 (0.01 to 0.03) | 0.01 (0.01 to 0.02) | 0.01 (0.00 to 0.02) |
| E_max_ | 0.03 (0.01 to 0.10) | 0.05 (0.01 to 0.15) | 0.04 (0.01 to 0.28) |

**Supplementary Figures**

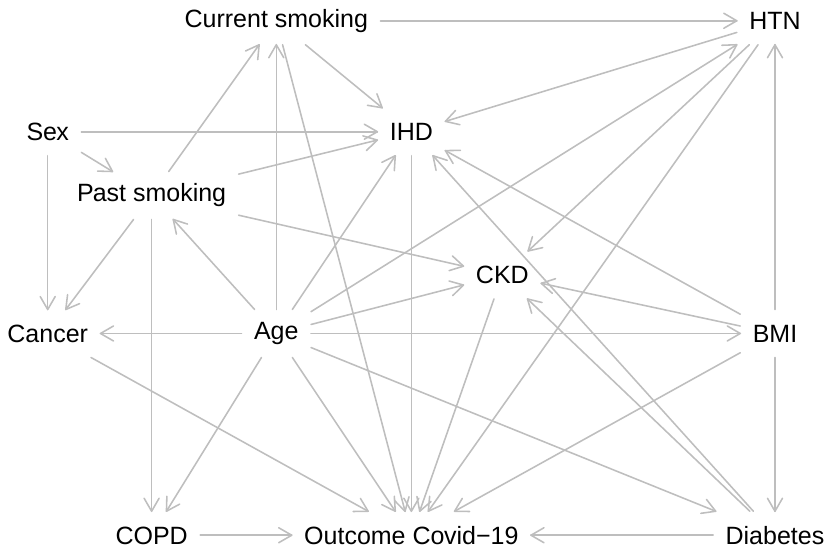

**Supplementary Figure 1.** The directed acyclic graph that describes the hypothesized relationships between the included variables and severity of COVID-19.

Abbreviations: BMI, body mass index; COPD, chronic obstructive pulmonary disease; HTN, hypertension.

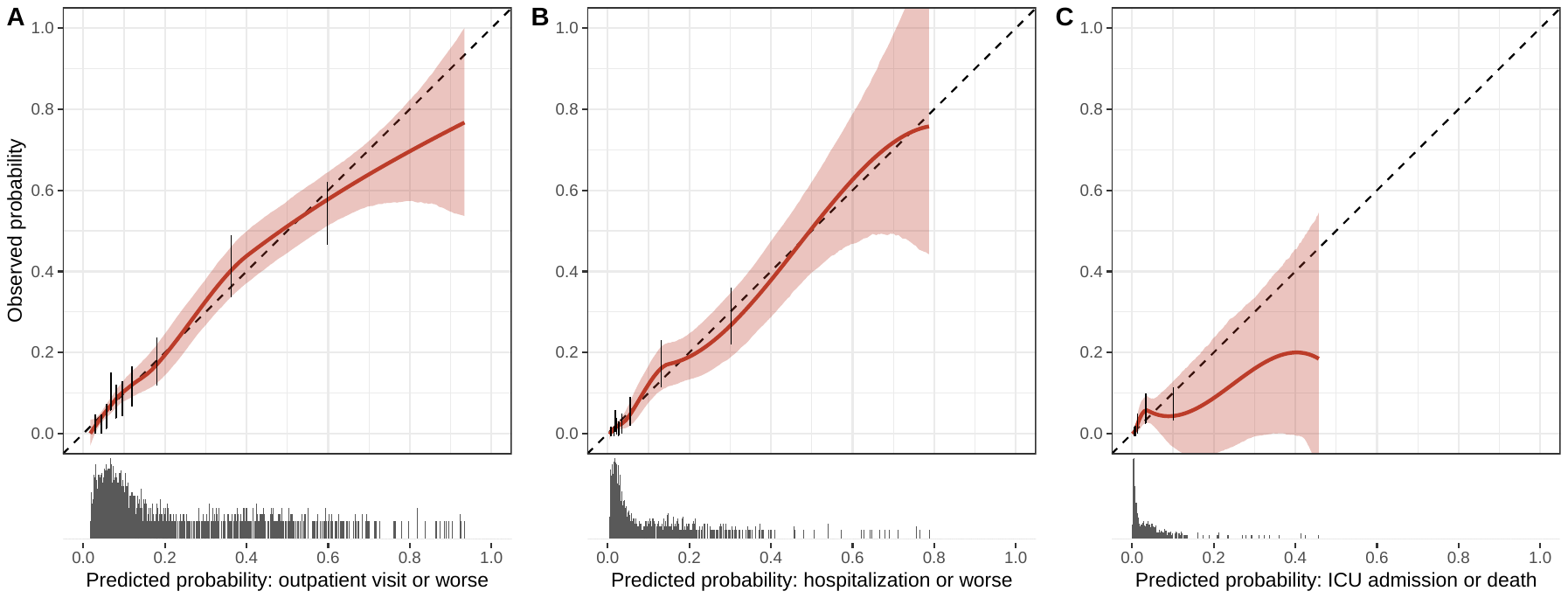

**Supplementary Figure 2.** Calibration curves of the prognostic model using the derivation cohort illustrating the relationship between the observed and predicted probability of outpatient visit or worse (Panel A), hospitalization or worse (Panel B) and admission to intensive care unit or death (Panel C). The sample distribution of predicted probabilities is demonstrated as marginal histograms. The sample is divided into 10 equally large groups and the aggregate relationship between observed and predicted probability is depicted as a black point and pointrange. The weighted scatterplot smoothing (LOWESS) relationship between the observed and predicted probabilities is shown as a red line with the shaded area representing 95% confidence intervals based on bootstrap resampling. These are compared to the dashed black line, reflecting a perfect relationship between observed and predicted probabilities.

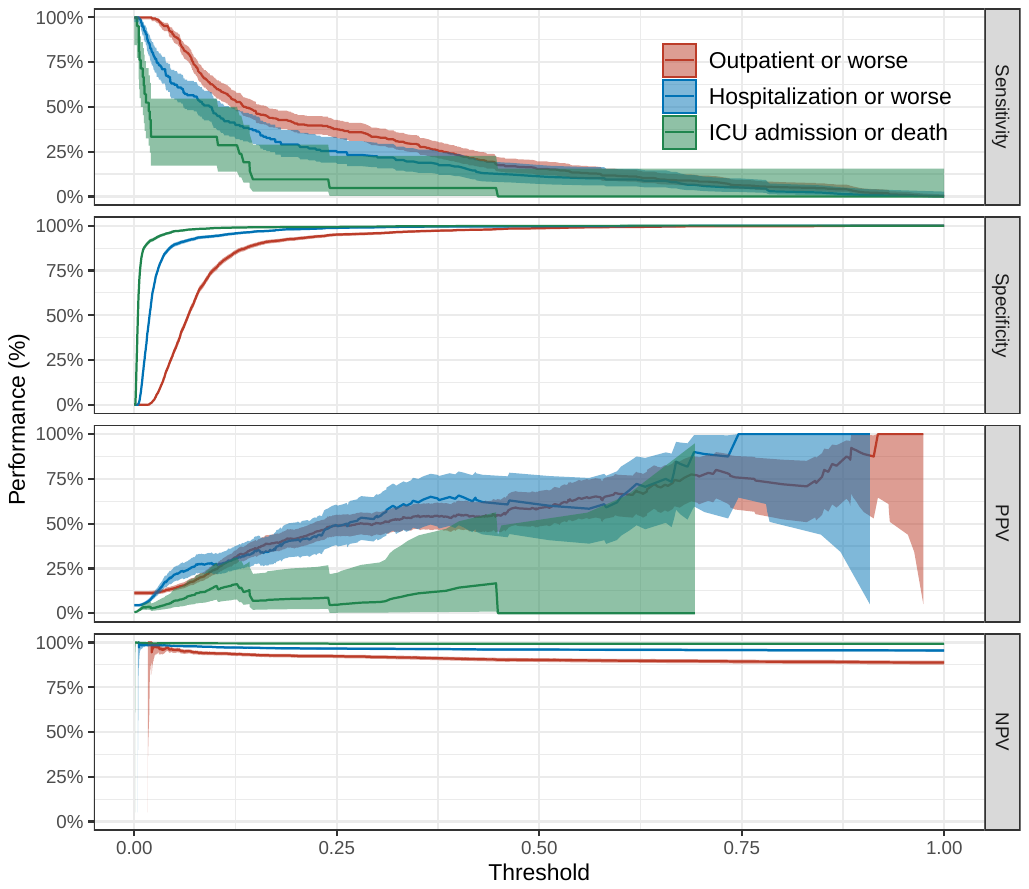

**Supplementary Figure 3.** The sensitivity, specificity, positive predictive value (PPV) and negative predictive value (NPV) of the prognostic model in the validation cohort for the three outcomes. The x-axis shows the threshold of predicted probability of the outcome at which the performance is measured.

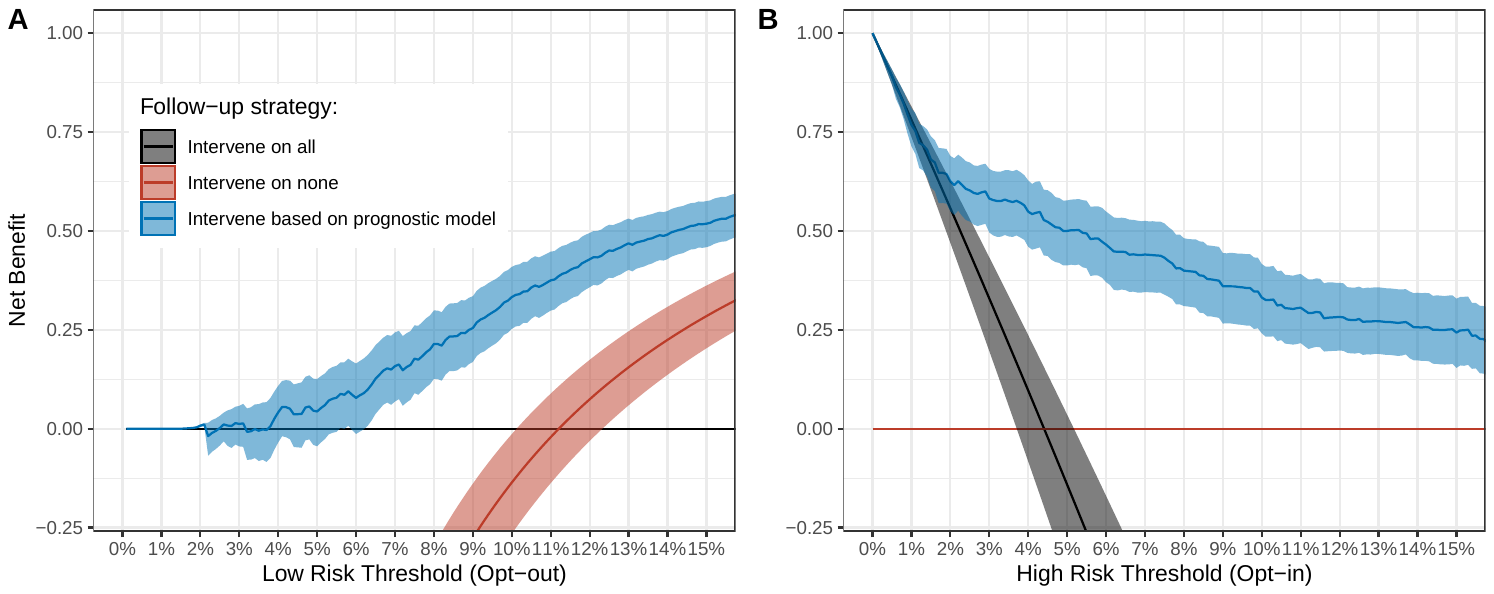

**Supplementary Figure 4.** Net benefit of the prognostic model (blue line) compared with the strategies of enrolling all individuals (black line) and enrolling none (red line). Panel A depicts the use of the prognostic model to omit low-risk patients from follow-up, in which the y-axis represents the net increase in the proportion of low-risk persons who avoided unnecessary monitoring compared with the strategy of enrolling all patients into the telehealth service. Panel B illustrates the use of the prognostic model to select high-risk patients into for rigorous follow-up or therapeutic intervention, in which the y-axis represents the net increase in the proportion of high-risk persons who were appropriately enrolled into more frequent follow-up or treatment compared with the strategy of enrolling no patients into more robust clinical care.

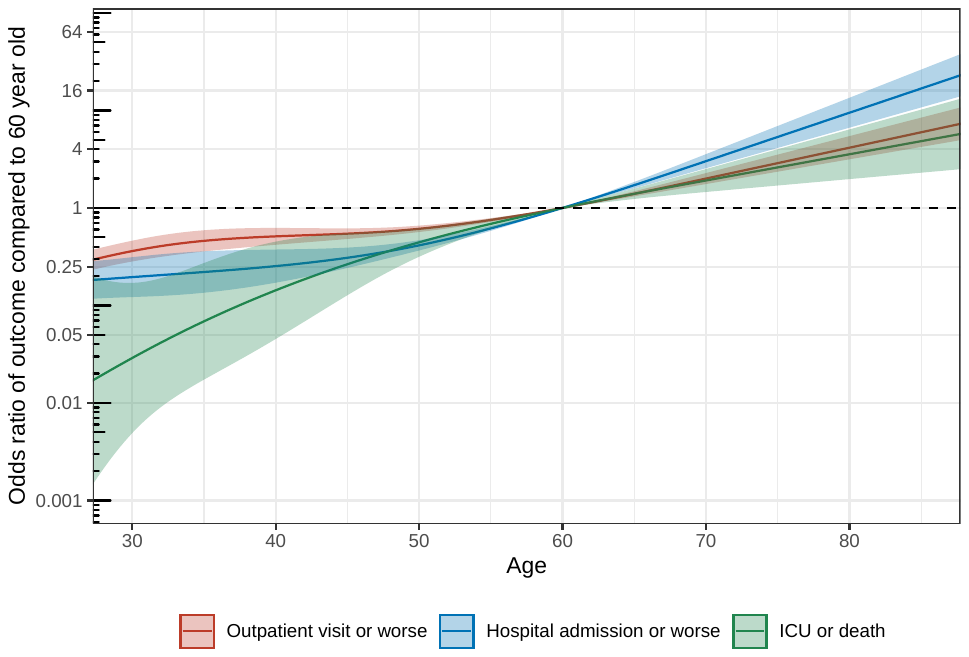

**Supplementary Figure 5.** The odds ratio of experiencing each of the three outcomes of the study when compared to an individual aged 60 years. The results are based on an analysis in which the effects of age are allowed to be non-linear using a four knot restricted cubic spline transformation. The y-axis represents the odds ratio and is shown on the logarithmic scale.

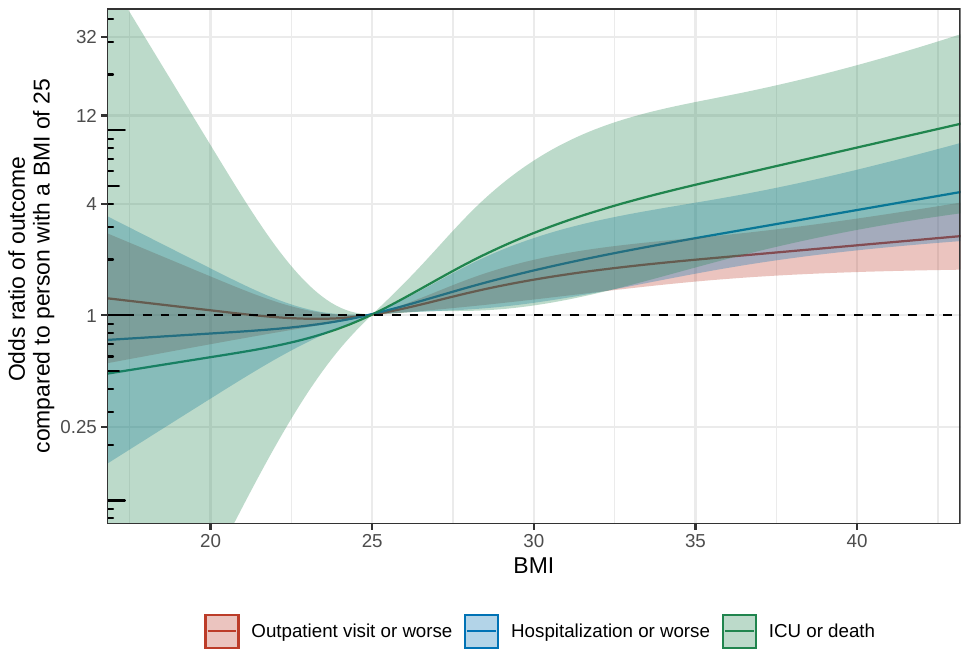

**Supplementary Figure 6.** The odds ratio of experiencing each of the three outcomes of the study when compared to an individual with a body mass index (BMI) of 25 kg/m^2^. The results are based on an analysis in which the effects of BMI are allowed to be non-linear using a four knot restricted cubic spline transformation with the relationship adjusted for a four knot restricted cubic spline transformation of age. The y-axis represents the odds ratio and is shown on the logarithmic scale.

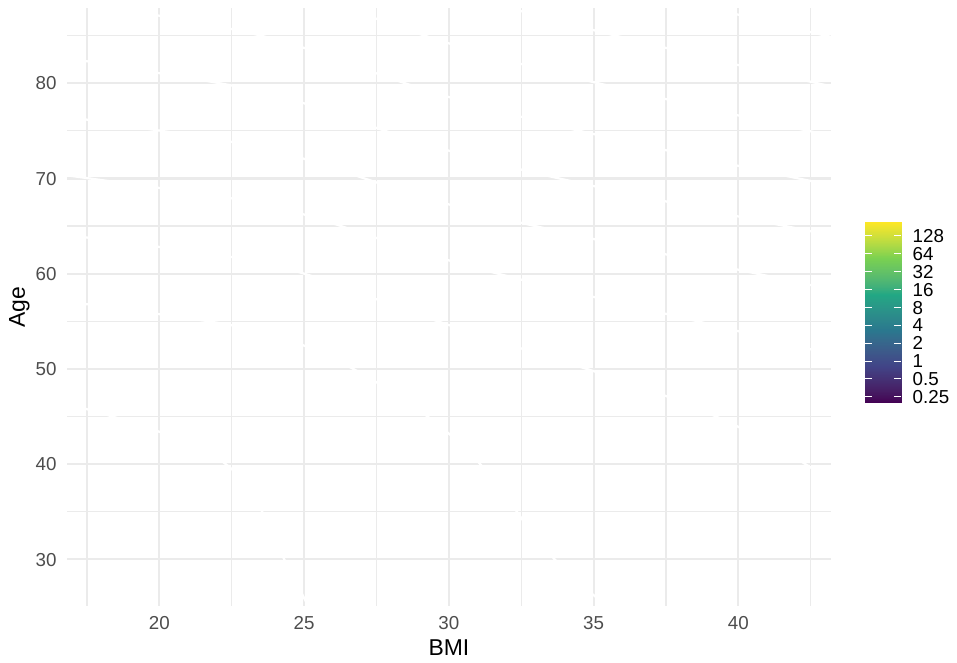

**Supplementary Figure 7.** A density plot showing the odds ratio of hospitalizationfor COVID-19 as a continuous function of age and body mass index (BMI) when compared to an individual who is 60 years of age and has a BMI of 25 kg/m^2^. The results are based on an analysis in which the effects of age and BMI are allowed to be non-linear using four knot restricted cubic spline transformations and an interaction is assumed between age and BMI. The white lines represent the odds ratios of 0.25, 0.5, 1, 2, 4, 8, 16, 32, 64 and 128.
