## Supplementary material for "Development and validation of a prognostic model for COVID-19: a population-based cohort study in Iceland": Statistical code and output: prognostic_model_20210717.html

Code for prognostic model


Code 

- Show All Code
- Hide All Code

### Code for prognostic model

###### Elias Eythorsson

###### July 17th, 2021

In the following code snippet the reader can find all the custom functions used in the analysis. The code snippet is initially hidden because the code is very long and no output is shown.

```
# This function belongs to the package CalibrationCurves and is unmodified but needs to
# be made explicit for the modified val.prob.ci.2.modified() function below.
BT.samples <- function(y, p, to.pred) {
  data.1 <- cbind.data.frame(y, p)
  
  # REPEAT TO PREVENT BT SAMPLES WITH NA'S
  repeat {
    BT.sample.rows <- sample(1:nrow(data.1), replace = T)
    BT.sample <- data.1[BT.sample.rows, ]
    loess(y ~ p, BT.sample) -> loess.BT
    predict(loess.BT, to.pred, type = "fitted") -> pred.loess
    if (!any(is.na(pred.loess))) {
      break
    }
  }
  return(pred.loess)
}

# Function modified to return the bootstrapped confidence intervals
val.prob.ci.2.modified <- function (
  p, y, logit, group, weights = rep(1, length(y)), normwt = F,
  pl = T, smooth = c("loess", "rcs", F), CL.smooth = "fill",
  CL.BT = F, lty.smooth = 1, col.smooth = "black", lwd.smooth = 1,
  nr.knots = 5, logistic.cal = F, lty.log = 1, col.log = "black",
  lwd.log = 1, xlab = "Predicted probability", ylab = "Observed proportion",
  xlim = c(-0.02, 1), ylim = c(-0.15, 1), m, g, cuts, emax.lim = c(0,1),
  legendloc = c(0.5, 0.27), statloc = c(0, 0.85),
  dostats = T, cl.level = 0.95, method.ci = "pepe", roundstats = 2,
  riskdist = "predicted", cex = 0.75, cex.leg = 0.75, connect.group = F,
  connect.smooth = T, g.group = 4, evaluate = 100, nmin = 0,
  d0lab = "0", d1lab = "1", cex.d01 = 0.7, dist.label = 0.04,
  line.bins = -0.05, dist.label2 = 0.03, cutoff, las = 1,
  length.seg = 1, y.intersp = 1, lty.ideal = 1, col.ideal = "red",
  lwd.ideal = 1, ...) {
  if (smooth[1] == F) {
    smooth <- "F"
  }
  smooth <- match.arg(smooth)
  if (!missing(p))
    if (any(!(p >= 0 | p <= 1))) {
      stop("Probabilities can not be > 1 or < 0.")
    }
  if (missing(p))
    p <- 1 / (1 + exp(-logit))
  else
    logit <- log(p / (1 - p))
  if (!all(y %in% 0:1)) {
    stop("The vector with the binary outcome can only contain the values 0 and 1.")
  }
  if (length(p) != length(y))
    stop("lengths of p or logit and y do not agree")
  names(p) <- names(y) <- names(logit) <- NULL
  if (!missing(group)) {
    if (length(group) == 1 && is.logical(group) && group)
      group <- rep("", length(y))
    if (!is.factor(group))
      group <- if (is.logical(group) || is.character(group))
        as.factor(group)
    else
      cut2(group, g = g.group)
    names(group) <- NULL
    nma <- !(is.na(p + y + weights) | is.na(group))
    ng <- length(levels(group))
  }
  else {
    nma <- !is.na(p + y + weights)
    ng <- 0
  }
  logit <- logit[nma]
  y <- y[nma]
  p <- p[nma]
  y <- y[order(p)]
  logit <- logit[order(p)]
  p <- p[order(p)]
  if (length(p) > 5000 & smooth == "loess") {
    warning("Number of observations > 5000, RCS is recommended.",
            immediate. = T)
  }
  if (length(p) > 1000 & CL.BT == T) {
    warning("Number of observations is > 1000, this could take a while...",
            immediate. = T)
  }
  if (length(unique(p)) == 1) {
    P <- mean(y)
    Intc <- log(P / (1 - P))
    n <- length(y)
    D <- -1 / n
    L01 <- -2 * sum(y * logit - log(1 + exp(logit)), na.rm = T)
    L.cal <- -2 * sum(y * Intc - log(1 + exp(Intc)), na.rm = T)
    U.chisq <- L01 - L.cal
    U.p <- 1 - pchisq(U.chisq, 1)
    U <- (U.chisq - 1) / n
    Q <- D - U
    stats <- c(0, 0.5, 0, D, 0, 1, U, U.chisq, U.p, Q, mean((y - p[1]) ^ 2), Intc, 0, rep(abs(p[1] - P), 2))
    names(stats) <- c("Dxy", "C (ROC)", "R2", "D", "D:Chi-sq", "D:p", "U",
                      "U:Chi-sq", "U:p", "Q", "Brier", "Intercept", "Slope",
                      "Emax", "Eavg", "ECI")
  }
  i <- !is.infinite(logit)
  nm <- sum(!i)
  if (nm > 0)
    warning(paste(
      nm,
      "observations deleted from logistic calibration due to probs. of 0 or 1"
    ))
  i.2 <- i
  f.or <- lrm(y[i] ~ logit[i])
  f <- lrm.fit(logit[i], y[i])
  cl.slope <- confint(f, level = cl.level)[2, ]
  f2 <- lrm.fit(offset = logit[i], y = y[i])
  if (f2$fail) {
    warning(
      "The lrm function did not converge when computing the calibration intercept!",
      immediate. = T
    )
    f2 <- list()
    f2$coef <- NA
    cl.interc <- rep(NA, 2)
  }
  else {
    cl.interc <- confint(f2, level = cl.level)
  }
  stats <- f$stats
  n <- stats["Obs"]
  predprob <- seq(emax.lim[1], emax.lim[2], by = 5e-04)
  lt <- f$coef[1] + f$coef[2] * log(predprob / (1 - predprob))
  calp <- 1 / (1 + exp(-lt))
  emax <- max(abs(predprob - calp))
  if (pl) {
    plot(0.5, 0.5, xlim = xlim, ylim = ylim, type = "n", xlab = xlab, ylab = ylab, las = las, ...)
    clip(0, 1, 0, 1)
    abline(0, 1, lty = lty.ideal, col = col.ideal, lwd = lwd.ideal)
    do.call("clip", as.list(par()$usr))
    lt <- lty.ideal
    lw.d <- lwd.ideal
    all.col <- col.ideal
    leg <- "Ideal"
    marks <- -1
    if (logistic.cal) {
      lt <- c(lt, lty.log)
      lw.d <- c(lw.d, lwd.log)
      all.col <- c(all.col, col.log)
      leg <- c(leg, "Logistic calibration")
      marks <- c(marks, -1)
    }
    if (smooth != "F") {
      all.col <- c(all.col, col.smooth)
    }
    if (smooth == "loess") {
      Sm <- loess(y ~ p, degree = 2)
      Sm <- data.frame(Sm$x, Sm$fitted)
      Sm.01 <- Sm
      if (connect.smooth == T & CL.smooth != "fill") {
        clip(0, 1, 0, 1)
        lines(Sm,
              lty = lty.smooth,
              lwd = lwd.smooth,
              col = col.smooth)
        do.call("clip", as.list(par()$usr))
        lt <- c(lt, lty.smooth)
        lw.d <- c(lw.d, lwd.smooth)
        marks <- c(marks, -1)
      }
      else if (connect.smooth == F & CL.smooth != "fill") {
        clip(0, 1, 0, 1)
        points(Sm, col = col.smooth)
        do.call("clip", as.list(par()$usr))
        lt <- c(lt, 0)
        lw.d <- c(lw.d, 1)
        marks <- c(marks, 1)
      }
      if (CL.smooth == T | CL.smooth == "fill") {
        to.pred <- seq(min(p), max(p), length = 500)
        if (CL.BT == T) {
          cat("Bootstrap samples are being generated.\n\n\n")
          res.BT <- replicate(2000, BT.samples(y, p,
                                               to.pred))
          CL.BT <- apply(res.BT, 1, quantile, c(0.025,
                                                0.975))
          colnames(CL.BT) <- to.pred
          if (CL.smooth == "fill") {
            clip(0, 1, 0, 1)
            polygon(
              x = c(to.pred, rev(to.pred)),
              y = c(CL.BT[2, ], rev(CL.BT[1, ])),
              col = rgb(177, 177,
                        177, 177, maxColorValue = 255),
              border = NA
            )
            if (connect.smooth == T) {
              lines(Sm,
                    lty = lty.smooth,
                    lwd = lwd.smooth,
                    col = col.smooth)
              lt <- c(lt, lty.smooth)
              lw.d <- c(lw.d, lwd.smooth)
              marks <- c(marks, -1)
            }
            else if (connect.smooth == F) {
              points(Sm, col = col.smooth)
              lt <- c(lt, 0)
              lw.d <- c(lw.d, 1)
              marks <- c(marks, 1)
            }
            do.call("clip", as.list(par()$usr))
            leg <- c(leg, "Flexible calibration (Loess)")
          }
          else {
            clip(0, 1, 0, 1)
            lines(
              to.pred,
              CL.BT[1, ],
              lty = 2,
              lwd = 1,
              col = col.smooth
            )
            clip(0, 1, 0, 1)
            lines(
              to.pred,
              CL.BT[2, ],
              lty = 2,
              lwd = 1,
              col = col.smooth
            )
            do.call("clip", as.list(par()$usr))
            leg <- c(leg, "Flexible calibration (Loess)",
                     "CL flexible")
            lt <- c(lt, 2)
            lw.d <- c(lw.d, 1)
            all.col <- c(all.col, col.smooth)
            marks <- c(marks, -1)
          }
        }
        else {
          Sm.0 <- loess(y ~ p, degree = 2)
          cl.loess <- predict(Sm.0, type = "fitted",
                              se = T)
          clip(0, 1, 0, 1)
          if (CL.smooth == "fill") {
            polygon(
              x = c(Sm.0$x, rev(Sm.0$x)),
              y = c(
                cl.loess$fit +
                  cl.loess$se.fit * 1.96,
                rev(cl.loess$fit -
                      cl.loess$se.fit * 1.96)
              ),
              col = rgb(177,
                        177, 177, 177, maxColorValue = 255),
              border = NA
            )
            if (connect.smooth == T) {
              lines(Sm,
                    lty = lty.smooth,
                    lwd = lwd.smooth,
                    col = col.smooth)
              lt <- c(lt, lty.smooth)
              lw.d <- c(lw.d, lwd.smooth)
              marks <- c(marks, -1)
            }
            else if (connect.smooth == F) {
              points(Sm, col = col.smooth)
              lt <- c(lt, 0)
              lw.d <- c(lw.d, 1)
              marks <- c(marks, 1)
            }
            do.call("clip", as.list(par()$usr))
            leg <- c(leg, "Flexible calibration (Loess)")
          }
          else {
            lines(
              Sm.0$x,
              cl.loess$fit + cl.loess$se.fit *
                1.96,
              lty = 2,
              lwd = 1,
              col = col.smooth
            )
            lines(
              Sm.0$x,
              cl.loess$fit - cl.loess$se.fit *
                1.96,
              lty = 2,
              lwd = 1,
              col = col.smooth
            )
            do.call("clip", as.list(par()$usr))
            leg <- c(leg, "Flexible calibration (Loess)",
                     "CL flexible")
            lt <- c(lt, 2)
            lw.d <- c(lw.d, 1)
            all.col <- c(all.col, col.smooth)
            marks <- c(marks,-1)
          }
        }
      }
      else {
        leg <- c(leg, "Flexible calibration (Loess)")
      }
      cal.smooth <- approx(Sm.01, xout = p)$y
      eavg <- mean(abs(p - cal.smooth))
      ECI <- mean((p - cal.smooth) ^ 2) * 100
    }
    if (smooth == "rcs") {
      par(lwd = lwd.smooth,
          bty = "n",
          col = col.smooth)
      if (!is.numeric(nr.knots)) {
        stop("Nr.knots must be numeric.")
      }
      if (nr.knots == 5) {
        tryCatch(
          rcspline.plot(
            p,
            y,
            model = "logistic",
            nk = 5,
            show = "prob",
            statloc = "none",
            add = T,
            showknots = F,
            xrange = c(min(na.omit(p)),
                       max(na.omit(p))),
            lty = lty.smooth
          ),
          error = function(e) {
            warning("The number of knots led to estimation problems, nk will be set to 4.",
                    immediate. = T)
            tryCatch(
              rcspline.plot(
                p,
                y,
                model = "logistic",
                nk = 4,
                show = "prob",
                statloc = "none",
                add = T,
                showknots = F,
                xrange = c(min(na.omit(p)),
                           max(na.omit(p))),
                lty = lty.smooth
              ),
              error = function(e) {
                warning("Nk 4 also led to estimation problems, nk will be set to 3.",
                        immediate. = T)
                rcspline.plot(
                  p,
                  y,
                  model = "logistic",
                  nk = 3,
                  show = "prob",
                  statloc = "none",
                  add = T,
                  showknots = F,
                  xrange = c(min(na.omit(p)),
                             max(na.omit(p))),
                  lty = lty.smooth
                )
              }
            )
          }
        )
      }
      else if (nr.knots == 4) {
        tryCatch(
          rcspline.plot(
            p,
            y,
            model = "logistic",
            nk = 4,
            show = "prob",
            statloc = "none",
            add = T,
            showknots = F,
            xrange = c(min(na.omit(p)),
                       max(na.omit(p))),
            lty = lty.smooth
          ),
          error = function(e) {
            warning("The number of knots led to estimation problems, nk will be set to 3.",
                    immediate. = T)
            rcspline.plot(
              p,
              y,
              model = "logistic",
              nk = 3,
              show = "prob",
              statloc = "none",
              add = T,
              showknots = F,
              xrange = c(min(na.omit(p)),
                         max(na.omit(p))),
              lty = lty.smooth
            )
          }
        )
      }
      else if (nr.knots == 3) {
        tryCatch(
          rcspline.plot(
            p,
            y,
            model = "logistic",
            nk = 3,
            show = "prob",
            statloc = "none",
            add = T,
            showknots = F,
            xrange = c(min(na.omit(p)),
                       max(na.omit(p))),
            lty = lty.smooth
          ),
          error = function(e) {
            stop("Nk = 3 led to estimation problems.")
          }
        )
      }
      else {
        stop(paste(
          "Number of knots = ",
          nr.knots,
          sep = "",
          ", only 5 >= nk >=3 is allowed."
        ))
      }
      par(lwd = 1,
          bty = "o",
          col = "black")
      leg <- c(leg, "Flexible calibration (RCS)", "CL flexible")
      lt <- c(lt, lty.smooth, 2)
      lw.d <- c(lw.d, rep(lwd.smooth, 2))
      all.col <- c(all.col, col.smooth)
      marks <- c(marks, -1, -1)
    }
    if (!missing(m) | !missing(g) | !missing(cuts)) {
      if (!missing(m))
        q <- cut2(p,
                  m = m,
                  levels.mean = T,
                  digits = 7)
      else if (!missing(g))
        q <- cut2(p,
                  g = g,
                  levels.mean = T,
                  digits = 7)
      else if (!missing(cuts))
        q <- cut2(p,
                  cuts = cuts,
                  levels.mean = T,
                  digits = 7)
      means <- as.single(levels(q))
      prop <- tapply(y, q, function(x)
        mean(x, na.rm = T))
      points(means, prop, pch = 2, cex = 1)
      ng <- tapply(y, q, length)
      og <- tapply(y, q, sum)
      ob <- og / ng
      se.ob <- sqrt(ob * (1 - ob) / ng)
      g <- length(as.single(levels(q)))
      for (i in 1:g)
        lines(c(means[i], means[i]), c(prop[i],
                                       min(1, prop[i] + 1.96 * se.ob[i])), type = "l")
      for (i in 1:g)
        lines(c(means[i], means[i]), c(prop[i],
                                       max(0, prop[i] - 1.96 * se.ob[i])), type = "l")
      if (connect.group) {
        lines(means, prop)
        lt <- c(lt, 1)
        lw.d <- c(lw.d, 1)
      }
      else {
        lt <- c(lt, 0)
        lw.d <- c(lw.d, 0)
      }
      leg <- c(leg, "Grouped observations")
      all.col <- c(all.col, col.smooth)
      marks <- c(marks, 2)
    }
  }
  lr <- stats["Model L.R."]
  p.lr <- stats["P"]
  D <- (lr - 1) / n
  L01 <- -2 * sum(y * logit - logb(1 + exp(logit)), na.rm = TRUE)
  U.chisq <- L01 - f$deviance[2]
  p.U <- 1 - pchisq(U.chisq, 2)
  U <- (U.chisq - 2) / n
  Q <- D - U
  Dxy <- stats["Dxy"]
  C <- stats["C"]
  R2 <- stats["R2"]
  B <- sum((p - y) ^ 2) / n
  Bmax <-
    mean(y) * (1 - mean(y)) ^ 2 + (1 - mean(y)) * mean(y) ^ 2
  Bscaled <- 1 - B / Bmax
  stats <- c(Dxy, C, R2, D, lr, p.lr, U, U.chisq, p.U, Q, B, f2$coef[1], f$coef[2], emax, Bscaled)
  names(stats) <- c(
    "Dxy", "C (ROC)", "R2", "D", "D:Chi-sq", "D:p", "U", "U:Chi-sq", "U:p", "Q", "Brier",
    "Intercept", "Slope", "Emax", "Brier scaled")
  if (smooth == "loess")
    stats <- c(stats, c(Eavg = eavg), c(ECI = ECI))
  if (!missing(cutoff)) {
    arrows(
      x0 = cutoff,
      y0 = 0.1,
      x1 = cutoff,
      y1 = -0.025,
      length = 0.15
    )
  }
  if (pl) {
    if (min(p) > plogis(-7) | max(p) < plogis(7)) {
      lrm.fit.1 <- lrm(y[i.2] ~ qlogis(p[i.2]))
      if (logistic.cal)
        lines(
          p[i.2],
          plogis(lrm.fit.1$linear.predictors),
          lwd = lwd.log,
          lty = lty.log,
          col = col.log
        )
    }
    else {
      logit <- seq(-7, 7, length = 200)
      prob <- 1 / (1 + exp(-logit))
      pred.prob <- f$coef[1] + f$coef[2] * logit
      pred.prob <- 1 / (1 + exp(-pred.prob))
      if (logistic.cal)
        lines(prob,
              pred.prob,
              lty = lty.log,
              lwd = lwd.log,
              col = col.log)
    }
    lp <- legendloc
    if (!is.logical(lp)) {
      if (!is.list(lp))
        lp <- list(x = lp[1], y = lp[2])
      legend(
        lp,
        leg,
        lty = lt,
        pch = marks,
        cex = cex.leg,
        bty = "n",
        lwd = lw.d,
        col = all.col,
        y.intersp = y.intersp
      )
    }
    if (is.character(riskdist)) {
      if (riskdist == "calibrated") {
        x <- f$coef[1] + f$coef[2] * log(p / (1 - p))
        x <- 1 / (1 + exp(-x))
        x[p == 0] <- 0
        x[p == 1] <- 1
      }
      else
        x <- p
      bins <- seq(0, min(1, max(xlim)), length = 101)
      x <- x[x >= 0 & x <= 1]
      f0 <- table(cut(x[y == 0], bins))
      f1 <- table(cut(x[y == 1], bins))
      j0 <- f0 > 0
      j1 <- f1 > 0
      bins0 <- (bins[-101])[j0]
      bins1 <- (bins[-101])[j1]
      f0 <- f0[j0]
      f1 <- f1[j1]
      maxf <- max(f0, f1)
      f0 <- (0.1 * f0) / maxf
      f1 <- (0.1 * f1) / maxf
      segments(bins1, line.bins, bins1, length.seg * f1 +
                 line.bins)
      segments(bins0, line.bins, bins0, length.seg * -f0 +
                 line.bins)
      lines(c(min(bins0, bins1) - 0.01, max(bins0, bins1) +
                0.01), c(line.bins, line.bins))
      text(max(bins0, bins1) + dist.label,
           line.bins +
             dist.label2,
           d1lab,
           cex = cex.d01)
      text(max(bins0, bins1) + dist.label,
           line.bins -
             dist.label2,
           d0lab,
           cex = cex.d01)
    }
  }
  if (dostats == T) {
    cat(
      paste(
        "\n\n A ",
        cl.level * 100,
        "% confidence interval is given for the calibration intercept, calibration slope and c-statistic. \n\n",
        sep = ""
      )
    )
  }
  stats
  return(CL.BT)
}

# Generic logistic function
expit <- function(xx) exp(xx)/ (1+exp(xx))

nejm_palette <- c(
  "#BC3C29FF",
  "#0072B5FF",
  "#E18727FF",
  "#20854EFF",
  "#7876B1FF",
  "#6F99ADFF",
  "#FFDC91FF"
)

# The original function was courtesy of Darren L Dahly https://darrendahly.github.io/post/homr/
# but was modified to use val.prob.ci.2()
boot_val2 <- function(data, predicted, outcome, ...){
  out <- list()
  for(i in 1:60){
    df <- sample_n(data, nrow(data), replace = TRUE)
    val <-  val.prob.ci.2(
      p = df[, predicted],
      y = df[, outcome],
      statloc = FALSE,
      dostats = FALSE
    )
    out[[i]] <- val
  }
  return(out)
}

# The original function was courtesy of Darren L Dahly https://darrendahly.github.io/post/homr/
# Slightly modified to fit with the output of boot_val2

rescale_brier <- function(x, p, ...){ 
  format(round(1 - (x / (mean(p) * (1 - mean(p)))), digits = 2), nsmall = 2)
}

calc_ci <- function(metric, boot_vals, n, scale_B) {
  if (metric == "C") {
    metric <- "Dxy"
    x <- unlist(map(boot_vals, `[`, c(metric)))
    x <- (x + 1)/2
  } else {
    x <- unlist(map(boot_vals, `[`, c(metric)))
  }
  if (metric == "B") {
    x <- as.numeric(rescale_brier(x, scale_B))
  }
  paste0("(", round(quantile(x, 0.025), n), " to ",
         round(quantile(x, 0.975), n), ")")
}

# Original courtesy of Karendeep Singh https://twitter.com/kdpsinghlab
# Slightly modified to fit our purposes. 
expand_preds <- function (.data, threshold, inc = NULL) {
  threshold <- unique(threshold)
  nth <- length(threshold)
  n_data <- nrow(.data)
  if (!is.null(inc))
    .data <- dplyr::select(.data, all_of(inc))
  .data <- .data[rep(1:nrow(.data), times = nth),]
  .data$.threshold <- rep(threshold, each = n_data)
  .data
}

recode_data <- function (obs, prob, threshold) {
  lvl <- levels(obs)
  if (getOption("yardstick.event_first", default = TRUE)) {
    pred <- ifelse(prob >= threshold, lvl[1], lvl[2])
  }
  else {
    pred <- ifelse(prob >= threshold, lvl[2], lvl[1])
  }
  factor(pred, levels = lvl)
}

two_class = function (...) {
  mets <- yardstick::metric_set(yardstick::sens,
                                yardstick::spec,
                                yardstick::ppv,
                                yardstick::npv)
  mets(...)
}

threshperf <- function(df, outcome, prediction) {
  
  thresholds = unique(c(0,sort(unique(df[[prediction]])), 1))
  
  df <- dplyr::select(df, dplyr::all_of(c(outcome, prediction)))
  
  df <- na.omit(df)
  
  df_orig <- df
  
  # IMPORTANT because order of levels matters to yardstick
  if (getOption('yardstick.event_first', default = TRUE)) {
    df[[outcome]] <- factor(df[[outcome]], levels = c(1,0))
  } else {
    df[[outcome]] <- factor(df[[outcome]], levels = c(0,1))
  }
  
  
  df <- df %>%
    expand_preds(threshold = thresholds,
                 inc = c(outcome, prediction)) %>%
    dplyr::mutate(alt_pred = recode_data(df[[outcome]], df[[prediction]], .threshold))
  
  
  df <- df %>% dplyr::group_by(.threshold)
  
  df_metrics <- df %>%
    two_class(truth = get(outcome), estimate = alt_pred)
  
  df_metrics <-
    df_metrics %>%
    dplyr::group_by(.threshold) %>%
    dplyr::mutate(denom =
                    dplyr::case_when(
                      .metric == 'sens' ~ sum(df_orig[[outcome]] == 1),
                      .metric == 'spec' ~ sum(df_orig[[outcome]] == 0),
                      .metric == 'ppv' ~ sum(df_orig[[prediction]] >= .threshold),
                      .metric == 'npv' ~ sum(df_orig[[prediction]] < .threshold),
                    )) %>%
    dplyr::ungroup() %>%
    dplyr::mutate(numer = round(.estimate * denom)) %>%
    na.omit()
  
  df_ci = Hmisc::binconf(x = df_metrics$numer, n = df_metrics$denom,
                         alpha = 0.05, method = 'wilson') %>%
    dplyr::as_tibble() %>%
    dplyr::rename(ll = Lower, ul = Upper) %>%
    dplyr::mutate_at(dplyr::vars(ul, ll), . %>% scales::oob_squish(range = c(0,1)))
  
  df_metrics = dplyr::bind_cols(df_metrics, df_ci)
  
  data.frame(df_metrics, check.names = FALSE, stringsAsFactors = FALSE)
}
```

### 1 Libraries

```
library(readxl)
library(tidyverse)
library(lubridate)
library(rms)
library(cowplot)
library(tableone)
library(rmda)
library(CalibrationCurves)
library(dagitty)
library(skimr)

sessionInfo()
```

```
## R version 3.6.3 (2020-02-29)
## Platform: x86_64-apple-darwin15.6.0 (64-bit)
## Running under: macOS Catalina 10.15.7
## 
## Matrix products: default
## BLAS:   /Library/Frameworks/R.framework/Versions/3.6/Resources/lib/libRblas.0.dylib
## LAPACK: /Library/Frameworks/R.framework/Versions/3.6/Resources/lib/libRlapack.dylib
## 
## locale:
## [1] en_US.UTF-8/en_US.UTF-8/en_US.UTF-8/C/en_US.UTF-8/en_US.UTF-8
## 
## attached base packages:
## [1] stats     graphics  grDevices utils     datasets  methods   base     
## 
## other attached packages:
##  [1] skimr_2.1.3             dagitty_0.3-1           CalibrationCurves_0.1.2
##  [4] rmda_1.6                tableone_0.12.0         cowplot_1.0.0          
##  [7] rms_5.1-4               SparseM_1.78            Hmisc_4.4-0            
## [10] Formula_1.2-3           survival_3.1-12         lattice_0.20-41        
## [13] lubridate_1.7.8         forcats_0.5.0           stringr_1.4.0          
## [16] dplyr_1.0.2             purrr_0.3.4             readr_1.3.1            
## [19] tidyr_1.1.2             tibble_3.0.3            ggplot2_3.3.2          
## [22] tidyverse_1.3.0         readxl_1.3.1           
## 
## loaded via a namespace (and not attached):
##  [1] TH.data_1.0-10       colorspace_1.4-1     ellipsis_0.3.0      
##  [4] class_7.3-16         htmlTable_1.13.3     base64enc_0.1-3     
##  [7] fs_1.4.1             rstudioapi_0.11      MatrixModels_0.4-1  
## [10] prodlim_2019.11.13   fansi_0.4.1          mvtnorm_1.1-0       
## [13] xml2_1.3.1           codetools_0.2-16     splines_3.6.3       
## [16] knitr_1.28           jsonlite_1.6.1       pROC_1.16.2         
## [19] caret_6.0-86         broom_0.7.0          cluster_2.1.0       
## [22] dbplyr_1.4.2         png_0.1-7            compiler_3.6.3      
## [25] httr_1.4.1           backports_1.1.6      assertthat_0.2.1    
## [28] Matrix_1.2-18        survey_4.0           cli_2.0.2           
## [31] acepack_1.4.1        htmltools_0.5.0      quantreg_5.55       
## [34] tools_3.6.3          gtable_0.3.0         glue_1.4.0          
## [37] reshape2_1.4.4       V8_3.0.2             Rcpp_1.0.4.6        
## [40] cellranger_1.1.0     vctrs_0.3.4          nlme_3.1-145        
## [43] iterators_1.0.12     timeDate_3043.102    xfun_0.24           
## [46] gower_0.2.2          rvest_0.3.5          lifecycle_0.2.0     
## [49] polspline_1.1.17     MASS_7.3-51.5        zoo_1.8-7           
## [52] scales_1.1.1         ipred_0.9-9          hms_0.5.3           
## [55] sandwich_2.5-1       RColorBrewer_1.1-2   curl_4.3            
## [58] yaml_2.2.1           gridExtra_2.3        pander_0.6.3        
## [61] rpart_4.1-15         reshape_0.8.8        latticeExtra_0.6-29 
## [64] stringi_1.4.6        highr_0.8            foreach_1.5.0       
## [67] checkmate_2.0.0      boot_1.3-24          lava_1.6.7          
## [70] repr_1.1.3           rlang_0.4.7          pkgconfig_2.0.3     
## [73] evaluate_0.14        recipes_0.1.13       htmlwidgets_1.5.1   
## [76] tidyselect_1.1.0     plyr_1.8.6           magrittr_1.5        
## [79] R6_2.4.1             generics_0.0.2       multcomp_1.4-13     
## [82] DBI_1.1.0            pillar_1.4.3         haven_2.3.1         
## [85] foreign_0.8-76       withr_2.1.2          nnet_7.3-13         
## [88] modelr_0.1.6         crayon_1.3.4         rmarkdown_2.9       
## [91] jpeg_0.1-8.1         grid_3.6.3           data.table_1.12.8   
## [94] ModelMetrics_1.2.2.2 reprex_0.3.0         digest_0.6.25       
## [97] stats4_3.6.3         munsell_0.5.0        mitools_2.4
```

### 2 Import data

```
df_outcome <- read_csv("_data/df_outcome_prognostic.csv")
df_phone <- read_csv("_data/df_phone_prognostic.csv")
df_tests <- read_csv("_data/df_tests_prognostic.csv")
dag_outcome <- downloadGraph("dagitty.net/mY9jiTj")
```

Only one variable needs to be created/modified. This is turning the outcome into a factor.

```
df_outcome <- df_outcome %>%
  mutate(outcome_fct = factor(outcome, levels = c(0, 1, 2, 3)))
```

Here we split df\_outcome into the derivation and validation cohorts

```
df_derivation <- df_outcome %>%
  filter(
    cohort == "Derivation",
    excluded == "Included"
  )

df_validation <- df_outcome %>%
  filter(
    cohort == "Validation",
    excluded == "Included"
  )
```

### 3 Data and missing data

First we summarize all the included data, showing the full distribution of all variables that were collected regardless of whether these were used in the analysis. We show these

```
skim(
  df_outcome %>%
    filter(excluded == "Included") %>%
    mutate_if(~ is.numeric(.) && all(unique(.) %in% c(0, 1, NA)), as.logical) %>%
    group_by(cohort)
  )
```

Data summary

|  |  |
| --- | --- |
| Name | %>%(…) |
| Number of rows | 4756 |
| Number of columns | 109 |
| \_\_\_\_\_\_\_\_\_\_\_\_\_\_\_\_\_\_\_\_\_\_\_ |  |
| Column type frequency: |  |
| character | 5 |
| Date | 3 |
| factor | 1 |
| logical | 82 |
| numeric | 17 |
| \_\_\_\_\_\_\_\_\_\_\_\_\_\_\_\_\_\_\_\_\_\_\_\_ |  |
| Group variables | cohort |

**Variable type: character**

| skim\_variable | cohort | n\_missing | complete\_rate | min | max | empty | n\_unique | whitespace |
| --- | --- | --- | --- | --- | --- | --- | --- | --- |
| kt | Derivation | 0 | 1 | 10 | 10 | 0 | 1625 | 0 |
| kt | Validation | 0 | 1 | 10 | 10 | 0 | 3131 | 0 |
| excluded | Derivation | 0 | 1 | 8 | 8 | 0 | 1 | 0 |
| excluded | Validation | 0 | 1 | 8 | 8 | 0 | 1 | 0 |
| zip | Derivation | 0 | 1 | 3 | 11 | 0 | 75 | 0 |
| zip | Validation | 0 | 1 | 3 | 11 | 0 | 100 | 0 |
| town | Derivation | 0 | 1 | 4 | 19 | 0 | 49 | 0 |
| town | Validation | 0 | 1 | 3 | 22 | 0 | 59 | 0 |
| distr | Derivation | 0 | 1 | 8 | 16 | 0 | 8 | 0 |
| distr | Validation | 0 | 1 | 8 | 16 | 0 | 8 | 0 |

**Variable type: Date**

| skim\_variable | cohort | n\_missing | complete\_rate | min | max | median | n\_unique |
| --- | --- | --- | --- | --- | --- | --- | --- |
| date\_pcr | Derivation | 0 | 1.00 | 2020-02-28 | 2020-04-30 | 2020-03-26 | 59 |
| date\_pcr | Validation | 0 | 1.00 | 2020-05-05 | 2020-12-31 | 2020-10-12 | 179 |
| date\_symptoms | Derivation | 54 | 0.97 | 2020-02-15 | 2020-04-29 | 2020-03-20 | 67 |
| date\_symptoms | Validation | 146 | 0.95 | 2020-03-03 | 2020-12-31 | 2020-10-09 | 184 |
| date\_discharge | Derivation | 11 | 0.99 | 2020-03-18 | 2020-05-26 | 2020-04-12 | 59 |
| date\_discharge | Validation | 40 | 0.99 | 2020-05-07 | 2021-01-14 | 2020-10-27 | 177 |

**Variable type: factor**

| skim\_variable | cohort | n\_missing | complete\_rate | ordered | n\_unique | top\_counts |
| --- | --- | --- | --- | --- | --- | --- |
| outcome\_fct | Derivation | 0 | 1 | FALSE | 4 | 0: 1363, 1: 162, 2: 71, 3: 29 |
| outcome\_fct | Validation | 0 | 1 | FALSE | 4 | 0: 2780, 1: 213, 2: 117, 3: 21 |

**Variable type: logical**

| skim\_variable | cohort | n\_missing | complete\_rate | mean | count |
| --- | --- | --- | --- | --- | --- |
| telehealth | Derivation | 0 | 1.00 | 0.99 | TRU: 1609, FAL: 16 |
| telehealth | Validation | 0 | 1.00 | 0.99 | TRU: 3089, FAL: 42 |
| outpatient | Derivation | 0 | 1.00 | 0.13 | FAL: 1414, TRU: 211 |
| outpatient | Validation | 0 | 1.00 | 0.10 | FAL: 2823, TRU: 308 |
| inpatient | Derivation | 0 | 1.00 | 0.06 | FAL: 1525, TRU: 100 |
| inpatient | Validation | 0 | 1.00 | 0.04 | FAL: 2993, TRU: 138 |
| icu | Derivation | 0 | 1.00 | 0.02 | FAL: 1598, TRU: 27 |
| icu | Validation | 0 | 1.00 | 0.01 | FAL: 3111, TRU: 20 |
| mortality | Derivation | 0 | 1.00 | 0.00 | FAL: 1619, TRU: 6 |
| mortality | Validation | 0 | 1.00 | 0.00 | FAL: 3127, TRU: 4 |
| sex | Derivation | 0 | 1.00 | 0.49 | FAL: 826, TRU: 799 |
| sex | Validation | 0 | 1.00 | 0.53 | TRU: 1656, FAL: 1475 |
| dm\_i | Derivation | 207 | 0.87 | 0.01 | FAL: 1409, TRU: 9 |
| dm\_i | Validation | 40 | 0.99 | 0.00 | FAL: 3081, TRU: 10 |
| dm\_ii | Derivation | 203 | 0.88 | 0.03 | FAL: 1382, TRU: 40 |
| dm\_ii | Validation | 40 | 0.99 | 0.02 | FAL: 3014, TRU: 77 |
| cardiovascular\_disease | Derivation | 195 | 0.88 | 0.08 | FAL: 1309, TRU: 121 |
| cardiovascular\_disease | Validation | 40 | 0.99 | 0.05 | FAL: 2930, TRU: 161 |
| hypertension | Derivation | 185 | 0.89 | 0.16 | FAL: 1211, TRU: 229 |
| hypertension | Validation | 40 | 0.99 | 0.11 | FAL: 2751, TRU: 340 |
| pulmonary\_disease | Derivation | 199 | 0.88 | 0.09 | FAL: 1299, TRU: 127 |
| pulmonary\_disease | Validation | 40 | 0.99 | 0.04 | FAL: 2973, TRU: 118 |
| chronic\_kidney\_disease | Derivation | 206 | 0.87 | 0.01 | FAL: 1407, TRU: 12 |
| chronic\_kidney\_disease | Validation | 40 | 0.99 | 0.00 | FAL: 3078, TRU: 13 |
| current\_cancer | Derivation | 207 | 0.87 | 0.01 | FAL: 1409, TRU: 9 |
| current\_cancer | Validation | 40 | 0.99 | 0.00 | FAL: 3084, TRU: 7 |
| prior\_cancer | Derivation | 204 | 0.87 | 0.04 | FAL: 1369, TRU: 52 |
| prior\_cancer | Validation | 40 | 0.99 | 0.02 | FAL: 3044, TRU: 47 |
| smoking\_current\_yes | Derivation | 246 | 0.85 | 0.06 | FAL: 1295, TRU: 84 |
| smoking\_current\_yes | Validation | 158 | 0.95 | 0.10 | FAL: 2662, TRU: 311 |
| smoking\_prior | Derivation | 841 | 0.48 | 0.41 | FAL: 462, TRU: 322 |
| smoking\_prior | Validation | 1055 | 0.66 | 0.45 | FAL: 1145, TRU: 931 |
| icd10\_htn | Derivation | 0 | 1.00 | 0.17 | FAL: 1350, TRU: 275 |
| icd10\_htn | Validation | 0 | 1.00 | 0.14 | FAL: 2680, TRU: 451 |
| antihypertensive | Derivation | 0 | 1.00 | 0.19 | FAL: 1309, TRU: 316 |
| antihypertensive | Validation | 0 | 1.00 | 0.15 | FAL: 2670, TRU: 461 |
| htn\_retrospective | Derivation | 0 | 1.00 | 0.15 | FAL: 1388, TRU: 237 |
| htn\_retrospective | Validation | 0 | 1.00 | 0.12 | FAL: 2764, TRU: 367 |
| icd10\_ihd | Derivation | 0 | 1.00 | 0.05 | FAL: 1546, TRU: 79 |
| icd10\_ihd | Validation | 0 | 1.00 | 0.04 | FAL: 3011, TRU: 120 |
| ihd\_drugs | Derivation | 0 | 1.00 | 0.10 | FAL: 1462, TRU: 163 |
| ihd\_drugs | Validation | 0 | 1.00 | 0.07 | FAL: 2905, TRU: 226 |
| ihd\_retrospective | Derivation | 0 | 1.00 | 0.04 | FAL: 1560, TRU: 65 |
| ihd\_retrospective | Validation | 0 | 1.00 | 0.03 | FAL: 3031, TRU: 100 |
| icd10\_chf | Derivation | 0 | 1.00 | 0.01 | FAL: 1608, TRU: 17 |
| icd10\_chf | Validation | 0 | 1.00 | 0.01 | FAL: 3098, TRU: 33 |
| chf\_drugs | Derivation | 0 | 1.00 | 0.20 | FAL: 1293, TRU: 332 |
| chf\_drugs | Validation | 0 | 1.00 | 0.16 | FAL: 2631, TRU: 500 |
| chf\_retrospective | Derivation | 0 | 1.00 | 0.01 | FAL: 1608, TRU: 17 |
| chf\_retrospective | Validation | 0 | 1.00 | 0.01 | FAL: 3102, TRU: 29 |
| icd10\_valvular | Derivation | 0 | 1.00 | 0.01 | FAL: 1606, TRU: 19 |
| icd10\_valvular | Validation | 0 | 1.00 | 0.01 | FAL: 3112, TRU: 19 |
| icd10\_arrhythmia | Derivation | 0 | 1.00 | 0.07 | FAL: 1517, TRU: 108 |
| icd10\_arrhythmia | Validation | 0 | 1.00 | 0.05 | FAL: 2972, TRU: 159 |
| icd10\_dm | Derivation | 0 | 1.00 | 0.16 | FAL: 1364, TRU: 261 |
| icd10\_dm | Validation | 0 | 1.00 | 0.15 | FAL: 2661, TRU: 470 |
| diabetes\_drugs | Derivation | 0 | 1.00 | 0.08 | FAL: 1493, TRU: 132 |
| diabetes\_drugs | Validation | 0 | 1.00 | 0.07 | FAL: 2924, TRU: 207 |
| diabetes\_retrospective | Derivation | 0 | 1.00 | 0.04 | FAL: 1564, TRU: 61 |
| diabetes\_retrospective | Validation | 0 | 1.00 | 0.04 | FAL: 3012, TRU: 119 |
| icd10\_pulm | Derivation | 0 | 1.00 | 0.19 | FAL: 1311, TRU: 314 |
| icd10\_pulm | Validation | 0 | 1.00 | 0.16 | FAL: 2636, TRU: 495 |
| pulm\_drugs | Derivation | 0 | 1.00 | 0.12 | FAL: 1429, TRU: 196 |
| pulm\_drugs | Validation | 0 | 1.00 | 0.09 | FAL: 2862, TRU: 269 |
| pulm\_retrospective | Derivation | 0 | 1.00 | 0.08 | FAL: 1493, TRU: 132 |
| pulm\_retrospective | Validation | 0 | 1.00 | 0.06 | FAL: 2946, TRU: 185 |
| icd10\_copd | Derivation | 0 | 1.00 | 0.02 | FAL: 1595, TRU: 30 |
| icd10\_copd | Validation | 0 | 1.00 | 0.02 | FAL: 3060, TRU: 71 |
| copd\_drugs | Derivation | 0 | 1.00 | 0.12 | FAL: 1429, TRU: 196 |
| copd\_drugs | Validation | 0 | 1.00 | 0.09 | FAL: 2862, TRU: 269 |
| copd\_retrospective | Derivation | 0 | 1.00 | 0.01 | FAL: 1605, TRU: 20 |
| copd\_retrospective | Validation | 0 | 1.00 | 0.02 | FAL: 3084, TRU: 47 |
| icd10\_asthma | Derivation | 0 | 1.00 | 0.18 | FAL: 1331, TRU: 294 |
| icd10\_asthma | Validation | 0 | 1.00 | 0.15 | FAL: 2670, TRU: 461 |
| asthma\_drugs | Derivation | 0 | 1.00 | 0.12 | FAL: 1429, TRU: 196 |
| asthma\_drugs | Validation | 0 | 1.00 | 0.09 | FAL: 2862, TRU: 269 |
| asthma\_retrospective | Derivation | 0 | 1.00 | 0.07 | FAL: 1504, TRU: 121 |
| asthma\_retrospective | Validation | 0 | 1.00 | 0.05 | FAL: 2964, TRU: 167 |
| icd10\_interstitial | Derivation | 0 | 1.00 | 0.00 | FAL: 1623, TRU: 2 |
| icd10\_interstitial | Validation | 0 | 1.00 | 0.01 | FAL: 3115, TRU: 16 |
| icd10\_osa | Derivation | 0 | 1.00 | 0.03 | FAL: 1572, TRU: 53 |
| icd10\_osa | Validation | 0 | 1.00 | 0.02 | FAL: 3053, TRU: 78 |
| ckd\_retrospective | Derivation | 0 | 1.00 | 0.02 | FAL: 1590, TRU: 35 |
| ckd\_retrospective | Validation | 0 | 1.00 | 0.03 | FAL: 3051, TRU: 80 |
| icd10\_cancer | Derivation | 0 | 1.00 | 0.05 | FAL: 1545, TRU: 80 |
| icd10\_cancer | Validation | 0 | 1.00 | 0.04 | FAL: 3008, TRU: 123 |
| icd10\_heart\_disease | Derivation | 0 | 1.00 | 0.11 | FAL: 1454, TRU: 171 |
| icd10\_heart\_disease | Validation | 0 | 1.00 | 0.08 | FAL: 2884, TRU: 247 |
| icd10\_pulmonary\_disease | Derivation | 0 | 1.00 | 0.08 | FAL: 1492, TRU: 133 |
| icd10\_pulmonary\_disease | Validation | 0 | 1.00 | 0.06 | FAL: 2931, TRU: 200 |
| fever | Derivation | 116 | 0.93 | 0.45 | FAL: 824, TRU: 685 |
| fever | Validation | 40 | 0.99 | 0.29 | FAL: 2186, TRU: 905 |
| no\_fever | Derivation | 157 | 0.90 | 0.51 | TRU: 746, FAL: 722 |
| no\_fever | Validation | 40 | 0.99 | 0.22 | FAL: 2416, TRU: 675 |
| chills | Derivation | 185 | 0.89 | 0.39 | FAL: 872, TRU: 568 |
| chills | Validation | 40 | 0.99 | 0.20 | FAL: 2473, TRU: 618 |
| dry\_cough | Derivation | 107 | 0.93 | 0.54 | TRU: 825, FAL: 693 |
| dry\_cough | Validation | 40 | 0.99 | 0.31 | FAL: 2119, TRU: 972 |
| wet\_cough | Derivation | 202 | 0.88 | 0.19 | FAL: 1153, TRU: 270 |
| wet\_cough | Validation | 40 | 0.99 | 0.09 | FAL: 2809, TRU: 282 |
| dyspnea\_rest | Derivation | 206 | 0.87 | 0.05 | FAL: 1349, TRU: 70 |
| dyspnea\_rest | Validation | 40 | 0.99 | 0.01 | FAL: 3062, TRU: 29 |
| dyspnea\_exertion | Derivation | 200 | 0.88 | 0.23 | FAL: 1097, TRU: 328 |
| dyspnea\_exertion | Validation | 40 | 0.99 | 0.05 | FAL: 2938, TRU: 153 |
| sob | Derivation | 200 | 0.88 | 0.20 | FAL: 1144, TRU: 281 |
| sob | Validation | 40 | 0.99 | 0.08 | FAL: 2855, TRU: 236 |
| sore\_throat | Derivation | 167 | 0.90 | 0.38 | FAL: 901, TRU: 557 |
| sore\_throat | Validation | 40 | 0.99 | 0.34 | FAL: 2051, TRU: 1040 |
| runny\_nose | Derivation | 160 | 0.90 | 0.40 | FAL: 878, TRU: 587 |
| runny\_nose | Validation | 40 | 0.99 | 0.22 | FAL: 2405, TRU: 686 |
| headache | Derivation | 160 | 0.90 | 0.63 | TRU: 925, FAL: 540 |
| headache | Validation | 40 | 0.99 | 0.43 | FAL: 1774, TRU: 1317 |
| muscle\_bone\_pain | Derivation | 132 | 0.92 | 0.59 | TRU: 883, FAL: 610 |
| muscle\_bone\_pain | Validation | 40 | 0.99 | 0.38 | FAL: 1907, TRU: 1184 |
| anorexia | Derivation | 201 | 0.88 | 0.33 | FAL: 961, TRU: 463 |
| anorexia | Validation | 40 | 0.99 | 0.11 | FAL: 2748, TRU: 343 |
| nausea | Derivation | 207 | 0.87 | 0.17 | FAL: 1173, TRU: 245 |
| nausea | Validation | 40 | 0.99 | 0.07 | FAL: 2874, TRU: 217 |
| vomiting | Derivation | 208 | 0.87 | 0.03 | FAL: 1375, TRU: 42 |
| vomiting | Validation | 40 | 0.99 | 0.01 | FAL: 3049, TRU: 42 |
| anosmia | Derivation | 203 | 0.88 | 0.38 | FAL: 886, TRU: 536 |
| anosmia | Validation | 40 | 0.99 | 0.09 | FAL: 2813, TRU: 278 |
| aguesia | Derivation | 204 | 0.87 | 0.41 | FAL: 841, TRU: 580 |
| aguesia | Validation | 40 | 0.99 | 0.09 | FAL: 2798, TRU: 293 |
| abdominal\_pain | Derivation | 205 | 0.87 | 0.14 | FAL: 1221, TRU: 199 |
| abdominal\_pain | Validation | 40 | 0.99 | 0.06 | FAL: 2919, TRU: 172 |
| diarrhea | Derivation | 205 | 0.87 | 0.19 | FAL: 1145, TRU: 275 |
| diarrhea | Validation | 40 | 0.99 | 0.09 | FAL: 2827, TRU: 264 |
| lethargy | Derivation | 129 | 0.92 | 0.61 | TRU: 917, FAL: 579 |
| lethargy | Validation | 40 | 0.99 | 0.40 | FAL: 1861, TRU: 1230 |
| diabetes\_prospective | Derivation | 202 | 0.88 | 0.03 | FAL: 1374, TRU: 49 |
| diabetes\_prospective | Validation | 40 | 0.99 | 0.03 | FAL: 3004, TRU: 87 |
| htn\_prospective | Derivation | 185 | 0.89 | 0.16 | FAL: 1211, TRU: 229 |
| htn\_prospective | Validation | 40 | 0.99 | 0.11 | FAL: 2751, TRU: 340 |
| heart\_prospective | Derivation | 195 | 0.88 | 0.08 | FAL: 1309, TRU: 121 |
| heart\_prospective | Validation | 40 | 0.99 | 0.05 | FAL: 2930, TRU: 161 |
| pulm\_prospective | Derivation | 199 | 0.88 | 0.09 | FAL: 1299, TRU: 127 |
| pulm\_prospective | Validation | 40 | 0.99 | 0.04 | FAL: 2973, TRU: 118 |
| ckd\_prospective | Derivation | 206 | 0.87 | 0.01 | FAL: 1407, TRU: 12 |
| ckd\_prospective | Validation | 40 | 0.99 | 0.00 | FAL: 3078, TRU: 13 |
| cancer\_prospective | Derivation | 203 | 0.88 | 0.04 | FAL: 1362, TRU: 60 |
| cancer\_prospective | Validation | 40 | 0.99 | 0.02 | FAL: 3037, TRU: 54 |
| smoking\_current | Derivation | 246 | 0.85 | 0.06 | FAL: 1295, TRU: 84 |
| smoking\_current | Validation | 158 | 0.95 | 0.10 | FAL: 2662, TRU: 311 |
| clinical\_score\_fct | Derivation | 36 | 0.98 | 0.18 | FAL: 1296, TRU: 293 |
| clinical\_score\_fct | Validation | 270 | 0.91 | 0.06 | FAL: 2684, TRU: 177 |
| n\_hospital\_fct | Derivation | 0 | 1.00 | 0.08 | FAL: 1503, TRU: 122 |
| n\_hospital\_fct | Validation | 0 | 1.00 | 0.09 | FAL: 2855, TRU: 276 |
| distr\_fct | Derivation | 0 | 1.00 | 0.72 | TRU: 1166, FAL: 459 |
| distr\_fct | Validation | 0 | 1.00 | 0.76 | TRU: 2394, FAL: 737 |
| flulike\_symptoms | Derivation | 37 | 0.98 | 0.91 | TRU: 1446, FAL: 142 |
| flulike\_symptoms | Validation | 40 | 0.99 | 0.76 | TRU: 2335, FAL: 756 |
| upper\_respiratory | Derivation | 129 | 0.92 | 0.76 | TRU: 1135, FAL: 361 |
| upper\_respiratory | Validation | 40 | 0.99 | 0.52 | TRU: 1592, FAL: 1499 |
| lower\_respiratory | Derivation | 186 | 0.89 | 0.45 | FAL: 790, TRU: 649 |
| lower\_respiratory | Validation | 40 | 0.99 | 0.18 | FAL: 2534, TRU: 557 |
| gastrointestinal | Derivation | 204 | 0.87 | 0.36 | FAL: 916, TRU: 505 |
| gastrointestinal | Validation | 40 | 0.99 | 0.17 | FAL: 2578, TRU: 513 |
| logistic\_outpatient | Derivation | 0 | 1.00 | 0.16 | FAL: 1363, TRU: 262 |
| logistic\_outpatient | Validation | 0 | 1.00 | 0.11 | FAL: 2780, TRU: 351 |
| logistic\_inpatient | Derivation | 0 | 1.00 | 0.06 | FAL: 1525, TRU: 100 |
| logistic\_inpatient | Validation | 0 | 1.00 | 0.04 | FAL: 2993, TRU: 138 |
| logistic\_icu | Derivation | 0 | 1.00 | 0.02 | FAL: 1596, TRU: 29 |
| logistic\_icu | Validation | 0 | 1.00 | 0.01 | FAL: 3110, TRU: 21 |

**Variable type: numeric**

| skim\_variable | cohort | n\_missing | complete\_rate | mean | sd | p0 | p25 | p50 | p75 | p100 | hist |
| --- | --- | --- | --- | --- | --- | --- | --- | --- | --- | --- | --- |
| age | Derivation | 0 | 1.00 | 44.43 | 15.75 | 18.0 | 31.00 | 45.0 | 56.00 | 93.0 | ▇▇▇▃▁ |
| age | Validation | 0 | 1.00 | 40.78 | 16.41 | 18.0 | 27.00 | 37.0 | 53.00 | 99.0 | ▇▅▃▁▁ |
| height | Derivation | 1110 | 0.32 | 174.90 | 9.85 | 150.0 | 168.00 | 175.0 | 182.00 | 207.0 | ▂▇▇▃▁ |
| height | Validation | 301 | 0.90 | 174.63 | 9.80 | 116.0 | 167.00 | 175.0 | 182.00 | 202.0 | ▁▁▃▇▂ |
| weight | Derivation | 1100 | 0.32 | 83.53 | 18.08 | 48.0 | 70.00 | 82.0 | 94.00 | 166.0 | ▅▇▃▁▁ |
| weight | Validation | 316 | 0.90 | 81.77 | 18.08 | 41.0 | 69.00 | 80.0 | 93.00 | 170.0 | ▃▇▃▁▁ |
| clinical | Derivation | 36 | 0.98 | 0.23 | 0.51 | 0.0 | 0.00 | 0.0 | 0.00 | 2.0 | ▇▁▂▁▁ |
| clinical | Validation | 270 | 0.91 | 0.06 | 0.26 | 0.0 | 0.00 | 0.0 | 0.00 | 2.0 | ▇▁▁▁▁ |
| retrospective\_height | Derivation | 1477 | 0.09 | 174.58 | 9.75 | 151.0 | 168.00 | 175.0 | 181.00 | 206.0 | ▂▇▇▃▁ |
| retrospective\_height | Validation | 2917 | 0.07 | 172.67 | 11.26 | 84.0 | 165.25 | 173.0 | 180.00 | 201.0 | ▁▁▁▇▅ |
| retrospective\_weight | Derivation | 1445 | 0.11 | 89.23 | 20.43 | 51.0 | 74.00 | 88.0 | 101.00 | 164.0 | ▅▇▅▂▁ |
| retrospective\_weight | Validation | 2852 | 0.09 | 88.18 | 20.95 | 33.0 | 75.00 | 87.0 | 100.00 | 169.0 | ▂▇▆▁▁ |
| n\_visits | Derivation | 0 | 1.00 | 8.42 | 8.97 | 0.0 | 2.00 | 6.0 | 11.00 | 76.0 | ▇▁▁▁▁ |
| n\_visits | Validation | 0 | 1.00 | 8.07 | 9.24 | 0.0 | 2.00 | 5.0 | 11.00 | 78.0 | ▇▁▁▁▁ |
| n\_hospital | Derivation | 0 | 1.00 | 0.10 | 0.39 | 0.0 | 0.00 | 0.0 | 0.00 | 5.0 | ▇▁▁▁▁ |
| n\_hospital | Validation | 0 | 1.00 | 0.12 | 0.50 | 0.0 | 0.00 | 0.0 | 0.00 | 7.0 | ▇▁▁▁▁ |
| n\_prescriptions | Derivation | 0 | 1.00 | 9.21 | 16.08 | 0.0 | 1.00 | 5.0 | 11.00 | 234.0 | ▇▁▁▁▁ |
| n\_prescriptions | Validation | 0 | 1.00 | 8.96 | 20.06 | 0.0 | 0.00 | 4.0 | 10.00 | 505.0 | ▇▁▁▁▁ |
| time\_diagnosis | Derivation | 0 | 1.00 | 29.37 | 9.71 | 2.0 | 23.00 | 29.0 | 36.00 | 64.0 | ▁▅▇▂▁ |
| time\_diagnosis | Validation | 0 | 1.00 | 230.31 | 31.38 | 69.0 | 216.00 | 229.0 | 245.00 | 309.0 | ▁▁▂▇▂ |
| time\_symptoms | Derivation | 54 | 0.97 | 5.68 | 5.63 | -30.0 | 3.00 | 5.0 | 8.00 | 51.0 | ▁▂▇▁▁ |
| time\_symptoms | Validation | 146 | 0.95 | 1.89 | 3.51 | -31.0 | 1.00 | 1.0 | 3.00 | 80.0 | ▁▇▁▁▁ |
| time\_followup | Derivation | 11 | 0.99 | 17.34 | 5.35 | 3.0 | 14.00 | 15.0 | 19.00 | 55.0 | ▁▇▁▁▁ |
| time\_followup | Validation | 40 | 0.99 | 15.16 | 3.51 | 2.0 | 14.00 | 15.0 | 16.00 | 67.0 | ▇▃▁▁▁ |
| bmi\_prospective | Derivation | 1115 | 0.31 | 27.16 | 5.23 | 16.5 | 23.42 | 26.2 | 30.10 | 56.1 | ▅▇▂▁▁ |
| bmi\_prospective | Validation | 326 | 0.90 | 26.72 | 5.06 | 15.9 | 23.10 | 25.9 | 29.40 | 57.4 | ▆▇▂▁▁ |
| bmi\_retrospective | Derivation | 1477 | 0.09 | 29.61 | 6.15 | 18.4 | 25.28 | 28.7 | 33.12 | 51.4 | ▅▇▅▁▁ |
| bmi\_retrospective | Validation | 2917 | 0.07 | 29.77 | 8.77 | 16.9 | 25.22 | 28.7 | 33.18 | 119.0 | ▇▁▁▁▁ |
| outcome | Derivation | 0 | 1.00 | 0.24 | 0.61 | 0.0 | 0.00 | 0.0 | 0.00 | 3.0 | ▇▁▁▁▁ |
| outcome | Validation | 0 | 1.00 | 0.16 | 0.50 | 0.0 | 0.00 | 0.0 | 0.00 | 3.0 | ▇▁▁▁▁ |
| clinical\_score\_fct\_total | Derivation | 36 | 0.98 | 0.23 | 0.51 | 0.0 | 0.00 | 0.0 | 0.00 | 2.0 | ▇▁▂▁▁ |
| clinical\_score\_fct\_total | Validation | 270 | 0.91 | 0.06 | 0.26 | 0.0 | 0.00 | 0.0 | 0.00 | 2.0 | ▇▁▁▁▁ |
| bmi | Derivation | 1019 | 0.37 | 27.49 | 5.44 | 16.5 | 23.70 | 26.5 | 30.40 | 56.1 | ▅▇▂▁▁ |
| bmi | Validation | 298 | 0.90 | 26.73 | 5.07 | 15.9 | 23.10 | 26.0 | 29.40 | 57.4 | ▆▇▂▁▁ |

```
vars_table1 <- c(
  "age",
  "sex",
  "outcome",
  "height",
  "weight",
  "bmi_prospective",
  "smoking_current",
  "smoking_prior",
  "clinical_score_fct",
  "htn_prospective",
  "icd10_htn",
  "antihypertensive",
  "heart_prospective",
  "icd10_ihd",
  "ihd_drugs",
  "icd10_heart_disease",
  "diabetes_prospective",
  "icd10_dm",
  "diabetes_drugs",
  "pulm_prospective",
  "icd10_pulm",
  "pulm_drugs",
  "icd10_pulmonary_disease",
  "ckd_prospective",
  "ckd_retrospective",
  "cancer_prospective",
  "icd10_cancer",
  "n_visits",
  "n_hospital",
  "n_prescriptions",
  "time_symptoms",
  "time_followup",
  "flulike_symptoms",
  "upper_respiratory",
  "lower_respiratory",
  "gastrointestinal"
)

catvars_table1 <- c(
  "sex",
  "outcome",
  "smoking_current",
  "smoking_prior",
  "clinical_score_fct",
  "htn_prospective",
  "icd10_htn",
  "antihypertensive",
  "heart_prospective",
  "icd10_ihd",
  "ihd_drugs",
  "icd10_heart_disease",
  "diabetes_prospective",
  "icd10_dm",
  "diabetes_drugs",
  "pulm_prospective",
  "icd10_pulm",
  "pulm_drugs",
  "icd10_pulmonary_disease",
  "ckd_prospective",
  "ckd_retrospective",
  "cancer_prospective",
  "icd10_cancer",
  "flulike_symptoms",
  "upper_respiratory",
  "lower_respiratory",
  "gastrointestinal"
)
```

Here we show the code that generates the data in Table 1 of the manuscript. Several additional variables are included in the version shown here. Some of these additional predictors are used in the multiple imputation procedure.

```
table1 <- CreateTableOne(
  vars = vars_table1,
  data = df_outcome %>% filter(excluded == "Included"),
  strata = "cohort",
  factorVars = catvars_table1
)

kableone(table1)
```

|  | Derivation | Validation | p | test |
| --- | --- | --- | --- | --- |
| n | 1625 | 3131 |  |  |
| age (mean (SD)) | 44.43 (15.75) | 40.78 (16.41) | <0.001 |  |
| sex = 1 (%) | 799 (49.2) | 1656 (52.9) | 0.016 |  |
| outcome (%) |  |  | <0.001 |  |
| 0 | 1363 (83.9) | 2780 (88.8) |  |  |
| 1 | 162 (10.0) | 213 ( 6.8) |  |  |
| 2 | 71 ( 4.4) | 117 ( 3.7) |  |  |
| 3 | 29 ( 1.8) | 21 ( 0.7) |  |  |
| height (mean (SD)) | 174.90 (9.85) | 174.63 (9.80) | 0.561 |  |
| weight (mean (SD)) | 83.53 (18.08) | 81.77 (18.08) | 0.040 |  |
| bmi\_prospective (mean (SD)) | 27.16 (5.23) | 26.72 (5.06) | 0.072 |  |
| smoking\_current = 1 (%) | 84 ( 6.1) | 311 (10.5) | <0.001 |  |
| smoking\_prior = 1 (%) | 322 (41.1) | 931 (44.8) | 0.076 |  |
| clinical\_score\_fct = 1 (%) | 293 (18.4) | 177 ( 6.2) | <0.001 |  |
| htn\_prospective = 1 (%) | 229 (15.9) | 340 (11.0) | <0.001 |  |
| icd10\_htn = 1 (%) | 275 (16.9) | 451 (14.4) | 0.025 |  |
| antihypertensive = 1 (%) | 316 (19.4) | 461 (14.7) | <0.001 |  |
| heart\_prospective = 1 (%) | 121 ( 8.5) | 161 ( 5.2) | <0.001 |  |
| icd10\_ihd = 1 (%) | 79 ( 4.9) | 120 ( 3.8) | 0.109 |  |
| ihd\_drugs = 1 (%) | 163 (10.0) | 226 ( 7.2) | 0.001 |  |
| icd10\_heart\_disease = 1 (%) | 171 (10.5) | 247 ( 7.9) | 0.003 |  |
| diabetes\_prospective = 1 (%) | 49 ( 3.4) | 87 ( 2.8) | 0.292 |  |
| icd10\_dm = 1 (%) | 261 (16.1) | 470 (15.0) | 0.363 |  |
| diabetes\_drugs = 1 (%) | 132 ( 8.1) | 207 ( 6.6) | 0.063 |  |
| pulm\_prospective = 1 (%) | 127 ( 8.9) | 118 ( 3.8) | <0.001 |  |
| icd10\_pulm = 1 (%) | 314 (19.3) | 495 (15.8) | 0.003 |  |
| pulm\_drugs = 1 (%) | 196 (12.1) | 269 ( 8.6) | <0.001 |  |
| icd10\_pulmonary\_disease = 1 (%) | 133 ( 8.2) | 200 ( 6.4) | 0.025 |  |
| ckd\_prospective = 1 (%) | 12 ( 0.8) | 13 ( 0.4) | 0.117 |  |
| ckd\_retrospective = 1 (%) | 35 ( 2.2) | 80 ( 2.6) | 0.450 |  |
| cancer\_prospective = 1 (%) | 60 ( 4.2) | 54 ( 1.7) | <0.001 |  |
| icd10\_cancer = 1 (%) | 80 ( 4.9) | 123 ( 3.9) | 0.125 |  |
| n\_visits (mean (SD)) | 8.42 (8.97) | 8.07 (9.24) | 0.206 |  |
| n\_hospital (mean (SD)) | 0.10 (0.39) | 0.12 (0.50) | 0.050 |  |
| n\_prescriptions (mean (SD)) | 9.21 (16.08) | 8.96 (20.06) | 0.667 |  |
| time\_symptoms (mean (SD)) | 5.68 (5.63) | 1.89 (3.51) | <0.001 |  |
| time\_followup (mean (SD)) | 17.34 (5.35) | 15.16 (3.51) | <0.001 |  |
| flulike\_symptoms = 1 (%) | 1446 (91.1) | 2335 (75.5) | <0.001 |  |
| upper\_respiratory = 1 (%) | 1135 (75.9) | 1592 (51.5) | <0.001 |  |
| lower\_respiratory = 1 (%) | 649 (45.1) | 557 (18.0) | <0.001 |  |
| gastrointestinal = 1 (%) | 505 (35.5) | 513 (16.6) | <0.001 |  |

Here we show the number and percent missing for each of the variables, stratified by derivation and validation cohorts.

```
summary(
  CreateContTable(
    vars = vars_table1,
    data = df_outcome %>% filter(excluded == "Included"),
    strata = "cohort",
    test = FALSE,
    smd = FALSE,
    funcNames = c("n", "miss", "p.miss")
  ))
```

```
## cohort: Derivation
##                            n miss p.miss
## age                     1625    0   0.00
## sex                     1625    0   0.00
## outcome                 1625    0   0.00
## height                  1625 1110  68.31
## weight                  1625 1100  67.69
## bmi_prospective         1625 1115  68.62
## smoking_current         1625  246  15.14
## smoking_prior           1625  841  51.75
## clinical_score_fct      1625   36   2.22
## htn_prospective         1625  185  11.38
## icd10_htn               1625    0   0.00
## antihypertensive        1625    0   0.00
## heart_prospective       1625  195  12.00
## icd10_ihd               1625    0   0.00
## ihd_drugs               1625    0   0.00
## icd10_heart_disease     1625    0   0.00
## diabetes_prospective    1625  202  12.43
## icd10_dm                1625    0   0.00
## diabetes_drugs          1625    0   0.00
## pulm_prospective        1625  199  12.25
## icd10_pulm              1625    0   0.00
## pulm_drugs              1625    0   0.00
## icd10_pulmonary_disease 1625    0   0.00
## ckd_prospective         1625  206  12.68
## ckd_retrospective       1625    0   0.00
## cancer_prospective      1625  203  12.49
## icd10_cancer            1625    0   0.00
## n_visits                1625    0   0.00
## n_hospital              1625    0   0.00
## n_prescriptions         1625    0   0.00
## time_symptoms           1625   54   3.32
## time_followup           1625   11   0.68
## flulike_symptoms        1625   37   2.28
## upper_respiratory       1625  129   7.94
## lower_respiratory       1625  186  11.45
## gastrointestinal        1625  204  12.55
## ------------------------------------------------------------ 
## cohort: Validation
##                            n miss p.miss
## age                     3131    0    0.0
## sex                     3131    0    0.0
## outcome                 3131    0    0.0
## height                  3131  301    9.6
## weight                  3131  316   10.1
## bmi_prospective         3131  326   10.4
## smoking_current         3131  158    5.0
## smoking_prior           3131 1055   33.7
## clinical_score_fct      3131  270    8.6
## htn_prospective         3131   40    1.3
## icd10_htn               3131    0    0.0
## antihypertensive        3131    0    0.0
## heart_prospective       3131   40    1.3
## icd10_ihd               3131    0    0.0
## ihd_drugs               3131    0    0.0
## icd10_heart_disease     3131    0    0.0
## diabetes_prospective    3131   40    1.3
## icd10_dm                3131    0    0.0
## diabetes_drugs          3131    0    0.0
## pulm_prospective        3131   40    1.3
## icd10_pulm              3131    0    0.0
## pulm_drugs              3131    0    0.0
## icd10_pulmonary_disease 3131    0    0.0
## ckd_prospective         3131   40    1.3
## ckd_retrospective       3131    0    0.0
## cancer_prospective      3131   40    1.3
## icd10_cancer            3131    0    0.0
## n_visits                3131    0    0.0
## n_hospital              3131    0    0.0
## n_prescriptions         3131    0    0.0
## time_symptoms           3131  146    4.7
## time_followup           3131   40    1.3
## flulike_symptoms        3131   40    1.3
## upper_respiratory       3131   40    1.3
## lower_respiratory       3131   40    1.3
## gastrointestinal        3131   40    1.3
```

The following code explores the distrribution of missing data more thoroughly. First we show a figure that summarises the number of individuals who have a certain number of missing variables (out of 14 included in the prognostic model). The left Y-axis shows the number of missing vaiables and the x-axis shows the number of individuals (with the exact number being displayed on the right Y-axis). First we show this for the combined derivation and validation cohorts.

```
naclust_all <- naclus(df = df_outcome %>%
                    filter(excluded == "Included") %>%
                    select(age, sex, bmi_prospective, htn_prospective,
                           heart_prospective, pulm_prospective,
                           diabetes_prospective, cancer_prospective,
                           smoking_current, flulike_symptoms, upper_respiratory,
                           lower_respiratory, gastrointestinal, clinical_score_fct)
                  )

naclust_derivation <- naclus(df = df_outcome %>%
                        filter(excluded == "Included", cohort == "Derivation") %>%
                        select(age, sex, bmi_prospective, htn_prospective,
                               heart_prospective, pulm_prospective,
                               diabetes_prospective, cancer_prospective,
                               smoking_current, flulike_symptoms, upper_respiratory,
                               lower_respiratory, gastrointestinal, clinical_score_fct)
)

naclust_validation <- naclus(df = df_outcome %>%
                               filter(excluded == "Included", cohort == "Validation") %>%
                               select(age, sex, bmi_prospective, htn_prospective,
                                      heart_prospective, pulm_prospective,
                                      diabetes_prospective, cancer_prospective,
                                      smoking_current, flulike_symptoms, upper_respiratory,
                                      lower_respiratory, gastrointestinal, clinical_score_fct)
)

naplot(naclust_all, which = "na per obs")
```

Then we show the same for the derivation (top) and validation cohort (bottom) seperately.

```
naplot(naclust_derivation, which = "na per obs")
```

```
naplot(naclust_validation, which = "na per obs")
```

Next we show two figures which summarize mean number of additional missing variables for observations for which the specific variable is missing (top) and the fraction of all individuals for which the specific variable is missing (bottom).

```
naplot(naclust_all, which = "mean na")
```

```
naplot(naclust_all, which = "na per var")
```

We show the same two figures for the derivation cohort

```
naplot(naclust_derivation, which = "mean na")
```

```
naplot(naclust_derivation, which = "na per var")
```

and for the validation cohort

```
naplot(naclust_validation, which = "mean na")
```

```
naplot(naclust_validation, which = "na per var")
```

### 4 Miscellaneous statistics presented in result

Number of individuals tested during the study period.

```
df_tests %>%
  count(kt) %>%
  nrow()
```

```
## [1] 175243
```

Number of individuals who were ever SARS-CoV-2 positive.

```
df_outcome %>% 
  nrow()
```

```
## [1] 6126
```

Number of individuals in the derivation cohort after applying exclusion criteria.

```
df_outcome %>%
  filter(
    cohort == "Derivation",
    excluded == "Included"
  ) %>%
  nrow()
```

```
## [1] 1625
```

Number of individuals excluded from derivation cohort with the reason given.

```
knitr::kable(
  df_outcome %>%
    filter(cohort == "Derivation") %>%
    count(excluded)
)
```

| excluded | n |
| --- | --- |
| Excluded, current infection excluded | 1 |
| Excluded, foreign citizen without NIN | 7 |
| Excluded, hospitalized due to complication of unrelated surgery | 1 |
| Excluded, incorrect documentation. PCR was never performed | 1 |
| Excluded, nursing home resident at the time of infection | 7 |
| Excluded, was infected during hospital stay | 9 |
| Excluded, was infected during inpatient rehabilitation | 1 |
| Excluded, younger than 18 years of age | 147 |
| Included | 1625 |

Number of individuals in the validation cohort after applying exclusion criteria.

```
df_outcome %>%
  filter(
    cohort == "Validation",
    excluded == "Included"
  ) %>%
  nrow()
```

```
## [1] 3131
```

Number of individuals excluded from validation cohort with the reason given.

```
knitr::kable(
  df_outcome %>%
    filter(cohort == "Validation") %>%
    count(excluded)
  )
```

| excluded | n |
| --- | --- |
| Excluded, current infection excluded | 214 |
| Excluded, diagnosed post-mortem | 1 |
| Excluded, foreign citizen without NIN | 368 |
| Excluded, nursing home resident at the time of infection | 24 |
| Excluded, was infected during hospital stay | 39 |
| Excluded, was infected during inpatient rehabilitation | 10 |
| Excluded, younger than 18 years of age | 540 |
| Included | 3131 |

Total number of the included patients for each outcome

```
knitr::kable(
  df_outcome %>%
    filter(excluded == "Included") %>%
    count(outcome)
)
```

| outcome | n |
| --- | --- |
| 0 | 4143 |
| 1 | 375 |
| 2 | 188 |
| 3 | 50 |

Distribution of the time from symptom onset to RT-PCR.

```
df_outcome %>%
  left_join(
    df_phone %>%
      select(kt, date_symptoms) %>%
      distinct()
    ) %>%
  filter(excluded == "Included") %>%
  mutate(time_symptoms_pcr = difftime(date_pcr, date_symptoms, units = "days")) %>%
  pull(time_symptoms_pcr) %>%
  quantile(.,
           probs = c(0, 0.01, 0.25, 0.5, 0.75, 0.99, 1),
           na.rm = TRUE)
```

```
## Time differences in days
##   0%   1%  25%  50%  75%  99% 100% 
##  -31   -4    1    2    4   20   80
```

Distribution of the time from RT-PCR to enrollment into telehealth monitoring

```
df_outcome %>%
  left_join(
    df_phone %>%
      filter(call_nr == 1) %>%
      select(kt, date) %>%
      distinct()
  ) %>%
  filter(excluded == "Included") %>%
  mutate(
    time_pcr_enrollment = difftime(date, date_pcr, units = "days")
  ) %>%
  pull(time_pcr_enrollment) %>%
  quantile(
    .,
    probs = c(0, 0.01, 0.25, 0.5, 0.75, 0.99, 1),
    na.rm = TRUE)
```

```
## Time differences in days
##   0%   1%  25%  50%  75%  99% 100% 
##  -12    0    0    0    1    4   34
```

Distribution of the follow-up time in telehealth monitoring

```
df_outcome %>%
  left_join(
    df_phone %>%
      filter(call_nr == 1) %>%
      select(kt, date, date_discharge) %>%
      distinct()
  ) %>%
  filter(excluded == "Included") %>%
  mutate(
    time_enrollment_discharge = difftime(date_discharge, date, units = "days")
  ) %>%
  pull(time_enrollment_discharge) %>%
  quantile(
    .,
    probs = c(0, 0.01, 0.25, 0.5, 0.75, 0.99, 1),
    na.rm = TRUE)
```

```
## Time differences in days
##   0%   1%  25%  50%  75%  99% 100% 
##  -11    7   14   15   16   32   67
```

### 5 Multiple imputation

Multiple imputation was performed using the function aregImpute(). In all cases the imputation procedure models continuous variables with a six knot restricted cubic splines. In the manuscript, 100 imputed datasets are constructed. for each of the imputations below. However, for the purposes of sharing the code and displaying the outputs of each code snippet, we have lowered the number of imputed datasets to 20 to save computational time. The first multiple imputation procedure ‘imputation\_outcomes’ is only used in the risk factor analysis, not the derivation or validation of the prognostic model.

```
imputation_outcomes <- aregImpute(
  formula = ~outcome_fct + age + sex + bmi + smoking_current + smoking_prior +
    htn_retrospective + ihd_retrospective + diabetes_retrospective + copd_retrospective +
    ckd_retrospective + icd10_cancer + n_visits + n_hospital_fct + n_prescriptions +
    time_diagnosis + time_symptoms + time_followup + distr,
  data = df_outcome %>% filter(excluded == "Included"),
  n.impute = 20,
  # n.impute = 100 in the true analysis, n.impute = 20 here for shorter computation
  nk = 6,
  tlinear = FALSE,
  x = TRUE,
  B = 200,
  group = df_outcome %>%
    filter(excluded == "Included") %>%
    pull(outcome_fct)
)
```

```
## Iteration 1 
Iteration 2 
Iteration 3 
Iteration 4 
Iteration 5 
Iteration 6 
Iteration 7 
Iteration 8 
Iteration 9 
Iteration 10 
Iteration 11 
Iteration 12 
Iteration 13 
Iteration 14 
Iteration 15 
Iteration 16 
Iteration 17 
Iteration 18 
Iteration 19 
Iteration 20 
Iteration 21 
Iteration 22 
Iteration 23
```

```
imputation_outcomes
```

```
## 
## Multiple Imputation using Bootstrap and PMM
## 
## aregImpute(formula = ~outcome_fct + age + sex + bmi + smoking_current + 
##     smoking_prior + htn_retrospective + ihd_retrospective + diabetes_retrospective + 
##     copd_retrospective + ckd_retrospective + icd10_cancer + n_visits + 
##     n_hospital_fct + n_prescriptions + time_diagnosis + time_symptoms + 
##     time_followup + distr, data = df_outcome %>% filter(excluded == 
##     "Included"), n.impute = 20, group = df_outcome %>% filter(excluded == 
##     "Included") %>% pull(outcome_fct), nk = 6, tlinear = FALSE, 
##     x = TRUE, B = 200)
## 
## n: 4756  p: 19   Imputations: 20     nk: 6 
## 
## Number of NAs:
##            outcome_fct                    age                    sex 
##                      0                      0                      0 
##                    bmi        smoking_current          smoking_prior 
##                   1317                    404                   1896 
##      htn_retrospective      ihd_retrospective diabetes_retrospective 
##                      0                      0                      0 
##     copd_retrospective      ckd_retrospective           icd10_cancer 
##                      0                      0                      0 
##               n_visits         n_hospital_fct        n_prescriptions 
##                      0                      0                      0 
##         time_diagnosis          time_symptoms          time_followup 
##                      0                    200                     51 
##                  distr 
##                      0 
## 
##                        type d.f.
## outcome_fct               c    3
## age                       s    5
## sex                       l    1
## bmi                       s    5
## smoking_current           l    1
## smoking_prior             l    1
## htn_retrospective         l    1
## ihd_retrospective         l    1
## diabetes_retrospective    l    1
## copd_retrospective        l    1
## ckd_retrospective         l    1
## icd10_cancer              l    1
## n_visits                  s    5
## n_hospital_fct            l    1
## n_prescriptions           s    4
## time_diagnosis            s    5
## time_symptoms             s    5
## time_followup             s    4
## distr                     c    7
## 
## R-squares for Predicting Non-Missing Values for Each Variable
## Using Last Imputations of Predictors
##             bmi smoking_current   smoking_prior   time_symptoms   time_followup 
##           0.241           0.204           0.325           0.339           0.386
```

```
par(mfrow=c(3,2))
plot(imputation_outcomes)
par(mfrow=c(1,1))
```

This imputation procedure is used for the derivation cohort

```
imputation_outcomes_derivation <-
  aregImpute(
    formula = ~ outcome_fct + age + sex + bmi_prospective + smoking_current++clinical_score_fct + htn_prospective +
      icd10_htn + antihypertensive + heart_prospective + icd10_heart_disease +
      diabetes_prospective + icd10_dm + diabetes_drugs + pulm_prospective +
      icd10_pulmonary_disease + ckd_prospective + ckd_retrospective +
      cancer_prospective + icd10_cancer +  n_visits + n_hospital_fct + n_prescriptions +
      time_symptoms + time_followup + flulike_symptoms + upper_respiratory +
      lower_respiratory + gastrointestinal + distr_fct,
    data = df_derivation %>% filter(excluded == "Included"),
    n.impute = 20,
    # n.impute = 100 in the true analysis, n.impute = 20 here for shorter computation
    nk = 6,
    tlinear = FALSE,
    x = TRUE,
    B = 200,
    group = df_derivation %>%
      filter(excluded == "Included") %>%
      pull(outcome_fct)
  )
```

```
## Iteration 1 
Iteration 2 
Iteration 3 
Iteration 4 
Iteration 5 
Iteration 6 
Iteration 7 
Iteration 8 
Iteration 9 
Iteration 10 
Iteration 11 
Iteration 12 
Iteration 13 
Iteration 14 
Iteration 15 
Iteration 16 
Iteration 17 
Iteration 18 
Iteration 19 
Iteration 20 
Iteration 21 
Iteration 22 
Iteration 23
```

```
imputation_outcomes_derivation
```

```
## 
## Multiple Imputation using Bootstrap and PMM
## 
## aregImpute(formula = ~outcome_fct + age + sex + bmi_prospective + 
##     smoking_current + +clinical_score_fct + htn_prospective + 
##     icd10_htn + antihypertensive + heart_prospective + icd10_heart_disease + 
##     diabetes_prospective + icd10_dm + diabetes_drugs + pulm_prospective + 
##     icd10_pulmonary_disease + ckd_prospective + ckd_retrospective + 
##     cancer_prospective + icd10_cancer + n_visits + n_hospital_fct + 
##     n_prescriptions + time_symptoms + time_followup + flulike_symptoms + 
##     upper_respiratory + lower_respiratory + gastrointestinal + 
##     distr_fct, data = df_derivation %>% filter(excluded == "Included"), 
##     n.impute = 20, group = df_derivation %>% filter(excluded == 
##         "Included") %>% pull(outcome_fct), nk = 6, tlinear = FALSE, 
##     x = TRUE, B = 200)
## 
## n: 1625  p: 30   Imputations: 20     nk: 6 
## 
## Number of NAs:
##             outcome_fct                     age                     sex 
##                       0                       0                       0 
##         bmi_prospective         smoking_current      clinical_score_fct 
##                    1115                     246                      36 
##         htn_prospective               icd10_htn        antihypertensive 
##                     185                       0                       0 
##       heart_prospective     icd10_heart_disease    diabetes_prospective 
##                     195                       0                     202 
##                icd10_dm          diabetes_drugs        pulm_prospective 
##                       0                       0                     199 
## icd10_pulmonary_disease         ckd_prospective       ckd_retrospective 
##                       0                     206                       0 
##      cancer_prospective            icd10_cancer                n_visits 
##                     203                       0                       0 
##          n_hospital_fct         n_prescriptions           time_symptoms 
##                       0                       0                      54 
##           time_followup        flulike_symptoms       upper_respiratory 
##                      11                      37                     129 
##       lower_respiratory        gastrointestinal               distr_fct 
##                     186                     204                       0 
## 
##                         type d.f.
## outcome_fct                c    3
## age                        s    5
## sex                        l    1
## bmi_prospective            s    5
## smoking_current            l    1
## clinical_score_fct         l    1
## htn_prospective            l    1
## icd10_htn                  l    1
## antihypertensive           l    1
## heart_prospective          l    1
## icd10_heart_disease        l    1
## diabetes_prospective       l    1
## icd10_dm                   l    1
## diabetes_drugs             l    1
## pulm_prospective           l    1
## icd10_pulmonary_disease    l    1
## ckd_prospective            l    1
## ckd_retrospective          l    1
## cancer_prospective         l    1
## icd10_cancer               l    1
## n_visits                   s    5
## n_hospital_fct             l    1
## n_prescriptions            s    5
## time_symptoms              s    5
## time_followup              s    5
## flulike_symptoms           l    1
## upper_respiratory          l    1
## lower_respiratory          l    1
## gastrointestinal           l    1
## distr_fct                  l    1
## 
## R-squares for Predicting Non-Missing Values for Each Variable
## Using Last Imputations of Predictors
##      bmi_prospective      smoking_current   clinical_score_fct 
##                0.430                0.101                0.309 
##      htn_prospective    heart_prospective diabetes_prospective 
##                0.761                0.558                0.478 
##     pulm_prospective      ckd_prospective   cancer_prospective 
##                0.305                0.223                0.526 
##        time_symptoms        time_followup     flulike_symptoms 
##                0.142                0.365                0.172 
##    upper_respiratory    lower_respiratory     gastrointestinal 
##                0.152                0.181                0.202
```

```
par(mfrow=c(1,3))
plot(imputation_outcomes_derivation)
```

```
par(mfrow=c(1,1))
```

This imputation procedure is used for the validation cohort

```
imputation_outcomes_validation <-
  aregImpute(
    formula = ~ outcome_fct + age + sex + bmi_prospective + smoking_current++clinical_score_fct + htn_prospective +
      icd10_htn + antihypertensive + heart_prospective + icd10_heart_disease +
      diabetes_prospective + icd10_dm + diabetes_drugs + pulm_prospective +
      icd10_pulmonary_disease + ckd_prospective + ckd_retrospective +
      cancer_prospective + icd10_cancer +  n_visits + n_hospital_fct + n_prescriptions +
      time_symptoms + time_followup + flulike_symptoms + upper_respiratory +
      lower_respiratory + gastrointestinal + distr_fct,
    data = df_validation %>% filter(excluded == "Included"),
    n.impute = 20,
    # n.impute = 100 in the true analysis, n.impute = 20 here for shorter computation
    nk = 6,
    tlinear = FALSE,
    x = TRUE,
    B = 200,
    group = df_validation %>%
      filter(excluded == "Included") %>%
      pull(outcome_fct)
  )
```

```
## Iteration 1 
Iteration 2 
Iteration 3 
Iteration 4 
Iteration 5 
Iteration 6 
Iteration 7 
Iteration 8 
Iteration 9 
Iteration 10 
Iteration 11 
Iteration 12 
Iteration 13 
Iteration 14 
Iteration 15 
Iteration 16 
Iteration 17 
Iteration 18 
Iteration 19 
Iteration 20 
Iteration 21 
Iteration 22 
Iteration 23
```

```
imputation_outcomes_validation
```

```
## 
## Multiple Imputation using Bootstrap and PMM
## 
## aregImpute(formula = ~outcome_fct + age + sex + bmi_prospective + 
##     smoking_current + +clinical_score_fct + htn_prospective + 
##     icd10_htn + antihypertensive + heart_prospective + icd10_heart_disease + 
##     diabetes_prospective + icd10_dm + diabetes_drugs + pulm_prospective + 
##     icd10_pulmonary_disease + ckd_prospective + ckd_retrospective + 
##     cancer_prospective + icd10_cancer + n_visits + n_hospital_fct + 
##     n_prescriptions + time_symptoms + time_followup + flulike_symptoms + 
##     upper_respiratory + lower_respiratory + gastrointestinal + 
##     distr_fct, data = df_validation %>% filter(excluded == "Included"), 
##     n.impute = 20, group = df_validation %>% filter(excluded == 
##         "Included") %>% pull(outcome_fct), nk = 6, tlinear = FALSE, 
##     x = TRUE, B = 200)
## 
## n: 3131  p: 30   Imputations: 20     nk: 6 
## 
## Number of NAs:
##             outcome_fct                     age                     sex 
##                       0                       0                       0 
##         bmi_prospective         smoking_current      clinical_score_fct 
##                     326                     158                     270 
##         htn_prospective               icd10_htn        antihypertensive 
##                      40                       0                       0 
##       heart_prospective     icd10_heart_disease    diabetes_prospective 
##                      40                       0                      40 
##                icd10_dm          diabetes_drugs        pulm_prospective 
##                       0                       0                      40 
## icd10_pulmonary_disease         ckd_prospective       ckd_retrospective 
##                       0                      40                       0 
##      cancer_prospective            icd10_cancer                n_visits 
##                      40                       0                       0 
##          n_hospital_fct         n_prescriptions           time_symptoms 
##                       0                       0                     146 
##           time_followup        flulike_symptoms       upper_respiratory 
##                      40                      40                      40 
##       lower_respiratory        gastrointestinal               distr_fct 
##                      40                      40                       0 
## 
##                         type d.f.
## outcome_fct                c    3
## age                        s    5
## sex                        l    1
## bmi_prospective            s    5
## smoking_current            l    1
## clinical_score_fct         l    1
## htn_prospective            l    1
## icd10_htn                  l    1
## antihypertensive           l    1
## heart_prospective          l    1
## icd10_heart_disease        l    1
## diabetes_prospective       l    1
## icd10_dm                   l    1
## diabetes_drugs             l    1
## pulm_prospective           l    1
## icd10_pulmonary_disease    l    1
## ckd_prospective            l    1
## ckd_retrospective          l    1
## cancer_prospective         l    1
## icd10_cancer               l    1
## n_visits                   s    5
## n_hospital_fct             l    1
## n_prescriptions            s    4
## time_symptoms              s    5
## time_followup              s    4
## flulike_symptoms           l    1
## upper_respiratory          l    1
## lower_respiratory          l    1
## gastrointestinal           l    1
## distr_fct                  l    1
## 
## R-squares for Predicting Non-Missing Values for Each Variable
## Using Last Imputations of Predictors
##      bmi_prospective      smoking_current   clinical_score_fct 
##                0.244                0.061                0.208 
##      htn_prospective    heart_prospective diabetes_prospective 
##                0.629                0.396                0.366 
##     pulm_prospective      ckd_prospective   cancer_prospective 
##                0.267                0.137                0.351 
##        time_symptoms        time_followup     flulike_symptoms 
##                0.285                0.299                0.251 
##    upper_respiratory    lower_respiratory     gastrointestinal 
##                0.136                0.167                0.131
```

```
par(mfrow=c(1,3))
plot(imputation_outcomes_validation)
```

```
par(mfrow=c(1,1))
```

### 6 Regression

Here we show (and compare) a complete case analysis to the multiple imputation analysis for both the derivation and validation cohorts. The multiply imputated models were included in the manuscript. Note: because we used 20 imputations in the code that is presented here (for computational reasons), instead of the 100 imputations that we used for analysis presented in the manuscript, the point-estimates of the beta-coefficients, and discrimination and calibration indices will be slightly different than is presented in the manuscript. ## Derivation cohort, complete case

```
dd_derivation <- datadist(df_derivation %>% filter(excluded == "Included"))
dd_derivation$limits$age <- c(45, 60, 75, 25, 88, 18, 102)
dd_derivation$limits$bmi_prospective <- c(25, 25, 35, 15, 44, 12, 56)
dd_derivation$limits$eGFR <- c(45, 90, 150, 30, 250, 10, 278)
options(datadist = "dd_derivation")

mod_cc_derivation <- lrm(
  outcome_fct ~ rcs(age, knots = c(21, 36, 51, 70)) + sex + bmi_prospective + htn_prospective + 
    heart_prospective + pulm_prospective + diabetes_prospective + cancer_prospective +
    smoking_current + flulike_symptoms + upper_respiratory + lower_respiratory +
    gastrointestinal + clinical_score_fct,
  data = df_derivation %>% filter(excluded == "Included")
)

mod_cc_derivation
```

```
## Frequencies of Missing Values Due to Each Variable
##          outcome_fct                  age                  sex 
##                    0                    0                    0 
##      bmi_prospective      htn_prospective    heart_prospective 
##                 1115                  185                  195 
##     pulm_prospective diabetes_prospective   cancer_prospective 
##                  199                  202                  203 
##      smoking_current     flulike_symptoms    upper_respiratory 
##                  246                   37                  129 
##    lower_respiratory     gastrointestinal   clinical_score_fct 
##                  186                  204                   36 
## 
## Logistic Regression Model
##  
##  lrm(formula = outcome_fct ~ rcs(age, knots = c(21, 36, 51, 70)) + 
##      sex + bmi_prospective + htn_prospective + heart_prospective + 
##      pulm_prospective + diabetes_prospective + cancer_prospective + 
##      smoking_current + flulike_symptoms + upper_respiratory + 
##      lower_respiratory + gastrointestinal + clinical_score_fct, 
##      data = df_derivation %>% filter(excluded == "Included"))
##  
##  
##  Frequencies of Responses
##  
##    0   1   2   3 
##  424  46  16   2 
##  
##  
##                        Model Likelihood     Discrimination    Rank Discrim.    
##                           Ratio Test           Indexes           Indexes       
##  Obs           488    LR chi2      97.44    R2       0.294    C       0.835    
##  max |deriv| 2e-05    d.f.            17    g        1.401    Dxy     0.670    
##                       Pr(> chi2) <0.0001    gr       4.059    gamma   0.671    
##                                             gp       0.050    tau-a   0.158    
##                                             Brier    0.031                     
##  
##                       Coef    S.E.   Wald Z Pr(>|Z|)
##  y>=1                 -3.5305 2.2713 -1.55  0.1201  
##  y>=2                 -5.2064 2.2835 -2.28  0.0226  
##  y>=3                 -7.5677 2.3847 -3.17  0.0015  
##  age                  -0.0267 0.0832 -0.32  0.7482  
##  age'                  0.5721 0.5206  1.10  0.2718  
##  age''                -1.7904 1.3526 -1.32  0.1856  
##  age'''                2.8096 1.6703  1.68  0.0926  
##  sex                  -0.0273 0.3144 -0.09  0.9307  
##  bmi_prospective      -0.0056 0.0294 -0.19  0.8501  
##  htn_prospective       0.2495 0.4447  0.56  0.5748  
##  heart_prospective    -0.5288 0.5998 -0.88  0.3779  
##  pulm_prospective     -0.0837 0.4774 -0.18  0.8609  
##  diabetes_prospective  0.2287 0.7014  0.33  0.7444  
##  cancer_prospective   -0.4272 0.8156 -0.52  0.6005  
##  smoking_current       0.1388 0.5923  0.23  0.8147  
##  flulike_symptoms      0.7032 0.8378  0.84  0.4013  
##  upper_respiratory    -0.1720 0.3889 -0.44  0.6583  
##  lower_respiratory     0.7090 0.3438  2.06  0.0392  
##  gastrointestinal      0.6447 0.3293  1.96  0.0502  
##  clinical_score_fct    2.0566 0.3353  6.13  <0.0001 
##
```

```
knitr::kable(summary(mod_cc_derivation))
```

|  | Low | High | Diff. | Effect | S.E. | Lower 0.95 | Upper 0.95 | Type |
| --- | --- | --- | --- | --- | --- | --- | --- | --- |
| age | 45 | 75 | 30 | 0.6769165 | 0.6673952 | -0.6311541 | 1.9849872 | 1 |
| Odds Ratio | 45 | 75 | 30 | 1.9678007 | NA | 0.5319775 | 7.2789539 | 2 |
| sex | 0 | 1 | 1 | -0.0273284 | 0.3143744 | -0.6434910 | 0.5888342 | 1 |
| Odds Ratio | 0 | 1 | 1 | 0.9730417 | NA | 0.5254549 | 1.8018866 | 2 |
| bmi\_prospective | 25 | 35 | 10 | -0.0556376 | 0.2943571 | -0.6325670 | 0.5212917 | 1 |
| Odds Ratio | 25 | 35 | 10 | 0.9458818 | NA | 0.5312264 | 1.6842018 | 2 |
| htn\_prospective | 0 | 1 | 1 | 0.2495160 | 0.4447483 | -0.6221748 | 1.1212067 | 1 |
| Odds Ratio | 0 | 1 | 1 | 1.2834041 | NA | 0.5367758 | 3.0685548 | 2 |
| heart\_prospective | 0 | 1 | 1 | -0.5288359 | 0.5997694 | -1.7043624 | 0.6466905 | 1 |
| Odds Ratio | 0 | 1 | 1 | 0.5892905 | NA | 0.1818883 | 1.9092119 | 2 |
| pulm\_prospective | 0 | 1 | 1 | -0.0836631 | 0.4774289 | -1.0194067 | 0.8520804 | 1 |
| Odds Ratio | 0 | 1 | 1 | 0.9197410 | NA | 0.3608090 | 2.3445193 | 2 |
| diabetes\_prospective | 0 | 1 | 1 | 0.2287086 | 0.7013633 | -1.1459381 | 1.6033553 | 1 |
| Odds Ratio | 0 | 1 | 1 | 1.2569757 | NA | 0.3179255 | 4.9696794 | 2 |
| cancer\_prospective | 0 | 1 | 1 | -0.4271837 | 0.8156249 | -2.0257791 | 1.1714118 | 1 |
| Odds Ratio | 0 | 1 | 1 | 0.6523437 | NA | 0.1318910 | 3.2265446 | 2 |
| smoking\_current | 0 | 1 | 1 | 0.1388103 | 0.5923195 | -1.0221146 | 1.2997353 | 1 |
| Odds Ratio | 0 | 1 | 1 | 1.1489061 | NA | 0.3598332 | 3.6683254 | 2 |
| flulike\_symptoms | 0 | 1 | 1 | 0.7032276 | 0.8378016 | -0.9388334 | 2.3452885 | 1 |
| Odds Ratio | 0 | 1 | 1 | 2.0202627 | NA | 0.3910838 | 10.4362832 | 2 |
| upper\_respiratory | 0 | 1 | 1 | -0.1719684 | 0.3888611 | -0.9341222 | 0.5901854 | 1 |
| Odds Ratio | 0 | 1 | 1 | 0.8420058 | NA | 0.3929306 | 1.8043228 | 2 |
| lower\_respiratory | 0 | 1 | 1 | 0.7090044 | 0.3438488 | 0.0350732 | 1.3829356 | 1 |
| Odds Ratio | 0 | 1 | 1 | 2.0319672 | NA | 1.0356955 | 3.9865873 | 2 |
| gastrointestinal | 0 | 1 | 1 | 0.6447378 | 0.3292705 | -0.0006205 | 1.2900961 | 1 |
| Odds Ratio | 0 | 1 | 1 | 1.9054873 | NA | 0.9993797 | 3.6331356 | 2 |
| clinical\_score\_fct | 0 | 1 | 1 | 2.0565989 | 0.3353390 | 1.3993465 | 2.7138513 | 1 |
| Odds Ratio | 0 | 1 | 1 | 7.8193303 | NA | 4.0525508 | 15.0872696 | 2 |

```
plot(summary(mod_cc_derivation))

ggplot(Predict(mod_cc_derivation))
```

```
plot(nomogram(mod_cc_derivation))
```

#### 6.1 Derivation cohort, multiple imputation

```
mod_multi_derivation <- fit.mult.impute(
  outcome_fct ~ rcs(age, knots = c(21, 36, 51, 70)) + sex + bmi_prospective + htn_prospective + 
    heart_prospective + pulm_prospective + diabetes_prospective + cancer_prospective +
    smoking_current + flulike_symptoms + upper_respiratory + lower_respiratory +
    gastrointestinal + clinical_score_fct,
  lrm,
  imputation_outcomes_derivation,
  data = df_derivation %>% filter(excluded == "Included"))
```

```
## 
## Variance Inflation Factors Due to Imputation:
## 
##                 y>=1                 y>=2                 y>=3 
##                 1.15                 1.15                 1.14 
##                  age                 age'                age'' 
##                 1.03                 1.01                 1.01 
##               age'''                  sex      bmi_prospective 
##                 1.02                 1.06                 3.94 
##      htn_prospective    heart_prospective     pulm_prospective 
##                 1.18                 1.11                 1.13 
## diabetes_prospective   cancer_prospective      smoking_current 
##                 1.08                 1.11                 1.25 
##     flulike_symptoms    upper_respiratory    lower_respiratory 
##                 1.09                 1.13                 1.11 
##     gastrointestinal   clinical_score_fct 
##                 1.19                 1.16 
## 
## Rate of Missing Information:
## 
##                 y>=1                 y>=2                 y>=3 
##                 0.13                 0.13                 0.12 
##                  age                 age'                age'' 
##                 0.02                 0.01                 0.01 
##               age'''                  sex      bmi_prospective 
##                 0.02                 0.06                 0.75 
##      htn_prospective    heart_prospective     pulm_prospective 
##                 0.16                 0.10                 0.12 
## diabetes_prospective   cancer_prospective      smoking_current 
##                 0.07                 0.10                 0.20 
##     flulike_symptoms    upper_respiratory    lower_respiratory 
##                 0.08                 0.12                 0.10 
##     gastrointestinal   clinical_score_fct 
##                 0.16                 0.14 
## 
## d.f. for t-distribution for Tests of Single Coefficients:
## 
##                 y>=1                 y>=2                 y>=3 
##              1160.79              1180.88              1217.85 
##                  age                 age'                age'' 
##             31065.24            375614.60            218300.08 
##               age'''                  sex      bmi_prospective 
##             49646.12              5689.50                34.13 
##      htn_prospective    heart_prospective     pulm_prospective 
##               782.06              1950.80              1361.75 
## diabetes_prospective   cancer_prospective      smoking_current 
##              3410.59              2077.85               483.77 
##     flulike_symptoms    upper_respiratory    lower_respiratory 
##              2640.44              1416.66              1798.49 
##     gastrointestinal   clinical_score_fct 
##               717.25               987.53 
## 
## The following fit components were averaged over the 20 model fits:
## 
##   stats linear.predictors
```

```
mod_multi_derivation
```

```
## Logistic Regression Model
##  
##  fit.mult.impute(formula = outcome_fct ~ rcs(age, knots = c(21, 
##      36, 51, 70)) + sex + bmi_prospective + htn_prospective + 
##      heart_prospective + pulm_prospective + diabetes_prospective + 
##      cancer_prospective + smoking_current + flulike_symptoms + 
##      upper_respiratory + lower_respiratory + gastrointestinal + 
##      clinical_score_fct, fitter = lrm, xtrans = imputation_outcomes_derivation, 
##      data = df_derivation %>% filter(excluded == "Included"))
##  
##  
##  Frequencies of Responses
##  
##     0    1    2    3 
##  1363  162   71   29 
##  
##                        Model Likelihood     Discrimination    Rank Discrim.    
##                           Ratio Test           Indexes           Indexes       
##  Obs          1625    LR chi2     324.75    R2       0.262    C       0.798    
##  max |deriv| 1e-06    d.f.            17    g        1.267    Dxy     0.596    
##                       Pr(> chi2) <0.0001    gr       3.549    gamma   0.597    
##                                             gp       0.077    tau-a   0.170    
##                                             Brier    0.047                     
##  
##                       Coef    S.E.   Wald Z Pr(>|Z|)
##  y>=1                 -4.0131 1.3724 -2.92  0.0035  
##  y>=2                 -5.3547 1.3757 -3.89  <0.0001 
##  y>=3                 -6.8351 1.3840 -4.94  <0.0001 
##  age                   0.0633 0.0466  1.36  0.1744  
##  age'                 -0.0802 0.2707 -0.30  0.7671  
##  age''                -0.0137 0.6924 -0.02  0.9843  
##  age'''                0.6489 0.8300  0.78  0.4343  
##  sex                  -0.1542 0.1594 -0.97  0.3333  
##  bmi_prospective      -0.0119 0.0296 -0.40  0.6871  
##  htn_prospective      -0.0300 0.2275 -0.13  0.8950  
##  heart_prospective     0.0035 0.2675  0.01  0.9897  
##  pulm_prospective      0.1852 0.2363  0.78  0.4331  
##  diabetes_prospective  0.6950 0.3514  1.98  0.0479  
##  cancer_prospective   -0.4514 0.3541 -1.27  0.2025  
##  smoking_current      -0.1570 0.3583 -0.44  0.6614  
##  flulike_symptoms     -0.3911 0.3221 -1.21  0.2247  
##  upper_respiratory    -0.2897 0.1880 -1.54  0.1234  
##  lower_respiratory     0.3074 0.1721  1.79  0.0742  
##  gastrointestinal      0.5211 0.1737  3.00  0.0027  
##  clinical_score_fct    1.8577 0.1748 10.63  <0.0001 
##
```

```
knitr::kable(summary(mod_multi_derivation))
```

|  | Low | High | Diff. | Effect | S.E. | Lower 0.95 | Upper 0.95 | Type |
| --- | --- | --- | --- | --- | --- | --- | --- | --- |
| age | 45 | 75 | 30 | 1.3561767 | 0.2983171 | 0.7714859 | 1.9408674 | 1 |
| Odds Ratio | 45 | 75 | 30 | 3.8813252 | NA | 2.1629779 | 6.9647895 | 2 |
| sex | 0 | 1 | 1 | -0.1541806 | 0.1593779 | -0.4665555 | 0.1581943 | 1 |
| Odds Ratio | 0 | 1 | 1 | 0.8571172 | NA | 0.6271588 | 1.1713938 | 2 |
| bmi\_prospective | 25 | 35 | 10 | -0.1191532 | 0.2958536 | -0.6990156 | 0.4607092 | 1 |
| Odds Ratio | 25 | 35 | 10 | 0.8876718 | NA | 0.4970744 | 1.5851978 | 2 |
| htn\_prospective | 0 | 1 | 1 | -0.0300305 | 0.2275300 | -0.4759812 | 0.4159201 | 1 |
| Odds Ratio | 0 | 1 | 1 | 0.9704159 | NA | 0.6212752 | 1.5157648 | 2 |
| heart\_prospective | 0 | 1 | 1 | 0.0034554 | 0.2674531 | -0.5207430 | 0.5276538 | 1 |
| Odds Ratio | 0 | 1 | 1 | 1.0034614 | NA | 0.5940790 | 1.6949509 | 2 |
| pulm\_prospective | 0 | 1 | 1 | 0.1852259 | 0.2362746 | -0.2778638 | 0.6483157 | 1 |
| Odds Ratio | 0 | 1 | 1 | 1.2034903 | NA | 0.7574000 | 1.9123171 | 2 |
| diabetes\_prospective | 0 | 1 | 1 | 0.6950074 | 0.3513757 | 0.0063237 | 1.3836910 | 1 |
| Odds Ratio | 0 | 1 | 1 | 2.0037238 | NA | 1.0063438 | 3.9896000 | 2 |
| cancer\_prospective | 0 | 1 | 1 | -0.4513596 | 0.3541437 | -1.1454684 | 0.2427492 | 1 |
| Odds Ratio | 0 | 1 | 1 | 0.6367618 | NA | 0.3180749 | 1.2747489 | 2 |
| smoking\_current | 0 | 1 | 1 | -0.1569522 | 0.3583486 | -0.8593025 | 0.5453981 | 1 |
| Odds Ratio | 0 | 1 | 1 | 0.8547449 | NA | 0.4234573 | 1.7252951 | 2 |
| flulike\_symptoms | 0 | 1 | 1 | -0.3911186 | 0.3221243 | -1.0224706 | 0.2402334 | 1 |
| Odds Ratio | 0 | 1 | 1 | 0.6763000 | NA | 0.3597052 | 1.2715459 | 2 |
| upper\_respiratory | 0 | 1 | 1 | -0.2896720 | 0.1880036 | -0.6581524 | 0.0788083 | 1 |
| Odds Ratio | 0 | 1 | 1 | 0.7485090 | NA | 0.5178072 | 1.0819969 | 2 |
| lower\_respiratory | 0 | 1 | 1 | 0.3073512 | 0.1721232 | -0.0300040 | 0.6447064 | 1 |
| Odds Ratio | 0 | 1 | 1 | 1.3598184 | NA | 0.9704417 | 1.9054275 | 2 |
| gastrointestinal | 0 | 1 | 1 | 0.5210712 | 0.1737077 | 0.1806103 | 0.8615321 | 1 |
| Odds Ratio | 0 | 1 | 1 | 1.6838304 | NA | 1.1979482 | 2.3667841 | 2 |
| clinical\_score\_fct | 0 | 1 | 1 | 1.8577451 | 0.1748408 | 1.5150634 | 2.2004268 | 1 |
| Odds Ratio | 0 | 1 | 1 | 6.4092683 | NA | 4.5497096 | 9.0288665 | 2 |

```
plot(summary(mod_multi_derivation))
```

```
ggplot(Predict(mod_multi_derivation))
```

```
plot(nomogram(mod_multi_derivation))
```

#### 6.2 Validation cohort, complete case

```
dd_validation <- datadist(df_validation %>% filter(excluded == "Included"))
dd_validation$limits$age <- c(45, 60, 75, 25, 88, 18, 102)
dd_validation$limits$bmi_prospective <- c(25, 25, 35, 15, 44, 12, 56)
dd_validation$limits$eGFR <- c(45, 90, 150, 30, 250, 10, 278)
options(datadist = "dd_validation")

mod_cc_validation <- lrm(
  outcome_fct ~ rcs(age, 4) + sex + bmi_prospective + htn_prospective + heart_prospective +
    pulm_prospective + diabetes_prospective  + cancer_prospective +
    smoking_current + flulike_symptoms + upper_respiratory + lower_respiratory +
    gastrointestinal + clinical_score_fct,
  data = df_validation %>% filter(excluded == "Included")
)

mod_cc_validation
```

```
## Frequencies of Missing Values Due to Each Variable
##          outcome_fct                  age                  sex 
##                    0                    0                    0 
##      bmi_prospective      htn_prospective    heart_prospective 
##                  326                   40                   40 
##     pulm_prospective diabetes_prospective   cancer_prospective 
##                   40                   40                   40 
##      smoking_current     flulike_symptoms    upper_respiratory 
##                  158                   40                   40 
##    lower_respiratory     gastrointestinal   clinical_score_fct 
##                   40                   40                  270 
## 
## Logistic Regression Model
##  
##  lrm(formula = outcome_fct ~ rcs(age, 4) + sex + bmi_prospective + 
##      htn_prospective + heart_prospective + pulm_prospective + 
##      diabetes_prospective + cancer_prospective + smoking_current + 
##      flulike_symptoms + upper_respiratory + lower_respiratory + 
##      gastrointestinal + clinical_score_fct, data = df_validation %>% 
##      filter(excluded == "Included"))
##  
##  
##  Frequencies of Responses
##  
##     0    1    2    3 
##  2323  184   88   15 
##  
##  
##                        Model Likelihood     Discrimination    Rank Discrim.    
##                           Ratio Test           Indexes           Indexes       
##  Obs          2610    LR chi2     329.24    R2       0.204    C       0.779    
##  max |deriv| 3e-11    d.f.            16    g        1.149    Dxy     0.557    
##                       Pr(> chi2) <0.0001    gr       3.156    gamma   0.559    
##                                             gp       0.048    tau-a   0.112    
##                                             Brier    0.032                     
##  
##                       Coef     S.E.   Wald Z Pr(>|Z|)
##  y>=1                  -7.2288 1.0320 -7.00  <0.0001 
##  y>=2                  -8.5135 1.0393 -8.19  <0.0001 
##  y>=3                 -10.6998 1.0749 -9.95  <0.0001 
##  age                    0.0804 0.0366  2.20  0.0282  
##  age'                  -0.3898 0.1830 -2.13  0.0331  
##  age''                  0.7920 0.3312  2.39  0.0168  
##  sex                   -0.1559 0.1377 -1.13  0.2575  
##  bmi_prospective        0.0527 0.0134  3.94  <0.0001 
##  htn_prospective        0.0727 0.1925  0.38  0.7055  
##  heart_prospective      0.0172 0.2429  0.07  0.9435  
##  pulm_prospective       0.9308 0.2360  3.94  <0.0001 
##  diabetes_prospective   0.3508 0.3233  1.09  0.2778  
##  cancer_prospective     0.3250 0.3518  0.92  0.3556  
##  smoking_current        0.1149 0.2293  0.50  0.6163  
##  flulike_symptoms       0.9227 0.2187  4.22  <0.0001 
##  upper_respiratory      0.0596 0.1406  0.42  0.6715  
##  lower_respiratory      0.3579 0.1597  2.24  0.0250  
##  gastrointestinal       0.2522 0.1615  1.56  0.1183  
##  clinical_score_fct     1.1961 0.1997  5.99  <0.0001 
##
```

```
knitr::kable(summary(mod_cc_validation))
```

|  | Low | High | Diff. | Effect | S.E. | Lower 0.95 | Upper 0.95 | Type |
| --- | --- | --- | --- | --- | --- | --- | --- | --- |
| age | 45 | 75 | 30 | 1.8166809 | 0.2171132 | 1.3911468 | 2.2422149 | 1 |
| Odds Ratio | 45 | 75 | 30 | 6.1514072 | NA | 4.0194570 | 9.4141599 | 2 |
| sex | 0 | 1 | 1 | -0.1559479 | 0.1377302 | -0.4258941 | 0.1139984 | 1 |
| Odds Ratio | 0 | 1 | 1 | 0.8556038 | NA | 0.6531855 | 1.1207503 | 2 |
| bmi\_prospective | 25 | 35 | 10 | 0.5270546 | 0.1338406 | 0.2647319 | 0.7893772 | 1 |
| Odds Ratio | 25 | 35 | 10 | 1.6939356 | NA | 1.3030816 | 2.2020247 | 2 |
| htn\_prospective | 0 | 1 | 1 | 0.0727315 | 0.1924860 | -0.3045342 | 0.4499971 | 1 |
| Odds Ratio | 0 | 1 | 1 | 1.0754417 | NA | 0.7374668 | 1.5683077 | 2 |
| heart\_prospective | 0 | 1 | 1 | 0.0172152 | 0.2428872 | -0.4588351 | 0.4932654 | 1 |
| Odds Ratio | 0 | 1 | 1 | 1.0173642 | NA | 0.6320195 | 1.6376551 | 2 |
| pulm\_prospective | 0 | 1 | 1 | 0.9307947 | 0.2359707 | 0.4683006 | 1.3932887 | 1 |
| Odds Ratio | 0 | 1 | 1 | 2.5365241 | NA | 1.5972775 | 4.0280756 | 2 |
| diabetes\_prospective | 0 | 1 | 1 | 0.3508364 | 0.3232891 | -0.2827986 | 0.9844714 | 1 |
| Odds Ratio | 0 | 1 | 1 | 1.4202549 | NA | 0.7536715 | 2.6763968 | 2 |
| cancer\_prospective | 0 | 1 | 1 | 0.3249767 | 0.3518187 | -0.3645753 | 1.0145287 | 1 |
| Odds Ratio | 0 | 1 | 1 | 1.3839984 | NA | 0.6944915 | 2.7580633 | 2 |
| smoking\_current | 0 | 1 | 1 | 0.1149151 | 0.2293367 | -0.3345766 | 0.5644068 | 1 |
| Odds Ratio | 0 | 1 | 1 | 1.1217782 | NA | 0.7156410 | 1.7584043 | 2 |
| flulike\_symptoms | 0 | 1 | 1 | 0.9227289 | 0.2186510 | 0.4941807 | 1.3512770 | 1 |
| Odds Ratio | 0 | 1 | 1 | 2.5161472 | NA | 1.6391548 | 3.8623546 | 2 |
| upper\_respiratory | 0 | 1 | 1 | 0.0596450 | 0.1406287 | -0.2159822 | 0.3352721 | 1 |
| Odds Ratio | 0 | 1 | 1 | 1.0614596 | NA | 0.8057497 | 1.3983209 | 2 |
| lower\_respiratory | 0 | 1 | 1 | 0.3579079 | 0.1597066 | 0.0448888 | 0.6709270 | 1 |
| Odds Ratio | 0 | 1 | 1 | 1.4303338 | NA | 1.0459115 | 1.9560497 | 2 |
| gastrointestinal | 0 | 1 | 1 | 0.2522338 | 0.1614931 | -0.0642869 | 0.5687546 | 1 |
| Odds Ratio | 0 | 1 | 1 | 1.2868969 | NA | 0.9377359 | 1.7660662 | 2 |
| clinical\_score\_fct | 0 | 1 | 1 | 1.1961454 | 0.1997478 | 0.8046468 | 1.5876439 | 1 |
| Odds Ratio | 0 | 1 | 1 | 3.3073438 | NA | 2.2359067 | 4.8922090 | 2 |

```
plot(summary(mod_cc_validation))
```

```
ggplot(Predict(mod_cc_validation))
```

```
plot(nomogram(mod_cc_validation))
```

#### 6.3 Validation cohort, multiple imputation

```
mod_multi_validation <- fit.mult.impute(
  outcome_fct ~ rcs(age, 4) + sex + bmi_prospective + htn_prospective + heart_prospective +
    pulm_prospective + diabetes_prospective  + cancer_prospective +
    smoking_current + flulike_symptoms + upper_respiratory + lower_respiratory +
    gastrointestinal + clinical_score_fct,
  lrm,
  imputation_outcomes_validation,
  data = df_validation %>% filter(excluded == "Included"))
```

```
## 
## Variance Inflation Factors Due to Imputation:
## 
##                 y>=1                 y>=2                 y>=3 
##                 1.01                 1.01                 1.01 
##                  age                 age'                age'' 
##                 1.01                 1.01                 1.01 
##                  sex      bmi_prospective      htn_prospective 
##                 1.01                 1.12                 1.02 
##    heart_prospective     pulm_prospective diabetes_prospective 
##                 1.03                 1.04                 1.01 
##   cancer_prospective      smoking_current     flulike_symptoms 
##                 1.02                 1.06                 1.03 
##    upper_respiratory    lower_respiratory     gastrointestinal 
##                 1.02                 1.03                 1.03 
##   clinical_score_fct 
##                 1.14 
## 
## Rate of Missing Information:
## 
##                 y>=1                 y>=2                 y>=3 
##                 0.01                 0.01                 0.01 
##                  age                 age'                age'' 
##                 0.01                 0.01                 0.01 
##                  sex      bmi_prospective      htn_prospective 
##                 0.01                 0.11                 0.02 
##    heart_prospective     pulm_prospective diabetes_prospective 
##                 0.03                 0.03                 0.01 
##   cancer_prospective      smoking_current     flulike_symptoms 
##                 0.01                 0.06                 0.03 
##    upper_respiratory    lower_respiratory     gastrointestinal 
##                 0.02                 0.03                 0.03 
##   clinical_score_fct 
##                 0.12 
## 
## d.f. for t-distribution for Tests of Single Coefficients:
## 
##                 y>=1                 y>=2                 y>=3 
##            236444.95            236854.22            239497.89 
##                  age                 age'                age'' 
##            718977.42            234338.99            151226.62 
##                  sex      bmi_prospective      htn_prospective 
##            206595.09              1552.02             43919.09 
##    heart_prospective     pulm_prospective diabetes_prospective 
##             19650.42             15843.28            167637.50 
##   cancer_prospective      smoking_current     flulike_symptoms 
##             84524.76              6004.68             27512.57 
##    upper_respiratory    lower_respiratory     gastrointestinal 
##             48099.49             19523.72             29322.72 
##   clinical_score_fct 
##              1330.74 
## 
## The following fit components were averaged over the 20 model fits:
## 
##   stats linear.predictors
```

```
mod_multi_validation
```

```
## Logistic Regression Model
##  
##  fit.mult.impute(formula = outcome_fct ~ rcs(age, 4) + sex + bmi_prospective + 
##      htn_prospective + heart_prospective + pulm_prospective + 
##      diabetes_prospective + cancer_prospective + smoking_current + 
##      flulike_symptoms + upper_respiratory + lower_respiratory + 
##      gastrointestinal + clinical_score_fct, fitter = lrm, xtrans = imputation_outcomes_validation, 
##      data = df_validation %>% filter(excluded == "Included"))
##  
##  
##  Frequencies of Responses
##  
##     0    1    2    3 
##  2780  213  117   21 
##  
##                        Model Likelihood     Discrimination    Rank Discrim.    
##                           Ratio Test           Indexes           Indexes       
##  Obs          3131    LR chi2     477.32    R2       0.240    C       0.800    
##  max |deriv| 7e-07    d.f.            16    g        1.185    Dxy     0.600    
##                       Pr(> chi2) <0.0001    gr       3.270    gamma   0.601    
##                                             gp       0.056    tau-a   0.123    
##                                             Brier    0.033                     
##  
##                       Coef     S.E.   Wald Z Pr(>|Z|)
##  y>=1                  -7.1319 0.9572  -7.45 <0.0001 
##  y>=2                  -8.3767 0.9635  -8.69 <0.0001 
##  y>=3                 -10.6596 0.9961 -10.70 <0.0001 
##  age                    0.0872 0.0339   2.57 0.0102  
##  age'                  -0.4311 0.1688  -2.55 0.0107  
##  age''                  0.8771 0.3049   2.88 0.0040  
##  sex                   -0.2285 0.1271  -1.80 0.0722  
##  bmi_prospective        0.0484 0.0129   3.74 0.0002  
##  htn_prospective       -0.0912 0.1798  -0.51 0.6119  
##  heart_prospective     -0.0037 0.2245  -0.02 0.9868  
##  pulm_prospective       0.9430 0.2208   4.27 <0.0001 
##  diabetes_prospective   0.3673 0.2894   1.27 0.2044  
##  cancer_prospective     0.4782 0.3223   1.48 0.1379  
##  smoking_current        0.1334 0.2130   0.63 0.5311  
##  flulike_symptoms       0.8090 0.1922   4.21 <0.0001 
##  upper_respiratory     -0.0291 0.1298  -0.22 0.8224  
##  lower_respiratory      0.4034 0.1499   2.69 0.0071  
##  gastrointestinal       0.2514 0.1527   1.65 0.0996  
##  clinical_score_fct     1.3425 0.1891   7.10 <0.0001 
##
```

```
knitr::kable(summary(mod_multi_validation))
```

|  | Low | High | Diff. | Effect | S.E. | Lower 0.95 | Upper 0.95 | Type |
| --- | --- | --- | --- | --- | --- | --- | --- | --- |
| age | 45 | 75 | 30 | 1.9837746 | 0.1930094 | 1.6054832 | 2.3620661 | 1 |
| Odds Ratio | 45 | 75 | 30 | 7.2701333 | NA | 4.9802653 | 10.6128558 | 2 |
| sex | 0 | 1 | 1 | -0.2285461 | 0.1271269 | -0.4777104 | 0.0206181 | 1 |
| Odds Ratio | 0 | 1 | 1 | 0.7956896 | NA | 0.6202018 | 1.0208321 | 2 |
| bmi\_prospective | 25 | 35 | 10 | 0.4839121 | 0.1294882 | 0.2301199 | 0.7377044 | 1 |
| Odds Ratio | 25 | 35 | 10 | 1.6224091 | NA | 1.2587509 | 2.0911297 | 2 |
| htn\_prospective | 0 | 1 | 1 | -0.0912167 | 0.1797691 | -0.4435577 | 0.2611243 | 1 |
| Odds Ratio | 0 | 1 | 1 | 0.9128199 | NA | 0.6417492 | 1.2983890 | 2 |
| heart\_prospective | 0 | 1 | 1 | -0.0037183 | 0.2245438 | -0.4438161 | 0.4363794 | 1 |
| Odds Ratio | 0 | 1 | 1 | 0.9962886 | NA | 0.6415834 | 1.5470957 | 2 |
| pulm\_prospective | 0 | 1 | 1 | 0.9429572 | 0.2208029 | 0.5101915 | 1.3757230 | 1 |
| Odds Ratio | 0 | 1 | 1 | 2.5675631 | NA | 1.6656101 | 3.9579372 | 2 |
| diabetes\_prospective | 0 | 1 | 1 | 0.3672834 | 0.2894118 | -0.1999533 | 0.9345201 | 1 |
| Odds Ratio | 0 | 1 | 1 | 1.4438070 | NA | 0.8187690 | 2.5459913 | 2 |
| cancer\_prospective | 0 | 1 | 1 | 0.4782180 | 0.3223003 | -0.1534790 | 1.1099151 | 1 |
| Odds Ratio | 0 | 1 | 1 | 1.6131972 | NA | 0.8577188 | 3.0341007 | 2 |
| smoking\_current | 0 | 1 | 1 | 0.1334080 | 0.2129896 | -0.2840440 | 0.5508600 | 1 |
| Odds Ratio | 0 | 1 | 1 | 1.1427161 | NA | 0.7527335 | 1.7347443 | 2 |
| flulike\_symptoms | 0 | 1 | 1 | 0.8089821 | 0.1921539 | 0.4323674 | 1.1855968 | 1 |
| Odds Ratio | 0 | 1 | 1 | 2.2456210 | NA | 1.5409011 | 3.2726394 | 2 |
| upper\_respiratory | 0 | 1 | 1 | -0.0291263 | 0.1297760 | -0.2834825 | 0.2252299 | 1 |
| Odds Ratio | 0 | 1 | 1 | 0.9712937 | NA | 0.7531563 | 1.2526106 | 2 |
| lower\_respiratory | 0 | 1 | 1 | 0.4033933 | 0.1498784 | 0.1096371 | 0.6971495 | 1 |
| Odds Ratio | 0 | 1 | 1 | 1.4968955 | NA | 1.1158731 | 2.0080207 | 2 |
| gastrointestinal | 0 | 1 | 1 | 0.2514349 | 0.1526945 | -0.0478409 | 0.5507107 | 1 |
| Odds Ratio | 0 | 1 | 1 | 1.2858692 | NA | 0.9532854 | 1.7344853 | 2 |
| clinical\_score\_fct | 0 | 1 | 1 | 1.3425161 | 0.1890711 | 0.9719435 | 1.7130887 | 1 |
| Odds Ratio | 0 | 1 | 1 | 3.8286649 | NA | 2.6430764 | 5.5460654 | 2 |

```
plot(summary(mod_multi_validation))
```

```
ggplot(Predict(mod_multi_validation))
```

```
plot(nomogram(mod_multi_validation))
```

#### 6.4 Validation dataframes

The dataframes used internal and external validation presented in the manuscript are from a single randomly selected imputed dataframe. First we show the creation of the dataframe for the derivation cohort.

```
df_val_derivation <-
  impute.transcan(
    imputation_outcomes_derivation,
    data = df_derivation,
    # in the analysis presented in the manuscript 100 imputed datasets were constructed
    # but in this version, they were only 20. The code below is sample(x = 1:100, size = 1)
    # in the analysis presented in the manuscript
    imputation = sample(x = 1:20, size = 1),
    list.out = TRUE
  )
```

```
## 
## 
## Imputed missing values with the following frequencies
##  and stored them in variables with their original names:
## 
##      bmi_prospective      smoking_current   clinical_score_fct 
##                 1115                  246                   36 
##      htn_prospective    heart_prospective diabetes_prospective 
##                  185                  195                  202 
##     pulm_prospective      ckd_prospective   cancer_prospective 
##                  199                  206                  203 
##        time_symptoms        time_followup     flulike_symptoms 
##                   54                   11                   37 
##    upper_respiratory    lower_respiratory     gastrointestinal 
##                  129                  186                  204
```

```
df_val_derivation <- as.data.frame(df_val_derivation)
```

and then we show the same for the validation cohort.

```
df_val_validation <-
  impute.transcan(
    imputation_outcomes_validation,
    data = df_validation,
    # in the analysis presented in the manuscript 100 imputed datasets were constructed
    # but in this version, they were only 20. The code below is sample(x = 1:100, size = 1)
    # in the analysis presented in the manuscript
    imputation = sample(x = 1:20, size = 1),
    list.out = TRUE
  )
```

```
## 
## 
## Imputed missing values with the following frequencies
##  and stored them in variables with their original names:
## 
##      bmi_prospective      smoking_current   clinical_score_fct 
##                  326                  158                  270 
##      htn_prospective    heart_prospective diabetes_prospective 
##                   40                   40                   40 
##     pulm_prospective      ckd_prospective   cancer_prospective 
##                   40                   40                   40 
##        time_symptoms        time_followup     flulike_symptoms 
##                  146                   40                   40 
##    upper_respiratory    lower_respiratory     gastrointestinal 
##                   40                   40                   40
```

```
df_val_validation <- as.data.frame(df_val_validation)
```

### 7 Internal validation: calibration plot

First we add the predicted probabilities for each of the included outcomes to the singly imputed dataframe and then produce variables that represent the ordinal outcome or worse.

```
df_val_derivation <- df_val_derivation %>%
  cbind.data.frame(
    predict(
      mod_multi_derivation,
      df_val_derivation,
      type = "fitted")
  ) %>%
  mutate(outcome = as.numeric(as.character(outcome_fct))) %>%
  mutate(
    outpatient_fct = case_when(
      outcome >= 1 ~ 1,
      outcome < 1 ~ 0
    ),
    inpatient_fct = case_when(
      outcome >= 2 ~ 1,
      outcome < 2 ~ 0
    ),
    icu_death_fct = case_when(
      outcome >= 3 ~ 1,
      outcome < 3 ~ 0
    ))
```

Here we use a modification of the val.prob.ci.2() function to capture the confidence intervals for use in producing the figure. See also the modified function definitions in a separate .R file. Note: in the analysis presented in the manuscript, the confidence intervals are constructed using 2000 bootstrap resamples. Here we only do 200 resamples to save computational time. This figure shows the internal calibration for the outcome of outpatient visit or worse.

```
cal_outpatient <- val.prob.ci.2.modified(
  p = df_val_derivation$`y>=1`,
  y = df_val_derivation$outpatient_fct,
  logit = "logit",
  # in the analysis presented in the manuscript the confidence intervals are based on
  # 2000 bootstrap resamples. Here we have modified the function to do only do 200
  CL.BT = TRUE
)
```

```
## Bootstrap samples are being generated.
## 
## 
## 
## 
##  A 95% confidence interval is given for the calibration intercept, calibration slope and c-statistic.
```

```
plot_calibrated_1 <-
  df_val_derivation %>%
  mutate(
    bin = ntile(`y>=1`, n = 10),
    Linear = "Linear calibration",
    LOESS = "LOESS calibration"
  ) %>%
  group_by(bin) %>%
  mutate(
    n = n(),
    # Get ests and CIs
    bin_pred = mean(`y>=1`),
    bin_prob = mean(as.numeric(outpatient_fct)),
    se = sqrt((bin_prob * (1 - bin_prob)) / n),
    ul = bin_prob + 1.96 * se,
    ll = bin_prob - 1.96 * se
  ) %>%
  ungroup() %>%
  ggplot(aes(x = bin_pred, y = bin_prob)) +
  geom_abline(lty = 2) +
  geom_ribbon(
    data = as.data.frame(t(cal_outpatient)),
    aes(
      x = as.numeric(rownames((t(cal_outpatient)))),
      y = NULL,
      ymin = t(cal_outpatient)[, 1],
      ymax = t(cal_outpatient)[, 2]
    ),
    alpha = .3,
    fill = "#BC3C29FF"
  ) +
  geom_smooth(
    aes(x = `y>=1`, y = as.numeric(outpatient_fct), color = LOESS), 
    se = FALSE, method = "loess"
  ) +
  geom_pointrange(aes(ymin = ll, ymax = ul), size = 0.3, color = "black") +
  scale_color_manual(values = c("#BC3C29FF")) +
  scale_y_continuous(breaks = seq(0, 1, by = 0.2)) +
  scale_x_continuous(breaks = seq(0, 1, by = 0.2), limits = c(0,1)) +
  coord_cartesian(ylim = c(0,1)) +
  labs(x = "Predicted probability: outpatient visit or worse",
       y = "Observed probability") +
  theme_bw() +
  theme(legend.title = element_blank())

xaxis_calibrated_rug_1 <- axis_canvas(plot_calibrated_1, axis = "x") +
  geom_histogram(
    data = df_val_derivation,
    aes(x = `y>=1`),
    bins = length(unique(round(df_val_derivation$`y>=1`, 4)))) +
  scale_y_sqrt()

combined_calibrated_1 <- plot_calibrated_1 + 
  theme(legend.position = "none")

combined_calibrated_1 <- insert_xaxis_grob(
  combined_calibrated_1,
  xaxis_calibrated_rug_1,
  position = "bottom"
)

combined_calibrated_1 <- ggdraw(combined_calibrated_1)

combined_calibrated_1
```

Here we use a modification of the val.prob.ci.2() function to capture the confidence intervals for use in producing the figure. 200 boostrap resamples are used instead of 2000. This figure shows the internal calibration for the outcome of hospital admission or worse.

```
cal_inpatient <- val.prob.ci.2.modified(
  p = df_val_derivation$`y>=2`,
  y = df_val_derivation$inpatient_fct,
  # 200 boostrap resamples are used instead of 2000 in the analysis presented in the manuscript
  CL.BT = TRUE
)
```

```
## Bootstrap samples are being generated.
## 
## 
## 
## 
##  A 95% confidence interval is given for the calibration intercept, calibration slope and c-statistic.
```

```
plot_calibrated_2 <-
  df_val_derivation %>%
  mutate(
    bin = ntile(`y>=2`, n = 10),
    Linear = "Linear calibration",
    LOESS = "LOESS calibration"
  ) %>%
  group_by(bin) %>%
  mutate(
    n = n(),
    # Get ests and CIs
    bin_pred = mean(`y>=2`),
    bin_prob = mean(as.numeric(inpatient_fct)),
    se = sqrt((bin_prob * (1 - bin_prob)) / n),
    ul = bin_prob + 1.96 * se,
    ll = bin_prob - 1.96 * se
  ) %>%
  ungroup() %>%
  ggplot(aes(x = bin_pred, y = bin_prob)) +
  geom_abline(lty = 2) +
  geom_ribbon(
    data = as.data.frame(t(cal_inpatient)),
    aes(
      x = as.numeric(rownames((t(cal_inpatient)))),
      y = NULL,
      ymin = t(cal_inpatient)[, 1],
      ymax = t(cal_inpatient)[, 2]
    ),
    alpha = .3,
    fill = "#BC3C29FF"
  ) +
  geom_smooth(
    aes(x = `y>=2`, y = as.numeric(inpatient_fct), color = LOESS), 
    se = FALSE, method = "loess"
  ) +
  geom_pointrange(aes(ymin = ll, ymax = ul), size = 0.3, color = "black") +
  scale_color_manual(values = c("#BC3C29FF")) +
  scale_y_continuous(breaks = seq(0, 1, by = 0.2)) +
  scale_x_continuous(breaks = seq(0, 1, by = 0.2), limits = c(0,1)) +
  coord_cartesian(ylim = c(0,1)) +
  labs(x = "Predicted probability: hospital admission or worse",
       y = "") +
  theme_bw() +
  theme(legend.title = element_blank())

xaxis_calibrated_rug_2 <- axis_canvas(plot_calibrated_2, axis = "x") +
  geom_histogram(
    data = df_val_derivation,
    aes(x = `y>=2`),
    bins = length(unique(round(df_val_derivation$`y>=2`, 4)))) +
  scale_y_sqrt()

combined_calibrated_2 <- plot_calibrated_2 + 
  theme(legend.position = "none")

combined_calibrated_2 <- insert_xaxis_grob(
  combined_calibrated_2,
  xaxis_calibrated_rug_2,
  position = "bottom"
)

combined_calibrated_2 <- ggdraw(combined_calibrated_2)

combined_calibrated_2
```

Here we use a modification of the val.prob.ci.2() function to capture the confidence intervals for use in producing the figure. 200 boostrap resamples are used instead of 2000. This figure shows the internal calibration for the outcome of intensive care unit admission or death.

```
cal_icu_death <- val.prob.ci.2.modified(
  p = df_val_derivation$`y>=3`,
  y = df_val_derivation$icu_death_fct,
  # 200 boostrap resamples are used instead of 2000
  CL.BT = TRUE
)
```

```
## Bootstrap samples are being generated.
## 
## 
## 
## 
##  A 95% confidence interval is given for the calibration intercept, calibration slope and c-statistic.
```

```
plot_calibrated_3 <-
  df_val_derivation %>%
  mutate(
    bin = ntile(`y>=3`, n = 10),
    Linear = "Linear calibration",
    LOESS = "LOESS calibration"
  ) %>%
  group_by(bin) %>%
  mutate(
    n = n(),
    # Get ests and CIs
    bin_pred = mean(`y>=3`),
    bin_prob = mean(as.numeric(icu_death_fct)),
    se = sqrt((bin_prob * (1 - bin_prob)) / n),
    ul = bin_prob + 1.96 * se,
    ll = bin_prob - 1.96 * se
  ) %>%
  ungroup() %>%
  ggplot(aes(x = bin_pred, y = bin_prob)) +
  geom_abline(lty = 2) +
  geom_ribbon(
    data = as.data.frame(t(cal_icu_death)),
    aes(
      x = as.numeric(rownames((t(cal_icu_death)))),
      y = NULL,
      ymin = t(cal_icu_death)[, 1],
      ymax = t(cal_icu_death)[, 2]
    ),
    alpha = .3,
    fill = "#BC3C29FF"
  ) +
  geom_smooth(
    aes(x = `y>=3`, y = as.numeric(icu_death_fct), color = LOESS), 
    se = FALSE, method = "loess"
  ) +
  geom_pointrange(aes(ymin = ll, ymax = ul), size = 0.3, color = "black") +
  scale_color_manual(values = c("#BC3C29FF")) +
  scale_y_continuous(breaks = seq(0, 1, by = 0.2)) +
  scale_x_continuous(breaks = seq(0, 1, by = 0.2), limits = c(0,1)) +
  coord_cartesian(ylim = c(0,1)) +
  labs(x = "Predicted probability: ICU admission or death",
       y = "") +
  theme_bw() +
  theme(legend.title = element_blank())

xaxis_calibrated_rug_3 <- axis_canvas(plot_calibrated_3, axis = "x") +
  geom_histogram(
    data = df_val_derivation,
    aes(x = `y>=3`),
    bins = length(unique(round(df_val_derivation$`y>=3`, 4)))) +
  scale_y_sqrt()

combined_calibrated_3 <- plot_calibrated_3 + 
  theme(legend.position = "none")

combined_calibrated_3 <- insert_xaxis_grob(
  combined_calibrated_3,
  xaxis_calibrated_rug_3,
  position = "bottom"
)

combined_calibrated_3 <- ggdraw(combined_calibrated_3)

combined_calibrated_3
```

Finally, the combined figure is constructed and presented.

```
fig_combined_calibrated <- 
  plot_grid(
    combined_calibrated_1,
    combined_calibrated_2,
    combined_calibrated_3,
    labels = "AUTO",
    align = "hv",
    ncol = 3
  )

fig_combined_calibrated
```

### 8 Model statistics: Internal validation

Here we obtain the point estimates of the discrimination and calibration indices for the outcome of outpatient visit or worse

```
point_1_cal <- val.prob.ci.2(
  p = df_val_derivation$`y>=1`,
  y = df_val_derivation$outpatient_fct
)
```

```
## 
## 
##  A 95% confidence interval is given for the calibration intercept, calibration slope and c-statistic.
```

```
point_1_cal
```

```
##           Dxy       C (ROC)            R2             D      D:Chi-sq 
##  6.063550e-01  8.031775e-01  2.819761e-01  1.802058e-01  2.938344e+02 
##           D:p             U      U:Chi-sq           U:p             Q 
##  0.000000e+00 -9.866981e-04  3.966155e-01  8.201174e-01  1.811925e-01 
##         Brier     Intercept         Slope          Emax  Brier scaled 
##  1.064930e-01 -1.198004e-02  9.619047e-01  2.035189e-02  2.125361e-01 
##          Eavg           ECI 
##  1.002583e-02  3.079009e-02
```

Here we use modified code that is based on a function written by Darren L Dahly (https://darrendahly.github.io/post/homr/) to produce 100 bootstrap resamples of the indices to produce confidence intervals. Note: for each bootstrap resample a calibration plot is printed. The variation in the flexible calibration curve is a good visual representation of the bootstrap variation.

```
par(mfrow=c(2,3))
discr_indexes_1_cal <- boot_val2(
  data = df_val_derivation,
  predicted = "y>=1",
  outcome = "outpatient_fct"
)
```

```
par(mfrow=c(1,1))
```

Next we use another function courtesy of Darren L Dahly to produce the confidence intervals.

```
C_ci_1_cal <- calc_ci("C", discr_indexes_1_cal, 3)
C_ci_1_cal
```

```
## [1] "(0.768 to 0.831)"
```

```
R2_ci_1_cal <- calc_ci("R2", discr_indexes_1_cal, 3)
R2_ci_1_cal
```

```
## [1] "(0.218 to 0.335)"
```

```
B_ci_1_cal <- calc_ci("Brier", discr_indexes_1_cal, 3)
B_ci_1_cal
```

```
## [1] "(0.095 to 0.115)"
```

```
Bscaled_ci_1_cal <- calc_ci("Brier scaled", discr_indexes_1_cal, 3)
Bscaled_ci_1_cal
```

```
## [1] "(0.154 to 0.26)"
```

```
Intercept_ci_1_cal <- calc_ci("Intercept", discr_indexes_1_cal, 3)
Intercept_ci_1_cal
```

```
## [1] "(-0.165 to 0.113)"
```

```
Slope_ci_1_cal <- calc_ci("Slope", discr_indexes_1_cal, 3)
Slope_ci_1_cal
```

```
## [1] "(0.842 to 1.088)"
```

```
Emax_ci_1_cal <-  calc_ci("Emax", discr_indexes_1_cal, 3)
Emax_ci_1_cal
```

```
## [1] "(0.005 to 0.092)"
```

```
Eavg_ci_1_cal <-  calc_ci("Eavg", discr_indexes_1_cal, 3)
Eavg_ci_1_cal
```

```
## [1] "(0.011 to 0.026)"
```

The same but for hospital admission or worse.

```
point_2_cal <- val.prob.ci.2(
  p = df_val_derivation$`y>=2`,
  y = df_val_derivation$inpatient_fct
)
```

```
## 
## 
##  A 95% confidence interval is given for the calibration intercept, calibration slope and c-statistic.
```

Note: for each bootstrap resample a calibration plot is printed. The variation in the flexible calibration curve is a good visual representation of the bootstrap variation.

```
par(mfrow=c(2,3))
discr_indexes_2_cal <- boot_val2(
  data = df_val_derivation,
  predicted = "y>=2",
  outcome = "inpatient_fct"
)
```

```
par(mfrow=c(1,1))

C_ci_2_cal <- calc_ci("C", discr_indexes_2_cal, 3)
C_ci_2_cal
```

```
## [1] "(0.822 to 0.886)"
```

```
R2_ci_2_cal <- calc_ci("R2", discr_indexes_2_cal, 3)
R2_ci_2_cal
```

```
## [1] "(0.24 to 0.371)"
```

```
B_ci_2_cal <- calc_ci("Brier", discr_indexes_2_cal, 3)
B_ci_2_cal
```

```
## [1] "(0.038 to 0.054)"
```

```
Bscaled_ci_2_cal <- calc_ci("Brier scaled", discr_indexes_2_cal, 3)
Bscaled_ci_2_cal
```

```
## [1] "(0.147 to 0.243)"
```

```
Intercept_ci_2_cal <- calc_ci("Intercept", discr_indexes_2_cal, 3)
Intercept_ci_2_cal
```

```
## [1] "(-0.243 to 0.201)"
```

```
Slope_ci_2_cal <- calc_ci("Slope", discr_indexes_2_cal, 3)
Slope_ci_2_cal
```

```
## [1] "(0.969 to 1.308)"
```

```
Emax_ci_2_cal <-  calc_ci("Emax", discr_indexes_2_cal, 3)
Emax_ci_2_cal
```

```
## [1] "(0.017 to 0.149)"
```

```
Eavg_ci_2_cal <-  calc_ci("Eavg", discr_indexes_2_cal, 3)
Eavg_ci_2_cal
```

```
## [1] "(0.008 to 0.017)"
```

The same but for intensive care admission or death

```
point_3_cal <- val.prob.ci.2(
  p = df_val_derivation$`y>=3`,
  y = df_val_derivation$icu_death_fct
)
```

```
## 
## 
##  A 95% confidence interval is given for the calibration intercept, calibration slope and c-statistic.
```

Note: for each bootstrap resample a calibration plot is printed. The variation in the flexible calibration curve is a good visual representation of the bootstrap variation.

```
par(mfrow=c(2,3))
discr_indexes_3_cal <- boot_val2(
  data = df_val_derivation,
  predicted = "y>=3",
  outcome = "icu_death_fct"
)
```

```
par(mfrow=c(1,1))

C_ci_3_cal <- calc_ci("C", discr_indexes_3_cal, 3)
C_ci_3_cal
```

```
## [1] "(0.816 to 0.922)"
```

```
R2_ci_3_cal <- calc_ci("R2", discr_indexes_3_cal, 3)
R2_ci_3_cal
```

```
## [1] "(0.132 to 0.331)"
```

```
B_ci_3_cal <- calc_ci("Brier", discr_indexes_3_cal, 3)
B_ci_3_cal
```

```
## [1] "(0.011 to 0.023)"
```

```
Bscaled_ci_3_cal <- calc_ci("Brier scaled", discr_indexes_3_cal, 3)
Bscaled_ci_3_cal
```

```
## [1] "(-0.03 to 0.114)"
```

```
Intercept_ci_3_cal <- calc_ci("Intercept", discr_indexes_3_cal, 3)
Intercept_ci_3_cal
```

```
## [1] "(-0.433 to 0.38)"
```

```
Slope_ci_3_cal <- calc_ci("Slope", discr_indexes_3_cal, 3)
Slope_ci_3_cal
```

```
## [1] "(0.761 to 1.295)"
```

```
Emax_ci_3_cal <-  calc_ci("Emax", discr_indexes_3_cal, 3)
Emax_ci_3_cal
```

```
## [1] "(0.014 to 0.272)"
```

```
Eavg_ci_3_cal <-  calc_ci("Eavg", discr_indexes_3_cal, 3)
Eavg_ci_3_cal
```

```
## [1] "(0.004 to 0.013)"
```

Finally, the table is produced.

```
table_internal_validation_indices <- tbl_df(
  cbind.data.frame(
    Indexes = c(
      "C-statistic",
      "R^2",
      "Brier's score",
      "Brier's score (rescaled)",
      "Calibration intercept",
      "Calibration slope",
      "Emax",
      "Eavg"
    ),
    "Outpatient" = c(
      paste(round(point_1_cal[2], 3), C_ci_1_cal),
      paste(round(point_1_cal[3], 3), R2_ci_1_cal),
      paste(round(point_1_cal[11], 3), B_ci_1_cal),
      paste(round(point_1_cal[15], 3), Bscaled_ci_1_cal),
      paste(round(point_1_cal[12], 3), Intercept_ci_1_cal),
      paste(round(point_1_cal[13], 3), Slope_ci_1_cal),
      paste(round(point_1_cal[14], 3), Emax_ci_1_cal),
      paste(round(point_1_cal[16], 3), Eavg_ci_1_cal)
    ),
    "Hospital admission" = c(
      paste(round(point_2_cal[2], 3), C_ci_2_cal),
      paste(round(point_2_cal[3], 3), R2_ci_2_cal),
      paste(round(point_2_cal[11], 3), B_ci_2_cal),
      paste(round(point_2_cal[15], 3), Bscaled_ci_2_cal),
      paste(round(point_2_cal[12], 3), Intercept_ci_2_cal),
      paste(round(point_2_cal[13], 3), Slope_ci_2_cal),
      paste(round(point_2_cal[14], 3), Emax_ci_2_cal),
      paste(round(point_2_cal[16], 3), Eavg_ci_2_cal)
    ),
    "ICU admission or death" = c(
      paste(round(point_3_cal[2], 3), C_ci_3_cal),
      paste(round(point_3_cal[3], 3), R2_ci_3_cal),
      paste(round(point_3_cal[11], 3), B_ci_3_cal),
      paste(round(point_3_cal[15], 3), Bscaled_ci_3_cal),
      paste(round(point_3_cal[12], 3), Intercept_ci_3_cal),
      paste(round(point_3_cal[13], 3), Slope_ci_3_cal),
      paste(round(point_3_cal[14], 3), Emax_ci_3_cal),
      paste(round(point_3_cal[16], 3), Eavg_ci_3_cal)
    )
  )
)

knitr::kable(table_internal_validation_indices)
```

| Indexes | Outpatient | Hospital admission | ICU admission or death |
| --- | --- | --- | --- |
| C-statistic | 0.803 (0.768 to 0.831) | 0.852 (0.822 to 0.886) | 0.874 (0.816 to 0.922) |
| R^2 | 0.282 (0.218 to 0.335) | 0.306 (0.24 to 0.371) | 0.218 (0.132 to 0.331) |
| Brier’s score | 0.106 (0.095 to 0.115) | 0.046 (0.038 to 0.054) | 0.017 (0.011 to 0.023) |
| Brier’s score (rescaled) | 0.213 (0.154 to 0.26) | 0.196 (0.147 to 0.243) | 0.047 (-0.03 to 0.114) |
| Calibration intercept | -0.012 (-0.165 to 0.113) | -0.014 (-0.243 to 0.201) | 0.006 (-0.433 to 0.38) |
| Calibration slope | 0.962 (0.842 to 1.088) | 1.11 (0.969 to 1.308) | 1.004 (0.761 to 1.295) |
| Emax | 0.02 (0.005 to 0.092) | 0.06 (0.017 to 0.149) | 0.005 (0.014 to 0.272) |
| Eavg | 0.01 (0.011 to 0.026) | 0.006 (0.008 to 0.017) | 0.004 (0.004 to 0.013) |

### 9 External validation calibration plot

First we add the predicted probabilities for each of the included outcomes to the singly imputed dataframe and then produce variables that represent the ordinal outcome or worse.

```
df_val_validation <- df_val_validation %>%
  cbind.data.frame(
    predict(
      # here we predict using the derivation model intercept and coefficients
      mod_multi_derivation,
      df_val_validation,
      type = "fitted")
  ) %>%
  mutate(outcome = as.numeric(as.character(outcome_fct))) %>%
  mutate(
    outpatient_fct = case_when(
      outcome >= 1 ~ 1,
      outcome < 1 ~ 0
    ),
    inpatient_fct = case_when(
      outcome >= 2 ~ 1,
      outcome < 2 ~ 0
    ),
    icu_death_fct = case_when(
      outcome >= 3 ~ 1,
      outcome < 3 ~ 0
    ))
```

Here we use a modification of the val.prob.ci.2() function to capture the confidence intervals for use in producing the figure. See also the modified function definitions in a separate .R file. Note: in the analysis presented in the manuscript, the confidence intervals are constructed using 2000 bootstrap resamples. Here we only do 200 resamples to save computational time. This figure shows the external calibration for the outcome of outpatient visit or worse.

```
val_outpatient <- val.prob.ci.2.modified(
  p = df_val_validation$`y>=1`,
  y = df_val_validation$outpatient_fct,
  logit = "logit",
  CL.BT = TRUE
)
```

```
## Bootstrap samples are being generated.
## 
## 
## 
## 
##  A 95% confidence interval is given for the calibration intercept, calibration slope and c-statistic.
```

```
plot_validated_1 <-
  df_val_validation %>%
  mutate(
    bin = ntile(`y>=1`, n = 10),
    Linear = "Linear calibration",
    LOESS = "LOESS calibration"
  ) %>%
  group_by(bin) %>%
  mutate(
    n = n(),
    # Get ests and CIs
    bin_pred = mean(`y>=1`),
    bin_prob = mean(as.numeric(outpatient_fct)),
    se = sqrt((bin_prob * (1 - bin_prob)) / n),
    ul = bin_prob + 1.96 * se,
    ll = bin_prob - 1.96 * se
  ) %>%
  ungroup() %>%
  ggplot(aes(x = bin_pred, y = bin_prob)) +
  geom_abline(lty = 2) +
  geom_ribbon(
    data = as.data.frame(t(val_outpatient)),
    aes(
      x = as.numeric(rownames((t(val_outpatient)))),
      y = NULL,
      ymin = t(val_outpatient)[, 1],
      ymax = t(val_outpatient)[, 2]
    ),
    alpha = .3,
    fill = "#BC3C29FF"
  ) +
  geom_smooth(
    aes(x = `y>=1`, y = as.numeric(outpatient_fct), color = LOESS),
    se = FALSE, method = "loess"
  ) +
  geom_pointrange(aes(ymin = ll, ymax = ul), size = 0.3, color = "black") +
  scale_color_manual(values = c("#BC3C29FF")) +
  scale_y_continuous(breaks = seq(0, 1, by = 0.2)) +
  scale_x_continuous(breaks = seq(0, 1, by = 0.2), limits = c(0,1)) +
  coord_cartesian(ylim = c(0,1)) +
  labs(x = "Predicted probability: outpatient visit or worse",
       y = "Observed probability") +
  theme_bw() +
  theme(legend.title = element_blank())

xaxis_validated_rug_1 <- axis_canvas(plot_validated_1, axis = "x") +
  geom_histogram(
    data = df_val_validation,
    aes(x = `y>=1`),
    bins = length(unique(round(df_val_validation$`y>=1`, 4)))) +
  scale_y_sqrt()

combined_validated_1 <- plot_validated_1 +
  theme(legend.position = "none")

combined_validated_1 <- insert_xaxis_grob(
  combined_validated_1,
  xaxis_validated_rug_1,
  position = "bottom"
)

combined_validated_1 <- ggdraw(combined_validated_1)

combined_validated_1
```

Here we use a modification of the val.prob.ci.2() function to capture the confidence intervals for use in producing the figure. 200 boostrap resamples are used instead of 2000. This figure shows the external calibration for the outcome of hospital admission or worse.

```
val_inpatient <- val.prob.ci.2.modified(
  p = df_val_validation$`y>=2`,
  y = df_val_validation$inpatient_fct,
  CL.BT = TRUE
)
```

```
## Bootstrap samples are being generated.
## 
## 
## 
## 
##  A 95% confidence interval is given for the calibration intercept, calibration slope and c-statistic.
```

```
plot_validated_2 <-
  df_val_validation %>%
  mutate(
    bin = ntile(`y>=2`, n = 10),
    Linear = "Linear calibration",
    LOESS = "LOESS calibration"
  ) %>%
  group_by(bin) %>%
  mutate(
    n = n(),
    # Get ests and CIs
    bin_pred = mean(`y>=2`),
    bin_prob = mean(as.numeric(inpatient_fct)),
    se = sqrt((bin_prob * (1 - bin_prob)) / n),
    ul = bin_prob + 1.96 * se,
    ll = bin_prob - 1.96 * se
  ) %>%
  ungroup() %>%
  ggplot(aes(x = bin_pred, y = bin_prob)) +
  geom_abline(lty = 2) +
  geom_ribbon(
    data = as.data.frame(t(val_inpatient)),
    aes(
      x = as.numeric(rownames((t(val_inpatient)))),
      y = NULL,
      ymin = t(val_inpatient)[, 1],
      ymax = t(val_inpatient)[, 2]
    ),
    alpha = .3,
    fill = "#BC3C29FF"
  ) +
  geom_smooth(
    aes(x = `y>=2`, y = as.numeric(inpatient_fct), color = LOESS),
    se = FALSE, method = "loess"
  ) +
  geom_pointrange(aes(ymin = ll, ymax = ul), size = 0.3, color = "black") +
  scale_color_manual(values = c("#BC3C29FF")) +
  scale_y_continuous(breaks = seq(0, 1, by = 0.2)) +
  scale_x_continuous(breaks = seq(0, 1, by = 0.2), limits = c(0,1)) +
  coord_cartesian(ylim = c(0,1)) +
  labs(x = "Predicted probability: hospital admission or worse",
       y = "") +
  theme_bw() +
  theme(legend.title = element_blank())

xaxis_validated_rug_2 <- axis_canvas(plot_validated_2, axis = "x") +
  geom_histogram(
    data = df_val_validation,
    aes(x = `y>=2`),
    bins = length(unique(round(df_val_validation$`y>=2`, 4)))) +
  scale_y_sqrt()

combined_validated_2 <- plot_validated_2 +
  theme(legend.position = "none")

combined_validated_2 <- insert_xaxis_grob(
  combined_validated_2,
  xaxis_validated_rug_2,
  position = "bottom"
)

combined_validated_2 <- ggdraw(combined_validated_2)

combined_validated_2
```

Here we use a modification of the val.prob.ci.2() function to capture the confidence intervals for use in producing the figure. 200 boostrap resamples are used instead of 2000. This figure shows the external calibration for the outcome of intensive care unit admission or death.

```
val_icu_death <- val.prob.ci.2.modified(
  p = df_val_validation$`y>=3`,
  y = df_val_validation$icu_death_fct,
  CL.BT = TRUE
)
```

```
## Bootstrap samples are being generated.
## 
## 
## 
## 
##  A 95% confidence interval is given for the calibration intercept, calibration slope and c-statistic.
```

```
plot_validated_3 <-
  df_val_validation %>%
  mutate(
    bin = ntile(`y>=3`, n = 10),
    Linear = "Linear calibration",
    LOESS = "LOESS calibration"
  ) %>%
  group_by(bin) %>%
  mutate(
    n = n(),
    # Get ests and CIs
    bin_pred = mean(`y>=3`),
    bin_prob = mean(as.numeric(icu_death_fct)),
    se = sqrt((bin_prob * (1 - bin_prob)) / n),
    ul = bin_prob + 1.96 * se,
    ll = bin_prob - 1.96 * se
  ) %>%
  ungroup() %>%
  ggplot(aes(x = bin_pred, y = bin_prob)) +
  geom_abline(lty = 2) +
  geom_ribbon(
    data = as.data.frame(t(val_icu_death)),
    aes(
      x = as.numeric(rownames((t(val_icu_death)))),
      y = NULL,
      ymin = t(val_icu_death)[, 1],
      ymax = t(val_icu_death)[, 2]
    ),
    alpha = .3,
    fill = "#BC3C29FF"
  ) +
  geom_smooth(
    aes(x = `y>=3`, y = as.numeric(icu_death_fct), color = LOESS),
    se = FALSE, method = "loess"
  ) +
  geom_pointrange(aes(ymin = ll, ymax = ul), size = 0.3, color = "black") +
  scale_color_manual(values = c("#BC3C29FF")) +
  scale_y_continuous(breaks = seq(0, 1, by = 0.2)) +
  scale_x_continuous(breaks = seq(0, 1, by = 0.2), limits = c(0,1)) +
  coord_cartesian(ylim = c(0,1)) +
  labs(x = "Predicted probability: ICU admission or death",
       y = "") +
  theme_bw() +
  theme(legend.title = element_blank())

xaxis_validated_rug_3 <- axis_canvas(plot_validated_3, axis = "x") +
  geom_histogram(
    data = df_val_validation,
    aes(x = `y>=3`),
    bins = length(unique(round(df_val_validation$`y>=3`, 4)))) +
  scale_y_sqrt()

combined_validated_3 <- plot_validated_3 +
  theme(legend.position = "none")

combined_validated_3 <- insert_xaxis_grob(
  combined_validated_3,
  xaxis_validated_rug_3,
  position = "bottom"
)

combined_validated_3 <- ggdraw(combined_validated_3)

combined_validated_3
```

Finally, the combined figure is constructed and presented.

```
fig_combined_validated <-
  plot_grid(
    combined_validated_1,
    combined_validated_2,
    combined_validated_3,
    labels = "AUTO",
    align = "hv",
    ncol = 3
  )

fig_combined_validated
```

### 10 Model statistics: External validation

Here we obtain the point estimates of the discrimination and calibration indices for the outcome of outpatient visit or worse

```
point_1_val <- val.prob.ci.2(
  p = df_val_validation$`y>=1`,
  y = df_val_validation$outpatient_fct
)
```

```
## 
## 
##  A 95% confidence interval is given for the calibration intercept, calibration slope and c-statistic.
```

Here we use modified code that is based on a function written by Darren L Dahly (https://darrendahly.github.io/post/homr/) to produce 60 (100 are used in the analysis presented in the manuscript but is 60 here for the sake of brevity of the output) bootstrap resamples of the indices to produce confidence intervals. Note: for each bootstrap resample a calibration plot is printed. The variation in the flexible calibration curve is a good visual representation of the bootstrap variation.

```
par(mfrow=c(2,3))
discr_indexes_1_val <- boot_val2(
  data = df_val_validation,
  predicted = "y>=1",
  outcome = "outpatient_fct"
)
```

```
par(mfrow=c(1,1))
```

Next we use another function courtesy of Darren L Dahly to produce the confidence intervals.

```
C_ci_1_val <- calc_ci("C", discr_indexes_1_val, 3)
C_ci_1_val
```

```
## [1] "(0.719 to 0.77)"
```

```
R2_ci_1_val <- calc_ci("R2", discr_indexes_1_val, 3)
R2_ci_1_val
```

```
## [1] "(0.161 to 0.229)"
```

```
B_ci_1_val <- calc_ci("Brier", discr_indexes_1_val, 3)
B_ci_1_val
```

```
## [1] "(0.079 to 0.092)"
```

```
Bscaled_ci_1_val <- calc_ci("Brier scaled", discr_indexes_1_val, 3)
Bscaled_ci_1_val
```

```
## [1] "(0.114 to 0.183)"
```

```
Intercept_ci_1_val <- calc_ci("Intercept", discr_indexes_1_val, 3)
Intercept_ci_1_val
```

```
## [1] "(-0.036 to 0.154)"
```

```
Slope_ci_1_val <- calc_ci("Slope", discr_indexes_1_val, 3)
Slope_ci_1_val
```

```
## [1] "(0.823 to 1.028)"
```

```
Emax_ci_1_val <-  calc_ci("Emax", discr_indexes_1_val, 3)
Emax_ci_1_val
```

```
## [1] "(0.005 to 0.105)"
```

```
Eavg_ci_1_val <-  calc_ci("Eavg", discr_indexes_1_val, 3)
Eavg_ci_1_val
```

```
## [1] "(0.006 to 0.023)"
```

The same but for hospital admission or worse.

```
point_2_val <- val.prob.ci.2(
  p = df_val_validation$`y>=2`,
  y = df_val_validation$inpatient_fct
)
```

```
## 
## 
##  A 95% confidence interval is given for the calibration intercept, calibration slope and c-statistic.
```

Note: for each bootstrap resample a calibration plot is printed. The variation in the flexible calibration curve is a good visual representation of the bootstrap variation.

```
par(mfrow=c(2,3))
discr_indexes_2_val <- boot_val2(
  data = df_val_validation,
  predicted = "y>=2",
  outcome = "inpatient_fct"
)
```

```
par(mfrow=c(1,1))

C_ci_2_val <- calc_ci("C", discr_indexes_2_val, 3)
C_ci_2_val
```

```
## [1] "(0.772 to 0.845)"
```

```
R2_ci_2_val <- calc_ci("R2", discr_indexes_2_val, 3)
R2_ci_2_val
```

```
## [1] "(0.187 to 0.293)"
```

```
B_ci_2_val <- calc_ci("Brier", discr_indexes_2_val, 3)
B_ci_2_val
```

```
## [1] "(0.03 to 0.041)"
```

```
Bscaled_ci_2_val <- calc_ci("Brier scaled", discr_indexes_2_val, 3)
Bscaled_ci_2_val
```

```
## [1] "(0.099 to 0.197)"
```

```
Intercept_ci_2_val <- calc_ci("Intercept", discr_indexes_2_val, 3)
Intercept_ci_2_val
```

```
## [1] "(-0.02 to 0.332)"
```

```
Slope_ci_2_val <- calc_ci("Slope", discr_indexes_2_val, 3)
Slope_ci_2_val
```

```
## [1] "(0.888 to 1.169)"
```

```
Emax_ci_2_val <-  calc_ci("Emax", discr_indexes_2_val, 3)
Emax_ci_2_val
```

```
## [1] "(0.012 to 0.179)"
```

```
Eavg_ci_2_val <-  calc_ci("Eavg", discr_indexes_2_val, 3)
Eavg_ci_2_val
```

```
## [1] "(0.005 to 0.026)"
```

The same but for intensive care admission or death

```
point_3_val <- val.prob.ci.2(
  p = df_val_validation$`y>=3`,
  y = df_val_validation$icu_death_fct
)
```

```
## 
## 
##  A 95% confidence interval is given for the calibration intercept, calibration slope and c-statistic.
```

Note: for each bootstrap resample a calibration plot is printed. The variation in the flexible calibration curve is a good visual representation of the bootstrap variation.

```
par(mfrow=c(2,3))
discr_indexes_3_val <- boot_val2(
  data = df_val_validation,
  predicted = "y>=3",
  outcome = "icu_death_fct"
)
```

```
par(mfrow=c(1,1))

C_ci_3_val <- calc_ci("C", discr_indexes_3_val, 3)
C_ci_3_val
```

```
## [1] "(0.787 to 0.927)"
```

```
R2_ci_3_val <- calc_ci("R2", discr_indexes_3_val, 3)
R2_ci_3_val
```

```
## [1] "(0.101 to 0.265)"
```

```
B_ci_3_val <- calc_ci("Brier", discr_indexes_3_val, 3)
B_ci_3_val
```

```
## [1] "(0.004 to 0.01)"
```

```
Bscaled_ci_3_val <- calc_ci("Brier scaled", discr_indexes_3_val, 3)
Bscaled_ci_3_val
```

```
## [1] "(-0.235 to 0.049)"
```

```
Intercept_ci_3_val <- calc_ci("Intercept", discr_indexes_3_val, 3)
Intercept_ci_3_val
```

```
## [1] "(-1.099 to -0.073)"
```

```
Slope_ci_3_val <- calc_ci("Slope", discr_indexes_3_val, 3)
Slope_ci_3_val
```

```
## [1] "(0.7 to 1.096)"
```

```
Emax_ci_3_val <-  calc_ci("Emax", discr_indexes_3_val, 3)
Emax_ci_3_val
```

```
## [1] "(0.045 to 0.478)"
```

```
Eavg_ci_3_val <-  calc_ci("Eavg", discr_indexes_3_val, 3)
Eavg_ci_3_val
```

```
## [1] "(0.004 to 0.02)"
```

Finally, the table is produced.

```
table_external_validation_indices <- tbl_df(
  cbind.data.frame(
    Indexes = c(
      "C-statistic",
      "R^2",
      "Brier's score",
      "Brier's score (rescaled)",
      "Calibration intercept",
      "Calibration slope",
      "Emax",
      "Eavg"
    ),
    "Outpatient" = c(
      paste(round(point_1_val[2], 3), C_ci_1_val),
      paste(round(point_1_val[3], 3), R2_ci_1_val),
      paste(round(point_1_val[11], 3), B_ci_1_val),
      paste(round(point_1_val[15], 3), Bscaled_ci_1_val),
      paste(round(point_1_val[12], 3), Intercept_ci_1_val),
      paste(round(point_1_val[13], 3), Slope_ci_1_val),
      paste(round(point_1_val[14], 3), Emax_ci_1_val),
      paste(round(point_1_val[16], 3), Eavg_ci_1_val)
    ),
    "Hospital admission" = c(
      paste(round(point_2_val[2], 3), C_ci_2_val),
      paste(round(point_2_val[3], 3), R2_ci_2_val),
      paste(round(point_2_val[11], 3), B_ci_2_val),
      paste(round(point_2_val[15], 3), Bscaled_ci_2_val),
      paste(round(point_2_val[12], 3), Intercept_ci_2_val),
      paste(round(point_2_val[13], 3), Slope_ci_2_val),
      paste(round(point_2_val[14], 3), Emax_ci_2_val),
      paste(round(point_2_val[16], 3), Eavg_ci_2_val)
    ),
    "ICU admission or death" = c(
      paste(round(point_3_val[2], 3), C_ci_3_val),
      paste(round(point_3_val[3], 3), R2_ci_3_val),
      paste(round(point_3_val[11], 3), B_ci_3_val),
      paste(round(point_3_val[15], 3), Bscaled_ci_3_val),
      paste(round(point_3_val[12], 3), Intercept_ci_3_val),
      paste(round(point_3_val[13], 3), Slope_ci_3_val),
      paste(round(point_3_val[14], 3), Emax_ci_3_val),
      paste(round(point_3_val[16], 3), Eavg_ci_3_val)
    )
  )
)

knitr::kable(table_external_validation_indices)
```

| Indexes | Outpatient | Hospital admission | ICU admission or death |
| --- | --- | --- | --- |
| C-statistic | 0.743 (0.719 to 0.77) | 0.809 (0.772 to 0.845) | 0.855 (0.787 to 0.927) |
| R^2 | 0.194 (0.161 to 0.229) | 0.246 (0.187 to 0.293) | 0.162 (0.101 to 0.265) |
| Brier’s score | 0.085 (0.079 to 0.092) | 0.036 (0.03 to 0.041) | 0.007 (0.004 to 0.01) |
| Brier’s score (rescaled) | 0.147 (0.114 to 0.183) | 0.157 (0.099 to 0.197) | -0.07 (-0.235 to 0.049) |
| Calibration intercept | 0.044 (-0.036 to 0.154) | 0.171 (-0.02 to 0.332) | -0.563 (-1.099 to -0.073) |
| Calibration slope | 0.918 (0.823 to 1.028) | 1.032 (0.888 to 1.169) | 0.858 (0.7 to 1.096) |
| Emax | 0.04 (0.005 to 0.105) | 0.064 (0.012 to 0.179) | 0.281 (0.045 to 0.478) |
| Eavg | 0.01 (0.006 to 0.023) | 0.011 (0.005 to 0.026) | 0.005 (0.004 to 0.02) |

### 11 Decision curve analysis

Here we show the code the produces the decision curve analysis shown in the supplement.

```
df_val_validation <- df_val_validation %>%
  mutate(
    validation_outcome = as.numeric(as.character(outcome_fct)),
    validation_outcome = if_else(validation_outcome >= 1, 1L, 0L),
    validation_outcome2 = as.numeric(as.character(outcome_fct)),
    validation_outcome2 = if_else(validation_outcome2 >= 2, 1L, 0L)
  )
```

Formula obtained from Function(mod\_multi\_derivation) and the intercept for the outcome in question is added.

```
mod_multi_derivation$coefficients
```

```
##                 y>=1                 y>=2                 y>=3 
##         -4.013091209         -5.354681369         -6.835070202 
##                  age                 age'                age'' 
##          0.063268780         -0.080158608         -0.013653520 
##               age'''                  sex      bmi_prospective 
##          0.648892481         -0.154180568         -0.011915320 
##      htn_prospective    heart_prospective     pulm_prospective 
##         -0.030030515          0.003455387          0.185225920 
## diabetes_prospective   cancer_prospective      smoking_current 
##          0.695007365         -0.451359558         -0.156952206 
##     flulike_symptoms    upper_respiratory    lower_respiratory 
##         -0.391118579         -0.289672028          0.307351189 
##     gastrointestinal   clinical_score_fct 
##          0.521071192          1.857745119
```

```
Function(mod_multi_validation)
```

```
## function (age = 60, sex = 0, bmi_prospective = 25, htn_prospective = 0, 
##     heart_prospective = 0, pulm_prospective = 0, diabetes_prospective = 0, 
##     cancer_prospective = 0, smoking_current = 0, flulike_symptoms = 0, 
##     upper_respiratory = 0, lower_respiratory = 0, gastrointestinal = 0, 
##     clinical_score_fct = 0) 
## {
##     +0.087192992 * age - 0.00016572945 * pmax(age - 20, 0)^3 + 
##         0.00033722722 * pmax(age - 30, 0)^3 - 0.00021496456 * 
##         pmax(age - 46, 0)^3 + 4.346679e-05 * pmax(age - 71, 0)^3 - 
##         0.22854614 * sex + 0.048391215 * bmi_prospective - 0.091216687 * 
##         htn_prospective - 0.0037183195 * heart_prospective + 
##         0.94295724 * pulm_prospective + 0.36728339 * diabetes_prospective + 
##         0.47821803 * cancer_prospective + 0.133408 * smoking_current + 
##         0.8089821 * flulike_symptoms - 0.029126342 * upper_respiratory + 
##         0.40339332 * lower_respiratory + 0.25143489 * gastrointestinal + 
##         1.3425161 * clinical_score_fct
## }
## <environment: 0x7ff1950c2d30>
```

The numbers that are used in the following code are those used in the main analysis presented in the manuscript, not those that result from all the code above. Here we show the prediction results for outpatient visit or worse.

```
df_val_validation$decision_curve_mainmodel <-
  with(
    df_val_validation,
    expit(
      -4.0254 + 0.061663814 * age - 3.4200024e-05 * pmax(age - 21, 0) ^ 3 + 1.8974129e-06 *
        pmax(age - 32, 0) ^ 3 + 0.00025025406 * pmax(age - 45, 0) ^ 3 - 0.0003101768 *
        pmax(age - 55, 0) ^ 3 + 9.222535e-05 * pmax(age - 70, 0) ^ 3 - 0.16878381 *
        sex - 0.0095656848 * bmi_prospective - 0.023077179 * htn_prospective - 0.063300568 *
        heart_prospective + 0.10252031 * pulm_prospective + 0.67146533 * diabetes_prospective -
        0.39568289 * cancer_prospective - 0.15070199 * smoking_current - 0.3101669 *
        flulike_symptoms - 0.32716673 * upper_respiratory + 0.27943717 * lower_respiratory +
        0.49405381 * gastrointestinal + 1.865029 * clinical_score_fct
    )
  )

# and here we produce the prediction results for hospital admission or worse. 

df_val_validation$decision_curve_mainmodel2 <-
  with(
    df_val_validation,
    expit(
      -5.3625 + 0.061663814 * age - 3.4200024e-05 * pmax(age - 21, 0) ^ 3 + 1.8974129e-06 *
        pmax(age - 32, 0) ^ 3 + 0.00025025406 * pmax(age - 45, 0) ^ 3 - 0.0003101768 *
        pmax(age - 55, 0) ^ 3 + 9.222535e-05 * pmax(age - 70, 0) ^ 3 - 0.16878381 *
        sex - 0.0095656848 * bmi_prospective - 0.023077179 * htn_prospective - 0.063300568 *
        heart_prospective + 0.10252031 * pulm_prospective + 0.67146533 * diabetes_prospective -
        0.39568289 * cancer_prospective - 0.15070199 * smoking_current - 0.3101669 *
        flulike_symptoms - 0.32716673 * upper_respiratory + 0.27943717 * lower_respiratory +
        0.49405381 * gastrointestinal + 1.865029 * clinical_score_fct
    )
  )
```

The discrimination and calibration for the outcome of intensive care admission or death were not sufficiently good to warrant producing a decision curve. Finally, produce the decision curve analysis with the following code. In the main paper, we use 10.000 bootstraps, but here we use 1.000 to lower computational time. First we produce the full\_model for the outcome of outpatient visit or worse.

```
full_model <- decision_curve(validation_outcome ~ decision_curve_mainmodel,
                             data = df_val_validation,
                             fitted.risk = TRUE,
                             policy = 'opt-out',
                             thresholds = seq(0, 1, by = .001),
                             bootstraps = 1000)
```

Next we produce full\_model2 for the outcome of hospital admission or worse.

```
full_model2 <- decision_curve(validation_outcome2 ~ decision_curve_mainmodel2,
                              data = df_val_validation,
                              fitted.risk = TRUE,
                              policy = 'opt-in',
                              thresholds = seq(0, 1, by = .001),
                              bootstraps = 1000)
```

The following code produces the decision curve figure for outpatient visit or worse

```
plot_outpatient_dca <- full_model$derived.data %>%
  select(thresholds, sNB, sNB_lower, sNB_upper, model) %>%
  ggplot(aes(x = thresholds, y = sNB, color = model, fill = model)) +
  geom_line() +
  geom_ribbon(aes(ymin = sNB_lower, ymax = sNB_upper, color = NULL), alpha = 0.5) +
  scale_color_manual(
    name = "Follow-up strategy:",
    labels = c(
      "Intervene on all",
      "Intervene on none",
      "Intervene based on prognostic model"
    ),
    values = c("black", nejm_palette[c(1, 2)])
  ) +
  scale_fill_manual(
    name = "Follow-up strategy:",
    labels = c(
      "Intervene on all",
      "Intervene on none",
      "Intervene based on prognostic model"
    ),
    values = c("black", nejm_palette[c(1, 2)])
  ) +
  scale_x_continuous(breaks = seq(0, 1, by = 0.01), labels = scales::percent_format(accuracy = 1)) +
  coord_cartesian(xlim = c(0, 0.15), ylim = c(-0.2, 1)) +
  labs(x = "Low Risk Threshold (Opt-out)", y = "Net Benefit") +
  theme_bw() +
  theme(
    panel.grid.minor.x = element_blank(),
    legend.position = c(0.35, 0.7)
  )

plot_outpatient_dca
```

The following code produces the decision curve figure for hospital admission or worse

```
plot_inpatient_dca <- full_model2$derived.data %>%
  select(thresholds, sNB, sNB_lower, sNB_upper, model) %>%
  ggplot(aes(x = thresholds, y = sNB, color = model, fill = model)) +
  geom_line() +
  geom_ribbon(aes(ymin = sNB_lower, ymax = sNB_upper, color = NULL), alpha = 0.5) +
  scale_color_manual(
    name = "Follow-up strategy:",
    labels = c(
      "Intervene on all",
      "Intervene on none",
      "Intervene based on prognostic model"
    ),
    values = c("black", nejm_palette[c(1, 2)])
  ) +
  scale_fill_manual(
    name = "Follow-up strategy:",
    labels = c(
      "Intervene on all",
      "Intervene on none",
      "Intervene based on prognostic model"
    ),
    values = c("black", nejm_palette[c(1, 2)])
  ) +
  scale_x_continuous(breaks = seq(0, 1, by = 0.01), labels = scales::percent_format(accuracy = 1)) +
  coord_cartesian(xlim = c(0, 0.15), ylim = c(-0.2, 1)) +
  labs(x = "High Risk Threshold (Opt-in)", y = "") +
  theme_bw() +
  theme(
    panel.grid.minor.x = element_blank(),
    legend.position = "none")

plot_inpatient_dca
```

and here we show the combind figure.

```
fig_combined_dca <-
  plot_grid(
    plot_outpatient_dca,
    plot_inpatient_dca,
    labels = "AUTO",
    align = "hv",
    ncol = 2
  )

fig_combined_dca
```

### 12 Risk factor analysis

The following code produces the risk factor analysis. The variables selected into each model are based on the minimal sufficient adjustment set as described by the directed acyclic graph. This was produced with dagitty and is available at dagitty.net/mY9jiTj

```
plot(dag_outcome)
```

```
dd <- datadist(df_outcome %>% filter(excluded == "Included"))
dd$limits$age <- c(45, 60, 75, 25, 88, 18, 102)
dd$limits$bmi <- c(25, 25, 35, 15, 44, 12, 56)
dd$limits$eGFR <- c(45, 90, 150, 30, 250, 10, 278)
options(datadist = "dd")
```

#### 12.1 Risk factor: Age

Here we show the odds ratio of outpatient visit or worse for an adult 75 years of age compared to 40 years of age.

```
age_1_OR <- round(summary(fit.mult.impute(
  logistic_outpatient ~ rcs(age, 4),
  lrm,
  imputation_outcomes,
  data = df_outcome %>% filter(excluded == "Included"))
)[2, c(4,6,7)], 2)
```

```
## 
## Variance Inflation Factors Due to Imputation:
## 
## Intercept       age      age'     age'' 
##         1         1         1         1 
## 
## Rate of Missing Information:
## 
## Intercept       age      age'     age'' 
##         0         0         0         0 
## 
## d.f. for t-distribution for Tests of Single Coefficients:
## 
##    Intercept          age         age'        age'' 
##          Inf 5.921027e+29 9.362153e+29 3.530174e+29 
## 
## The following fit components were averaged over the 20 model fits:
## 
##   stats linear.predictors
```

```
age_1_OR
```

```
##     Effect Lower 0.95 Upper 0.95 
##       5.29       4.11       6.80
```

Here we show the odds ratio of hospital admission or worse for an adult 75 years of age compared to 40 years of age.

```
age_2_OR <- round(summary(fit.mult.impute(
  logistic_inpatient ~ rcs(age, 4),
  lrm,
  imputation_outcomes,
  data = df_outcome %>% filter(excluded == "Included"))
)[2, c(4,6,7)], 2)
```

```
## 
## Variance Inflation Factors Due to Imputation:
## 
## Intercept       age      age'     age'' 
##         1         1         1         1 
## 
## Rate of Missing Information:
## 
## Intercept       age      age'     age'' 
##         0         0         0         0 
## 
## d.f. for t-distribution for Tests of Single Coefficients:
## 
##    Intercept          age         age'        age'' 
## 3.621470e+30 1.817442e+33 2.590981e+37 2.794374e+33 
## 
## The following fit components were averaged over the 20 model fits:
## 
##   stats linear.predictors
```

```
age_2_OR
```

```
##     Effect Lower 0.95 Upper 0.95 
##      17.34      12.32      24.40
```

Here we show the odds ratio of intensive care unit admission or death for an adult 75 years of age compared to 40 years of age.

```
age_3_OR <- round(summary(fit.mult.impute(
  logistic_icu ~ rcs(age, 4),
  lrm,
  imputation_outcomes,
  data = df_outcome %>% filter(excluded == "Included"))
)[2, c(4,6,7)], 2)
```

```
## 
## Variance Inflation Factors Due to Imputation:
## 
## Intercept       age      age'     age'' 
##         1         1         1         1 
## 
## Rate of Missing Information:
## 
## Intercept       age      age'     age'' 
##         0         0         0         0 
## 
## d.f. for t-distribution for Tests of Single Coefficients:
## 
##    Intercept          age         age'        age'' 
## 1.333551e+31 2.515815e+32 1.208265e+34 1.470238e+35 
## 
## The following fit components were averaged over the 20 model fits:
## 
##   stats linear.predictors
```

```
age_3_OR
```

```
##     Effect Lower 0.95 Upper 0.95 
##       9.84       4.57      21.22
```

Here we show a figure that displays the OR of each outcome as a function of age

```
plot_OR_age <- Predict(
  fit.mult.impute(
    logistic_icu ~ rcs(age, 4),
    lrm,
    imputation_outcomes,
    data = df_outcome %>% filter(excluded == "Included")
  ),
  age,
  ref.zero = TRUE,
  fun = exp
) %>%
  mutate(outcome = "ICU or death") %>%
  union_all(
    Predict(
      fit.mult.impute(
        logistic_inpatient ~ rcs(age, 4),
        lrm,
        imputation_outcomes,
        data = df_outcome %>% filter(excluded == "Included")
      ),
      age,
      ref.zero = TRUE,
      fun = exp
    ) %>%
      mutate(outcome = "Hospital admission or worse")
  ) %>%
  union_all(
    Predict(
      fit.mult.impute(
        logistic_outpatient ~ rcs(age, 4),
        lrm,
        imputation_outcomes,
        data = df_outcome %>% filter(excluded == "Included")
      ),
      age,
      ref.zero = TRUE,
      fun = exp
    ) %>%
      mutate(outcome = "Outpatient visit or worse")
  ) %>%
  as.data.frame() %>%
  mutate(
    outcome = factor(
      outcome,
      levels = c(
        "Outpatient visit or worse",
        "Hospital admission or worse",
        "ICU or death"
      ))
  ) %>%
  ggplot(aes(x = age, y = yhat, color = outcome, fill = outcome)) +
  geom_line() +
  geom_ribbon(aes(ymin = lower, ymax = upper, color = NULL), alpha = .3) +
  geom_hline(yintercept = 1, lty = 2) +
  scale_color_manual(values = nejm_palette[c(1, 2, 4)]) +
  scale_fill_manual(values = nejm_palette[c(1, 2, 4)]) +
  scale_y_continuous() +
  scale_x_continuous(breaks = c(30, 40, 50, 60, 70, 80)) +
  scale_y_log10(
    breaks = c(0.001, 0.01, 0.05, 0.25, 1, 4, 16, 64),
    labels = c("0.001", "0.01", "0.05", "0.25", "1", "4", "16", "64"),
    minor_break = NULL,
    limits = c(0.00001, 64)
  ) +
  annotation_logticks(side = "l") +
  coord_cartesian(xlim = c(30, 85), ylim = c(0.001, 64)) +
  labs(x = "Age", y = "Odds ratio of outcome compared to 60 year old") +
  theme_bw() +
  theme(
    legend.title = element_blank(),
    legend.position = "bottom")
```

```
## 
## Variance Inflation Factors Due to Imputation:
## 
## Intercept       age      age'     age'' 
##         1         1         1         1 
## 
## Rate of Missing Information:
## 
## Intercept       age      age'     age'' 
##         0         0         0         0 
## 
## d.f. for t-distribution for Tests of Single Coefficients:
## 
##    Intercept          age         age'        age'' 
## 1.333551e+31 2.515815e+32 1.208265e+34 1.470238e+35 
## 
## The following fit components were averaged over the 20 model fits:
## 
##   stats linear.predictors 
## 
## 
## Variance Inflation Factors Due to Imputation:
## 
## Intercept       age      age'     age'' 
##         1         1         1         1 
## 
## Rate of Missing Information:
## 
## Intercept       age      age'     age'' 
##         0         0         0         0 
## 
## d.f. for t-distribution for Tests of Single Coefficients:
## 
##    Intercept          age         age'        age'' 
## 3.621470e+30 1.817442e+33 2.590981e+37 2.794374e+33 
## 
## The following fit components were averaged over the 20 model fits:
## 
##   stats linear.predictors 
## 
## 
## Variance Inflation Factors Due to Imputation:
## 
## Intercept       age      age'     age'' 
##         1         1         1         1 
## 
## Rate of Missing Information:
## 
## Intercept       age      age'     age'' 
##         0         0         0         0 
## 
## d.f. for t-distribution for Tests of Single Coefficients:
## 
##    Intercept          age         age'        age'' 
##          Inf 5.921027e+29 9.362153e+29 3.530174e+29 
## 
## The following fit components were averaged over the 20 model fits:
## 
##   stats linear.predictors
```

```
plot_OR_age
```

#### 12.2 Risk factor: Sex

Here we show the OR of outpatient visit or worse for a male compared to a female

```
sex_1_OR <- round(summary(fit.mult.impute(
  logistic_outpatient ~ sex,
  lrm,
  imputation_outcomes,
  data = df_outcome %>% filter(excluded == "Included"))
)[2, c(4,6,7)], 2)
```

```
## 
## Variance Inflation Factors Due to Imputation:
## 
## Intercept       sex 
##         1         1 
## 
## Rate of Missing Information:
## 
## Intercept       sex 
##         0         0 
## 
## d.f. for t-distribution for Tests of Single Coefficients:
## 
##    Intercept          sex 
## 8.827792e+25          Inf 
## 
## The following fit components were averaged over the 20 model fits:
## 
##   stats linear.predictors
```

```
sex_1_OR
```

```
##     Effect Lower 0.95 Upper 0.95 
##       0.68       0.57       0.81
```

Here we show the OR of hospital admission or worse for a male compared to a female

```
sex_2_OR <- round(summary(fit.mult.impute(
  logistic_inpatient ~ sex,
  lrm,
  imputation_outcomes,
  data = df_outcome %>% filter(excluded == "Included"))
)[2, c(4,6,7)], 2)
```

```
## 
## Variance Inflation Factors Due to Imputation:
## 
## Intercept       sex 
##         1         1 
## 
## Rate of Missing Information:
## 
## Intercept       sex 
##         0         0 
## 
## d.f. for t-distribution for Tests of Single Coefficients:
## 
##    Intercept          sex 
## 1.817315e+26 7.137654e+31 
## 
## The following fit components were averaged over the 20 model fits:
## 
##   stats linear.predictors
```

```
sex_2_OR
```

```
##     Effect Lower 0.95 Upper 0.95 
##       1.14       0.87       1.48
```

Here we show the OR of intensive care admission or death for a male compared to a female.

```
sex_3_OR <- round(summary(fit.mult.impute(
  logistic_icu ~ sex,
  lrm,
  imputation_outcomes,
  data = df_outcome %>% filter(excluded == "Included"))
)[2, c(4,6,7)], 2)
```

```
## 
## Variance Inflation Factors Due to Imputation:
## 
## Intercept       sex 
##         1         1 
## 
## Rate of Missing Information:
## 
## Intercept       sex 
##         0         0 
## 
## d.f. for t-distribution for Tests of Single Coefficients:
## 
##    Intercept          sex 
##          Inf 1.514914e+31 
## 
## The following fit components were averaged over the 20 model fits:
## 
##   stats linear.predictors
```

```
sex_3_OR
```

```
##     Effect Lower 0.95 Upper 0.95 
##       1.68       0.94       2.99
```

#### 12.3 Risk factor: BMI

Here we show the univariate model for BMI. Importantly, for this portion of the analysis, we also use weight and height information that was obtained from medical records to enrich the prospectively collected height and weight data so that more individuals have complete measured information. This reduces the variance produced by multiply imputing the missing values. This also applies the multivariable model for BMI shown below. For the risk factor analysis, BMI is always modeled as a restricted cubic spline with four knots. Here we show the univariate odds ratio for outpatient visit or worse of an adult with a BMI of 35 compared to an adult with a BMI of 25.

```
bmi_1_OR <- round(summary(fit.mult.impute(
  logistic_outpatient ~ rcs(bmi, 4),
  lrm,
  imputation_outcomes,
  data = df_outcome %>% filter(excluded == "Included"))
)[2, c(4,6,7)], 2)
```

```
## 
## Variance Inflation Factors Due to Imputation:
## 
## Intercept       bmi      bmi'     bmi'' 
##      1.26      1.25      1.22      1.21 
## 
## Rate of Missing Information:
## 
## Intercept       bmi      bmi'     bmi'' 
##      0.20      0.20      0.18      0.17 
## 
## d.f. for t-distribution for Tests of Single Coefficients:
## 
## Intercept       bmi      bmi'     bmi'' 
##    459.65    476.15    603.96    643.37 
## 
## The following fit components were averaged over the 20 model fits:
## 
##   stats linear.predictors
```

```
bmi_1_OR
```

```
##     Effect Lower 0.95 Upper 0.95 
##       2.30       1.79       2.96
```

Here we show the univariate odds ratio for hospital admission or worse of an adult with a BMI of 35 compared to an adult with a BMI of 25.

```
bmi_2_OR <- round(summary(fit.mult.impute(
  logistic_inpatient ~ rcs(bmi, 4),
  lrm,
  imputation_outcomes,
  data = df_outcome %>% filter(excluded == "Included"))
)[2, c(4,6,7)], 2)
```

```
## 
## Variance Inflation Factors Due to Imputation:
## 
## Intercept       bmi      bmi'     bmi'' 
##      1.46      1.47      1.47      1.46 
## 
## Rate of Missing Information:
## 
## Intercept       bmi      bmi'     bmi'' 
##      0.32      0.32      0.32      0.32 
## 
## d.f. for t-distribution for Tests of Single Coefficients:
## 
## Intercept       bmi      bmi'     bmi'' 
##    191.07    188.49    188.49    189.44 
## 
## The following fit components were averaged over the 20 model fits:
## 
##   stats linear.predictors
```

```
bmi_2_OR
```

```
##     Effect Lower 0.95 Upper 0.95 
##       2.88       1.95       4.24
```

Here we show the univariate odds ratio for intensive care admission or death of an adult with a BMI of 35 compared to an adult with a BMI of 25.

```
bmi_3_OR <- round(summary(fit.mult.impute(
  logistic_icu ~ rcs(bmi, 4),
  lrm,
  imputation_outcomes,
  data = df_outcome %>% filter(excluded == "Included"))
)[2, c(4,6,7)], 2)
```

```
## 
## Variance Inflation Factors Due to Imputation:
## 
## Intercept       bmi      bmi'     bmi'' 
##      1.28      1.30      1.38      1.41 
## 
## Rate of Missing Information:
## 
## Intercept       bmi      bmi'     bmi'' 
##      0.22      0.23      0.28      0.29 
## 
## d.f. for t-distribution for Tests of Single Coefficients:
## 
## Intercept       bmi      bmi'     bmi'' 
##    390.39    362.38    246.47    221.39 
## 
## The following fit components were averaged over the 20 model fits:
## 
##   stats linear.predictors
```

```
bmi_3_OR
```

```
##     Effect Lower 0.95 Upper 0.95 
##       6.73       2.29      19.76
```

Here we show the multivariable models for BMI as suggested by the DAG. Here we show the multivariable odds ratio for outpatient visit or worse of an adult with a BMI of 35 compared to an adult with a BMI of 25.

```
bmi_1_aOR <- round(summary(fit.mult.impute(
  logistic_outpatient ~ rcs(bmi, 4) + rcs(age, 4),
  lrm,
  imputation_outcomes,
  data = df_outcome %>% filter(excluded == "Included"))
)[2, c(4,6,7)], 2)
```

```
## 
## Variance Inflation Factors Due to Imputation:
## 
## Intercept       bmi      bmi'     bmi''       age      age'     age'' 
##      1.23      1.30      1.27      1.25      1.01      1.00      1.00 
## 
## Rate of Missing Information:
## 
## Intercept       bmi      bmi'     bmi''       age      age'     age'' 
##      0.19      0.23      0.21      0.20      0.01      0.00      0.00 
## 
## d.f. for t-distribution for Tests of Single Coefficients:
## 
##  Intercept        bmi       bmi'      bmi''        age       age'      age'' 
##     535.90     352.14     428.28     464.73  453699.80 3439184.02 4181480.23 
## 
## The following fit components were averaged over the 20 model fits:
## 
##   stats linear.predictors
```

```
bmi_1_aOR
```

```
##     Effect Lower 0.95 Upper 0.95 
##       1.94       1.48       2.53
```

Here we show the multivariable odds ratio for hospital admission or worse of an adult with a BMI of 35 compared to an adult with a BMI of 25.

```
bmi_2_aOR <- round(summary(fit.mult.impute(
  logistic_inpatient ~ rcs(bmi, 4) + rcs(age, 4),
  lrm,
  imputation_outcomes,
  data = df_outcome %>% filter(excluded == "Included"))
)[2, c(4,6,7)], 2)
```

```
## 
## Variance Inflation Factors Due to Imputation:
## 
## Intercept       bmi      bmi'     bmi''       age      age'     age'' 
##      1.41      1.53      1.53      1.52      1.01      1.00      1.00 
## 
## Rate of Missing Information:
## 
## Intercept       bmi      bmi'     bmi''       age      age'     age'' 
##      0.29      0.35      0.35      0.34      0.01      0.00      0.00 
## 
## d.f. for t-distribution for Tests of Single Coefficients:
## 
##  Intercept        bmi       bmi'      bmi''        age       age'      age'' 
##     225.54     156.72     157.87     161.38  581848.57 1177051.97 1174639.04 
## 
## The following fit components were averaged over the 20 model fits:
## 
##   stats linear.predictors
```

```
bmi_2_aOR
```

```
##     Effect Lower 0.95 Upper 0.95 
##       2.54       1.64       3.95
```

Here we show the multivariable odds ratio for intensive care admission or death of an adult with a BMI of 35 compared to an adult with a BMI of 25.

```
bmi_3_aOR <- round(summary(fit.mult.impute(
  logistic_icu ~ rcs(bmi, 4) + rcs(age, 4),
  lrm,
  imputation_outcomes,
  data = df_outcome %>% filter(excluded == "Included"))
)[2, c(4,6,7)], 2)
```

```
## 
## Variance Inflation Factors Due to Imputation:
## 
## Intercept       bmi      bmi'     bmi''       age      age'     age'' 
##      1.12      1.29      1.37      1.40      1.00      1.00      1.00 
## 
## Rate of Missing Information:
## 
## Intercept       bmi      bmi'     bmi''       age      age'     age'' 
##      0.10      0.22      0.27      0.28      0.00      0.00      0.00 
## 
## d.f. for t-distribution for Tests of Single Coefficients:
## 
##  Intercept        bmi       bmi'      bmi''        age       age'      age'' 
##    1761.02     380.83     259.92     236.57 9065634.72 7012827.70 5934298.06 
## 
## The following fit components were averaged over the 20 model fits:
## 
##   stats linear.predictors
```

```
bmi_3_aOR
```

```
##     Effect Lower 0.95 Upper 0.95 
##       5.20       1.71      15.84
```

Here we show a figure that displays the OR of each outcome as a function of BMI.

```
plot_OR_bmi <- Predict(
  fit.mult.impute(
    logistic_icu ~ rcs(bmi, 4) + rcs(age, 4),
    lrm,
    imputation_outcomes,
    data = df_outcome %>% filter(excluded == "Included")
  ),
  bmi,
  age,
  ref.zero = TRUE,
  fun = exp
) %>%
  mutate(outcome = "ICU or death") %>%
  union_all(
    Predict(
      fit.mult.impute(
        logistic_inpatient ~ rcs(bmi, 4) + rcs(age, 4),
        lrm,
        imputation_outcomes,
        data = df_outcome %>% filter(excluded == "Included")
      ),
      bmi,
      age,
      ref.zero = TRUE,
      fun = exp
    ) %>%
      mutate(outcome = "Hospital admission or worse")
  ) %>%
  union_all(
    Predict(
      fit.mult.impute(
        logistic_outpatient ~ rcs(bmi, 4) + rcs(age, 4),
        lrm,
        imputation_outcomes,
        data = df_outcome %>% filter(excluded == "Included")
      ),
      bmi,
      age,
      ref.zero = TRUE,
      fun = exp
    ) %>%
      mutate(outcome = "Outpatient visit or worse")
  ) %>%
  as.data.frame() %>%
  mutate(
    outcome = factor(
      outcome,
      levels = c(
        "Outpatient visit or worse",
        "Hospital admission or worse",
        "ICU or death"
      ))
  ) %>%
  filter(age >= 60, age < 60.2) %>%
  ggplot(aes(x = bmi, y = yhat, color = outcome, fill = outcome)) +
  geom_line() +
  geom_ribbon(aes(ymin = lower, ymax = upper, color = NULL), alpha = .3) +
  geom_hline(yintercept = 1, lty = 2) +
  scale_color_manual(values = nejm_palette[c(1, 2, 4)]) +
  scale_fill_manual(values = nejm_palette[c(1, 2, 4)]) +
  scale_y_log10(
    breaks = c(0.25, 1, 4, 12, 32),
    labels = c("0.25", "1", "4", "12", "32"),
    minor_break = NULL,
    limits = c(0.000001, 512)
  ) +
  annotation_logticks(side = "l") +
  coord_cartesian(ylim = c(0.1, 34), xlim = c(18, 42)) +
  labs(
    x = "BMI",
    y = "Odds ratio of outcome \ncompared to person with a BMI of 25") +
  theme_bw() +
  theme(
    legend.title = element_blank(),
    legend.position = "bottom")
```

```
## 
## Variance Inflation Factors Due to Imputation:
## 
## Intercept       bmi      bmi'     bmi''       age      age'     age'' 
##      1.12      1.29      1.37      1.40      1.00      1.00      1.00 
## 
## Rate of Missing Information:
## 
## Intercept       bmi      bmi'     bmi''       age      age'     age'' 
##      0.10      0.22      0.27      0.28      0.00      0.00      0.00 
## 
## d.f. for t-distribution for Tests of Single Coefficients:
## 
##  Intercept        bmi       bmi'      bmi''        age       age'      age'' 
##    1761.02     380.83     259.92     236.57 9065634.72 7012827.70 5934298.06 
## 
## The following fit components were averaged over the 20 model fits:
## 
##   stats linear.predictors 
## 
## 
## Variance Inflation Factors Due to Imputation:
## 
## Intercept       bmi      bmi'     bmi''       age      age'     age'' 
##      1.41      1.53      1.53      1.52      1.01      1.00      1.00 
## 
## Rate of Missing Information:
## 
## Intercept       bmi      bmi'     bmi''       age      age'     age'' 
##      0.29      0.35      0.35      0.34      0.01      0.00      0.00 
## 
## d.f. for t-distribution for Tests of Single Coefficients:
## 
##  Intercept        bmi       bmi'      bmi''        age       age'      age'' 
##     225.54     156.72     157.87     161.38  581848.57 1177051.97 1174639.04 
## 
## The following fit components were averaged over the 20 model fits:
## 
##   stats linear.predictors 
## 
## 
## Variance Inflation Factors Due to Imputation:
## 
## Intercept       bmi      bmi'     bmi''       age      age'     age'' 
##      1.23      1.30      1.27      1.25      1.01      1.00      1.00 
## 
## Rate of Missing Information:
## 
## Intercept       bmi      bmi'     bmi''       age      age'     age'' 
##      0.19      0.23      0.21      0.20      0.01      0.00      0.00 
## 
## d.f. for t-distribution for Tests of Single Coefficients:
## 
##  Intercept        bmi       bmi'      bmi''        age       age'      age'' 
##     535.90     352.14     428.28     464.73  453699.80 3439184.02 4181480.23 
## 
## The following fit components were averaged over the 20 model fits:
## 
##   stats linear.predictors
```

```
plot_OR_bmi
```

Here we show a different figure that shows the OR of hospital admission as a function of both age and BMI. Both age and BMI are modelled using restrcited cubic splines with four knots and a linear interaction is assumed between age and BMI.

```
plot_OR_age_bmi <- Predict(
  fit.mult.impute(
    logistic_inpatient ~ rcs(bmi, 4) + rcs(age, 4) + age  %ia% bmi,
    lrm,
    imputation_outcomes,
    data = df_outcome %>% filter(excluded == "Included")
  ),
  bmi,
  age,
  ref.zero = TRUE,
  fun = exp
) %>%
  tbl_df() %>%
  ggplot(aes(x = bmi, y = age, color = yhat)) +
  geom_point() +
  geom_contour(
    aes(color = NULL, z = yhat),
    breaks = c(0.1, 0.25, 0.5, 1, 2, 4, 8, 16, 32, 64, 128, 256, 512),
    color = "white"
  ) +
  scale_x_continuous(breaks = c(20, 25, 30, 35, 40)) +
  scale_y_continuous(breaks = c(30, 40, 50, 60, 70, 80)) +
  scale_color_viridis_c(
    breaks = c(0.1, 0.25, 0.5, 1, 2, 4, 8, 16, 32, 64, 128, 256, 512),
    labels = c(0.1, 0.25, 0.5, 1, 2, 4, 8, 16, 32, 64, 128, 256, 512),
    trans = scales::pseudo_log_trans(sigma = 0.001)
  ) +
  coord_cartesian(xlim = c(18, 42), ylim = c(28, 85)) +
  labs(x = "BMI", y = "Age") +
  theme_minimal() +
  theme(legend.title = element_blank())
```

```
## 
## Variance Inflation Factors Due to Imputation:
## 
## Intercept       bmi      bmi'     bmi''       age      age'     age'' age * bmi 
##      1.41      1.53      1.50      1.49      1.01      1.02      1.02      1.17 
## 
## Rate of Missing Information:
## 
## Intercept       bmi      bmi'     bmi''       age      age'     age'' age * bmi 
##      0.29      0.35      0.33      0.33      0.01      0.02      0.02      0.15 
## 
## d.f. for t-distribution for Tests of Single Coefficients:
## 
## Intercept       bmi      bmi'     bmi''       age      age'     age'' age * bmi 
##    224.13    157.77    171.76    174.08 108738.54  76914.77  83964.90    880.78 
## 
## The following fit components were averaged over the 20 model fits:
## 
##   stats linear.predictors
```

```
plot_OR_age_bmi
```

#### 12.4 Risk factor: Current smoking

Here we show the univariate odds ratio of outpatient visit or worse between those who smoke currently, compared to those who do not.

```
smoking_current_1_OR <- round(summary(fit.mult.impute(
  logistic_outpatient ~ smoking_current,
  lrm,
  imputation_outcomes,
  data = df_outcome %>% filter(excluded == "Included"))
)[2, c(4,6,7)], 2)
```

```
## 
## Variance Inflation Factors Due to Imputation:
## 
##       Intercept smoking_current 
##            1.01            1.11 
## 
## Rate of Missing Information:
## 
##       Intercept smoking_current 
##            0.01            0.10 
## 
## d.f. for t-distribution for Tests of Single Coefficients:
## 
##       Intercept smoking_current 
##       286570.05         1996.42 
## 
## The following fit components were averaged over the 20 model fits:
## 
##   stats linear.predictors
```

```
smoking_current_1_OR
```

```
##     Effect Lower 0.95 Upper 0.95 
##       0.78       0.55       1.09
```

Here we show the univariate odds ratio of hospital admission or worse between those who smoke currently, compared to those who do not.

```
smoking_current_2_OR <- round(summary(fit.mult.impute(
  logistic_inpatient ~ smoking_current,
  lrm,
  imputation_outcomes,
  data = df_outcome %>% filter(excluded == "Included"))
)[2, c(4,6,7)], 2)
```

```
## 
## Variance Inflation Factors Due to Imputation:
## 
##       Intercept smoking_current 
##            1.01            1.31 
## 
## Rate of Missing Information:
## 
##       Intercept smoking_current 
##            0.01            0.23 
## 
## d.f. for t-distribution for Tests of Single Coefficients:
## 
##       Intercept smoking_current 
##       149879.28          347.52 
## 
## The following fit components were averaged over the 20 model fits:
## 
##   stats linear.predictors
```

```
smoking_current_2_OR
```

```
##     Effect Lower 0.95 Upper 0.95 
##       0.33       0.14       0.76
```

Here we show the univariate odds ratio of intensive care admission or death between those who smoke currently, compared to those who do not.

```
smoking_current_3_OR <- round(summary(fit.mult.impute(
  logistic_icu ~ smoking_current,
  lrm,
  imputation_outcomes,
  data = df_outcome %>% filter(excluded == "Included"))
)[2, c(4,6,7)], 2)
```

```
## 
## Variance Inflation Factors Due to Imputation:
## 
##       Intercept smoking_current 
##            1.02            1.41 
## 
## Rate of Missing Information:
## 
##       Intercept smoking_current 
##            0.02            0.29 
## 
## d.f. for t-distribution for Tests of Single Coefficients:
## 
##       Intercept smoking_current 
##        36638.08          228.24 
## 
## The following fit components were averaged over the 20 model fits:
## 
##   stats linear.predictors
```

```
smoking_current_3_OR
```

```
##     Effect Lower 0.95 Upper 0.95 
##       0.32       0.04       2.44
```

Here we show the multivariable adjusted odds ratio of outpatient visit or worse between those who smoke currently, compared to those who do not, as suggested by the DAG.

```
smoking_current_1_aOR <- round(summary(fit.mult.impute(
  logistic_outpatient ~ smoking_current + rcs(age, 4) + smoking_prior,
  lrm,
  imputation_outcomes,
  data = df_outcome %>% filter(excluded == "Included"))
)[2, c(4,6,7)], 2)
```

```
## 
## Variance Inflation Factors Due to Imputation:
## 
##       Intercept smoking_current             age            age'           age'' 
##            1.00            1.11            1.00            1.00            1.00 
##   smoking_prior 
##            1.51 
## 
## Rate of Missing Information:
## 
##       Intercept smoking_current             age            age'           age'' 
##            0.00            0.10            0.00            0.00            0.00 
##   smoking_prior 
##            0.34 
## 
## d.f. for t-distribution for Tests of Single Coefficients:
## 
##       Intercept smoking_current             age            age'           age'' 
##     37889524.04         1903.61      4382510.62     10596801.58      8500856.04 
##   smoking_prior 
##          167.37 
## 
## The following fit components were averaged over the 20 model fits:
## 
##   stats linear.predictors
```

```
smoking_current_1_aOR
```

```
##     Effect Lower 0.95 Upper 0.95 
##       0.73       0.50       1.05
```

Here we show the multivariable adjusted odds ratio of hospital admission or worse between those who smoke currently, compared to those who do not, as suggested by the DAG.

```
smoking_current_2_aOR <- round(summary(fit.mult.impute(
  logistic_inpatient ~ smoking_current + rcs(age, 4) + smoking_prior,
  lrm,
  imputation_outcomes,
  data = df_outcome %>% filter(excluded == "Included"))
)[2, c(4,6,7)], 2)
```

```
## 
## Variance Inflation Factors Due to Imputation:
## 
##       Intercept smoking_current             age            age'           age'' 
##            1.00            1.30            1.00            1.00            1.00 
##   smoking_prior 
##            2.02 
## 
## Rate of Missing Information:
## 
##       Intercept smoking_current             age            age'           age'' 
##            0.00            0.23            0.00            0.00            0.00 
##   smoking_prior 
##            0.50 
## 
## d.f. for t-distribution for Tests of Single Coefficients:
## 
##       Intercept smoking_current             age            age'           age'' 
##    143067142.72          365.11     10067682.74     19420999.28      8865854.34 
##   smoking_prior 
##           74.55 
## 
## The following fit components were averaged over the 20 model fits:
## 
##   stats linear.predictors
```

```
smoking_current_2_aOR
```

```
##     Effect Lower 0.95 Upper 0.95 
##       0.41       0.17       0.98
```

Here we show the multivariable adjusted odds ratio of intensive care admission or death between those who smoke currently, compared to those who do not, as suggested by the DAG.

```
smoking_current_3_aOR <- round(summary(fit.mult.impute(
  logistic_icu ~ smoking_current + rcs(age, 4) + smoking_prior,
  lrm,
  imputation_outcomes,
  data = df_outcome %>% filter(excluded == "Included"))
)[2, c(4,6,7)], 2)
```

```
## 
## Variance Inflation Factors Due to Imputation:
## 
##       Intercept smoking_current             age            age'           age'' 
##            1.00            1.48            1.00            1.00            1.00 
##   smoking_prior 
##            2.89 
## 
## Rate of Missing Information:
## 
##       Intercept smoking_current             age            age'           age'' 
##            0.00            0.32            0.00            0.00            0.00 
##   smoking_prior 
##            0.65 
## 
## d.f. for t-distribution for Tests of Single Coefficients:
## 
##       Intercept smoking_current             age            age'           age'' 
##      5938074.19          181.50      7927509.83      1117552.44       857997.67 
##   smoking_prior 
##           44.43 
## 
## The following fit components were averaged over the 20 model fits:
## 
##   stats linear.predictors
```

```
smoking_current_3_aOR
```

```
##     Effect Lower 0.95 Upper 0.95 
##       0.56       0.07       4.77
```

#### 12.5 Risk factor: Prior smoking

Here we show the univariate odds ratio of outpatient visit or worse between those who have a prior smoking history, and those who have never smoked.

```
smoking_prior_1_OR <- round(summary(fit.mult.impute(
  logistic_outpatient ~ smoking_prior,
  lrm,
  imputation_outcomes,
  data = df_outcome %>% filter(excluded == "Included"))
)[2, c(4,6,7)], 2)
```

```
## 
## Variance Inflation Factors Due to Imputation:
## 
##     Intercept smoking_prior 
##          1.27          1.46 
## 
## Rate of Missing Information:
## 
##     Intercept smoking_prior 
##          0.21          0.32 
## 
## d.f. for t-distribution for Tests of Single Coefficients:
## 
##     Intercept smoking_prior 
##        412.43        190.00 
## 
## The following fit components were averaged over the 20 model fits:
## 
##   stats linear.predictors
```

```
smoking_prior_1_OR
```

```
##     Effect Lower 0.95 Upper 0.95 
##       1.87       1.52       2.30
```

Here we show the univariate odds ratio of hospital admission or worse between those who have a prior smoking history, and those who have never smoked.

```
smoking_prior_2_OR <- round(summary(fit.mult.impute(
  logistic_inpatient ~ smoking_prior,
  lrm,
  imputation_outcomes,
  data = df_outcome %>% filter(excluded == "Included"))
)[2, c(4,6,7)], 2)
```

```
## 
## Variance Inflation Factors Due to Imputation:
## 
##     Intercept smoking_prior 
##          1.50          1.84 
## 
## Rate of Missing Information:
## 
##     Intercept smoking_prior 
##          0.34          0.46 
## 
## d.f. for t-distribution for Tests of Single Coefficients:
## 
##     Intercept smoking_prior 
##        169.28         91.70 
## 
## The following fit components were averaged over the 20 model fits:
## 
##   stats linear.predictors
```

```
smoking_prior_2_OR
```

```
##     Effect Lower 0.95 Upper 0.95 
##       2.07       1.45       2.96
```

Here we show the univariate odds ratio of intensive care admission or worse between those who have a prior smoking history, and those who have never smoked.

```
smoking_prior_3_OR <- round(summary(fit.mult.impute(
  logistic_icu ~ smoking_prior,
  lrm,
  imputation_outcomes,
  data = df_outcome %>% filter(excluded == "Included"))
)[2, c(4,6,7)], 2)
```

```
## 
## Variance Inflation Factors Due to Imputation:
## 
##     Intercept smoking_prior 
##          1.75          2.73 
## 
## Rate of Missing Information:
## 
##     Intercept smoking_prior 
##          0.43          0.63 
## 
## d.f. for t-distribution for Tests of Single Coefficients:
## 
##     Intercept smoking_prior 
##        102.68         47.38 
## 
## The following fit components were averaged over the 20 model fits:
## 
##   stats linear.predictors
```

```
smoking_prior_3_OR
```

```
##     Effect Lower 0.95 Upper 0.95 
##       1.36       0.53       3.48
```

Here we show the multivariable adjusted odds ratio of outpatient visit or worse between those who have a prior smoking history, and those who have never smoked, as suggested by the DAG.

```
smoking_prior_1_aOR <- round(summary(
  fit.mult.impute(
    logistic_outpatient ~ smoking_prior + rcs(age, 4) + sex,
    lrm,
    imputation_outcomes,
    data = df_outcome %>% filter(excluded == "Included")
  )
)[2, c(4, 6, 7)], 2)
```

```
## 
## Variance Inflation Factors Due to Imputation:
## 
##     Intercept smoking_prior           age          age'         age'' 
##          1.00          1.55          1.00          1.00          1.00 
##           sex 
##          1.01 
## 
## Rate of Missing Information:
## 
##     Intercept smoking_prior           age          age'         age'' 
##          0.00          0.35          0.00          0.00          0.00 
##           sex 
##          0.01 
## 
## d.f. for t-distribution for Tests of Single Coefficients:
## 
##     Intercept smoking_prior           age          age'         age'' 
##   37638025.06        152.38    2892859.30    8497502.89    7389722.16 
##           sex 
##     232335.63 
## 
## The following fit components were averaged over the 20 model fits:
## 
##   stats linear.predictors
```

```
smoking_prior_1_aOR
```

```
##     Effect Lower 0.95 Upper 0.95 
##       1.38       1.10       1.74
```

Here we show the multivariable adjusted odds ratio of hospital admission or worse between those who have a prior smoking history, and those who have never smoked, as suggested by the DAG.

```
smoking_prior_2_aOR <- round(summary(
  fit.mult.impute(
    logistic_inpatient ~ smoking_prior + rcs(age, 4) + sex,
    lrm,
    imputation_outcomes,
    data = df_outcome %>% filter(excluded == "Included")
  )
)[2, c(4, 6, 7)], 2)
```

```
## 
## Variance Inflation Factors Due to Imputation:
## 
##     Intercept smoking_prior           age          age'         age'' 
##          1.00          2.01          1.00          1.00          1.00 
##           sex 
##          1.01 
## 
## Rate of Missing Information:
## 
##     Intercept smoking_prior           age          age'         age'' 
##          0.00          0.50          0.00          0.00          0.00 
##           sex 
##          0.01 
## 
## d.f. for t-distribution for Tests of Single Coefficients:
## 
##     Intercept smoking_prior           age          age'         age'' 
##  270524219.02         75.03    4835301.35  158591737.30   59729059.99 
##           sex 
##     112250.25 
## 
## The following fit components were averaged over the 20 model fits:
## 
##   stats linear.predictors
```

```
smoking_prior_2_aOR
```

```
##     Effect Lower 0.95 Upper 0.95 
##       1.23       0.81       1.87
```

Here we show the multivariable adjusted odds ratio of intensive care admission or death between those who have a prior smoking history, and those who have never smoked, as suggested by the DAG.

```
smoking_prior_3_aOR <- round(summary(
  fit.mult.impute(
    logistic_icu ~ smoking_prior + rcs(age, 4) + sex,
    lrm,
    imputation_outcomes,
    data = df_outcome %>% filter(excluded == "Included")
  )
)[2, c(4, 6, 7)], 2)
```

```
## 
## Variance Inflation Factors Due to Imputation:
## 
##     Intercept smoking_prior           age          age'         age'' 
##          1.00          2.84          1.00          1.00          1.00 
##           sex 
##          1.03 
## 
## Rate of Missing Information:
## 
##     Intercept smoking_prior           age          age'         age'' 
##          0.00          0.65          0.00          0.00          0.00 
##           sex 
##          0.03 
## 
## d.f. for t-distribution for Tests of Single Coefficients:
## 
##     Intercept smoking_prior           age          age'         age'' 
##   33736774.57         45.24   53503993.97    4757878.09    3418769.28 
##           sex 
##      28586.07 
## 
## The following fit components were averaged over the 20 model fits:
## 
##   stats linear.predictors
```

```
smoking_prior_3_aOR
```

```
##     Effect Lower 0.95 Upper 0.95 
##       0.67       0.25       1.80
```

#### 12.6 Risk factor: Diabetes

Defined as the presence of an ICD-10 diagnosis of E08-E13 (diabetes mellitus) AND a filled prescription of antidiabetic drugs (ATC A10 and subgroups) within 13 prior to diagnosis

```
diabetes_1_OR <- round(summary(fit.mult.impute(
  logistic_outpatient ~ diabetes_retrospective,
  lrm,
  imputation_outcomes,
  data = df_outcome %>% filter(excluded == "Included"))
)[2, c(4,6,7)], 2)
```

```
## 
## Variance Inflation Factors Due to Imputation:
## 
##              Intercept diabetes_retrospective 
##                      1                      1 
## 
## Rate of Missing Information:
## 
##              Intercept diabetes_retrospective 
##                      0                      0 
## 
## d.f. for t-distribution for Tests of Single Coefficients:
## 
##              Intercept diabetes_retrospective 
##           1.291255e+26           1.515122e+28 
## 
## The following fit components were averaged over the 20 model fits:
## 
##   stats linear.predictors
```

```
diabetes_1_OR
```

```
##     Effect Lower 0.95 Upper 0.95 
##       3.35       2.42       4.64
```

```
diabetes_2_OR <- round(summary(fit.mult.impute(
  logistic_inpatient ~ diabetes_retrospective,
  lrm,
  imputation_outcomes,
  data = df_outcome %>% filter(excluded == "Included"))
)[2, c(4,6,7)], 2)
```

```
## 
## Variance Inflation Factors Due to Imputation:
## 
##              Intercept diabetes_retrospective 
##                      1                      1 
## 
## Rate of Missing Information:
## 
##              Intercept diabetes_retrospective 
##                      0                      0 
## 
## d.f. for t-distribution for Tests of Single Coefficients:
## 
##              Intercept diabetes_retrospective 
##           4.929343e+25           1.493018e+28 
## 
## The following fit components were averaged over the 20 model fits:
## 
##   stats linear.predictors
```

```
diabetes_2_OR
```

```
##     Effect Lower 0.95 Upper 0.95 
##       4.39       2.91       6.63
```

```
diabetes_3_OR <- round(summary(fit.mult.impute(
  logistic_icu ~ diabetes_retrospective,
  lrm,
  imputation_outcomes,
  data = df_outcome %>% filter(excluded == "Included"))
)[2, c(4,6,7)], 2)
```

```
## 
## Variance Inflation Factors Due to Imputation:
## 
##              Intercept diabetes_retrospective 
##                      1                      1 
## 
## Rate of Missing Information:
## 
##              Intercept diabetes_retrospective 
##                      0                      0 
## 
## d.f. for t-distribution for Tests of Single Coefficients:
## 
##              Intercept diabetes_retrospective 
##           1.360772e+26           1.323750e+29 
## 
## The following fit components were averaged over the 20 model fits:
## 
##   stats linear.predictors
```

```
diabetes_3_OR
```

```
##     Effect Lower 0.95 Upper 0.95 
##       6.67       3.28      13.56
```

```
diabetes_1_aOR <- round(summary(fit.mult.impute(
  logistic_outpatient ~ diabetes_retrospective + rcs(age, 4) + rcs(bmi, 4),
  lrm,
  imputation_outcomes,
  data = df_outcome %>% filter(excluded == "Included"))
)[2, c(4,6,7)], 2)
```

```
## 
## Variance Inflation Factors Due to Imputation:
## 
##              Intercept diabetes_retrospective                    age 
##                   1.23                   1.01                   1.01 
##                   age'                  age''                    bmi 
##                   1.00                   1.00                   1.30 
##                   bmi'                  bmi'' 
##                   1.26                   1.25 
## 
## Rate of Missing Information:
## 
##              Intercept diabetes_retrospective                    age 
##                   0.19                   0.01                   0.01 
##                   age'                  age''                    bmi 
##                   0.00                   0.00                   0.23 
##                   bmi'                  bmi'' 
##                   0.21                   0.20 
## 
## d.f. for t-distribution for Tests of Single Coefficients:
## 
##              Intercept diabetes_retrospective                    age 
##                 546.62               98482.27              435030.06 
##                   age'                  age''                    bmi 
##             3475158.62             4623921.60                 357.61 
##                   bmi'                  bmi'' 
##                 438.89                 477.53 
## 
## The following fit components were averaged over the 20 model fits:
## 
##   stats linear.predictors
```

```
diabetes_1_aOR
```

```
##     Effect Lower 0.95 Upper 0.95 
##       1.64       1.14       2.34
```

```
diabetes_2_aOR <- round(summary(fit.mult.impute(
  logistic_inpatient ~ diabetes_retrospective + rcs(age, 4) + rcs(bmi, 4),
  lrm,
  imputation_outcomes,
  data = df_outcome %>% filter(excluded == "Included"))
)[2, c(4,6,7)], 2)
```

```
## 
## Variance Inflation Factors Due to Imputation:
## 
##              Intercept diabetes_retrospective                    age 
##                   1.41                   1.03                   1.01 
##                   age'                  age''                    bmi 
##                   1.00                   1.00                   1.53 
##                   bmi'                  bmi'' 
##                   1.53                   1.52 
## 
## Rate of Missing Information:
## 
##              Intercept diabetes_retrospective                    age 
##                   0.29                   0.03                   0.01 
##                   age'                  age''                    bmi 
##                   0.00                   0.00                   0.35 
##                   bmi'                  bmi'' 
##                   0.35                   0.34 
## 
## d.f. for t-distribution for Tests of Single Coefficients:
## 
##              Intercept diabetes_retrospective                    age 
##                 227.95               28696.74              603543.90 
##                   age'                  age''                    bmi 
##             1285711.12             1330236.10                 158.19 
##                   bmi'                  bmi'' 
##                 159.06                 162.47 
## 
## The following fit components were averaged over the 20 model fits:
## 
##   stats linear.predictors
```

```
diabetes_2_aOR
```

```
##     Effect Lower 0.95 Upper 0.95 
##       1.53       0.94       2.48
```

```
diabetes_3_aOR <- round(summary(fit.mult.impute(
  logistic_icu ~ diabetes_retrospective + rcs(age, 4) + rcs(bmi, 4),
  lrm,
  imputation_outcomes,
  data = df_outcome %>% filter(excluded == "Included"))
)[2, c(4,6,7)], 2)
```

```
## 
## Variance Inflation Factors Due to Imputation:
## 
##              Intercept diabetes_retrospective                    age 
##                   1.12                   1.03                   1.00 
##                   age'                  age''                    bmi 
##                   1.00                   1.00                   1.29 
##                   bmi'                  bmi'' 
##                   1.38                   1.40 
## 
## Rate of Missing Information:
## 
##              Intercept diabetes_retrospective                    age 
##                   0.10                   0.03                   0.00 
##                   age'                  age''                    bmi 
##                   0.00                   0.00                   0.23 
##                   bmi'                  bmi'' 
##                   0.27                   0.29 
## 
## d.f. for t-distribution for Tests of Single Coefficients:
## 
##              Intercept diabetes_retrospective                    age 
##                1725.46               23499.27            11846856.21 
##                   age'                  age''                    bmi 
##             9366604.46             7849332.59                 367.37 
##                   bmi'                  bmi'' 
##                 253.70                 231.68 
## 
## The following fit components were averaged over the 20 model fits:
## 
##   stats linear.predictors
```

```
diabetes_3_aOR
```

```
##     Effect Lower 0.95 Upper 0.95 
##       1.89       0.87       4.08
```

#### 12.7 Risk factor: Hypertension

Defined as the presence of an ICD-10 diagnosis of I10-I19 AND a filled prescription within the last 13 months for drugs with ATC numbers C02 (antihypertensives), C07 (beta blocking agents), C08 (calcium channel blockers), or C09 (agents acting on the renin-angiotensin system).

```
htn_1_OR <- round(summary(fit.mult.impute(
  logistic_outpatient ~ htn_retrospective,
  lrm,
  imputation_outcomes,
  data = df_outcome %>% filter(excluded == "Included"))
)[2, c(4,6,7)], 2)
```

```
## 
## Variance Inflation Factors Due to Imputation:
## 
##         Intercept htn_retrospective 
##                 1                 1 
## 
## Rate of Missing Information:
## 
##         Intercept htn_retrospective 
##                 0                 0 
## 
## d.f. for t-distribution for Tests of Single Coefficients:
## 
##         Intercept htn_retrospective 
##      4.898365e+25      8.880237e+26 
## 
## The following fit components were averaged over the 20 model fits:
## 
##   stats linear.predictors
```

```
htn_1_OR
```

```
##     Effect Lower 0.95 Upper 0.95 
##       3.24       2.65       3.98
```

```
htn_2_OR <- round(summary(fit.mult.impute(
  logistic_inpatient ~ htn_retrospective,
  lrm,
  imputation_outcomes,
  data = df_outcome %>% filter(excluded == "Included"))
)[2, c(4,6,7)], 2)
```

```
## 
## Variance Inflation Factors Due to Imputation:
## 
##         Intercept htn_retrospective 
##                 1                 1 
## 
## Rate of Missing Information:
## 
##         Intercept htn_retrospective 
##                 0                 0 
## 
## d.f. for t-distribution for Tests of Single Coefficients:
## 
##         Intercept htn_retrospective 
##      1.112683e+26      7.244880e+26 
## 
## The following fit components were averaged over the 20 model fits:
## 
##   stats linear.predictors
```

```
htn_2_OR
```

```
##     Effect Lower 0.95 Upper 0.95 
##       6.00       4.57       7.88
```

```
htn_3_OR <- round(summary(fit.mult.impute(
  logistic_icu ~ htn_retrospective,
  lrm,
  imputation_outcomes,
  data = df_outcome %>% filter(excluded == "Included"))
)[2, c(4,6,7)], 2)
```

```
## 
## Variance Inflation Factors Due to Imputation:
## 
##         Intercept htn_retrospective 
##                 1                 1 
## 
## Rate of Missing Information:
## 
##         Intercept htn_retrospective 
##                 0                 0 
## 
## d.f. for t-distribution for Tests of Single Coefficients:
## 
##         Intercept htn_retrospective 
##      2.113560e+26      5.191275e+28 
## 
## The following fit components were averaged over the 20 model fits:
## 
##   stats linear.predictors
```

```
htn_3_OR
```

```
##     Effect Lower 0.95 Upper 0.95 
##       7.74       4.41      13.57
```

```
htn_1_aOR <- round(summary(fit.mult.impute(
  logistic_outpatient ~ htn_retrospective + rcs(age, 4) + rcs(bmi, 4) + smoking_current,
  lrm,
  imputation_outcomes,
  data = df_outcome %>% filter(excluded == "Included"))
)[2, c(4,6,7)], 2)
```

```
## 
## Variance Inflation Factors Due to Imputation:
## 
##         Intercept htn_retrospective               age              age' 
##              1.23              1.02              1.01              1.00 
##             age''               bmi              bmi'             bmi'' 
##              1.00              1.30              1.26              1.25 
##   smoking_current 
##              1.11 
## 
## Rate of Missing Information:
## 
##         Intercept htn_retrospective               age              age' 
##              0.19              0.02              0.01              0.00 
##             age''               bmi              bmi'             bmi'' 
##              0.00              0.23              0.21              0.20 
##   smoking_current 
##              0.10 
## 
## d.f. for t-distribution for Tests of Single Coefficients:
## 
##         Intercept htn_retrospective               age              age' 
##            543.65          36688.27         470050.17        3020964.53 
##             age''               bmi              bmi'             bmi'' 
##        3923946.98            355.99            435.72            473.58 
##   smoking_current 
##           2062.73 
## 
## The following fit components were averaged over the 20 model fits:
## 
##   stats linear.predictors
```

```
htn_1_aOR
```

```
##     Effect Lower 0.95 Upper 0.95 
##       1.13       0.88       1.46
```

```
htn_2_aOR <- round(summary(fit.mult.impute(
  logistic_inpatient ~ htn_retrospective + rcs(age, 4) + rcs(bmi, 4) + smoking_current,
  lrm,
  imputation_outcomes,
  data = df_outcome %>% filter(excluded == "Included"))
)[2, c(4,6,7)], 2)
```

```
## 
## Variance Inflation Factors Due to Imputation:
## 
##         Intercept htn_retrospective               age              age' 
##              1.40              1.02              1.01              1.00 
##             age''               bmi              bmi'             bmi'' 
##              1.00              1.52              1.52              1.51 
##   smoking_current 
##              1.28 
## 
## Rate of Missing Information:
## 
##         Intercept htn_retrospective               age              age' 
##              0.29              0.02              0.01              0.00 
##             age''               bmi              bmi'             bmi'' 
##              0.00              0.34              0.34              0.34 
##   smoking_current 
##              0.22 
## 
## d.f. for t-distribution for Tests of Single Coefficients:
## 
##         Intercept htn_retrospective               age              age' 
##            232.80          41049.37         403587.93         884988.94 
##             age''               bmi              bmi'             bmi'' 
##        1006633.45            160.97            163.90            168.18 
##   smoking_current 
##            407.09 
## 
## The following fit components were averaged over the 20 model fits:
## 
##   stats linear.predictors
```

```
htn_2_aOR
```

```
##     Effect Lower 0.95 Upper 0.95 
##       1.21       0.85       1.72
```

```
htn_3_aOR <- round(summary(fit.mult.impute(
  logistic_icu ~ htn_retrospective + rcs(age, 4) + rcs(bmi, 4) + smoking_current,
  lrm,
  imputation_outcomes,
  data = df_outcome %>% filter(excluded == "Included"))
)[2, c(4,6,7)], 2)
```

```
## 
## Variance Inflation Factors Due to Imputation:
## 
##         Intercept htn_retrospective               age              age' 
##              1.11              1.03              1.00              1.00 
##             age''               bmi              bmi'             bmi'' 
##              1.00              1.28              1.37              1.39 
##   smoking_current 
##              1.43 
## 
## Rate of Missing Information:
## 
##         Intercept htn_retrospective               age              age' 
##              0.10              0.03              0.00              0.00 
##             age''               bmi              bmi'             bmi'' 
##              0.00              0.22              0.27              0.28 
##   smoking_current 
##              0.30 
## 
## d.f. for t-distribution for Tests of Single Coefficients:
## 
##         Intercept htn_retrospective               age              age' 
##           1796.73          25017.66        6569966.53        6833012.27 
##             age''               bmi              bmi'             bmi'' 
##        7170365.10            389.04            264.64            240.40 
##   smoking_current 
##            210.83 
## 
## The following fit components were averaged over the 20 model fits:
## 
##   stats linear.predictors
```

```
htn_3_aOR
```

```
##     Effect Lower 0.95 Upper 0.95 
##       1.37       0.71       2.64
```

#### 12.8 Risk factor: Ischemic heart disease

Defined as the presence of an ICD-10 diagnosis of I20., I21., I23, I24, I25 AND a filled prescription within the last 13 months for drugs with ATC numbers B01AC (platelet aggregation inhibitors excl. heparin), C10 (lipid modifying agents), or C01DA (organic nitrates)

```
ihd_1_OR <- round(summary(fit.mult.impute(
  logistic_outpatient ~ ihd_retrospective,
  lrm,
  imputation_outcomes,
  data = df_outcome %>% filter(excluded == "Included"))
)[2, c(4,6,7)], 2)
```

```
## 
## Variance Inflation Factors Due to Imputation:
## 
##         Intercept ihd_retrospective 
##                 1                 1 
## 
## Rate of Missing Information:
## 
##         Intercept ihd_retrospective 
##                 0                 0 
## 
## d.f. for t-distribution for Tests of Single Coefficients:
## 
##         Intercept ihd_retrospective 
##      1.474405e+25      3.681088e+27 
## 
## The following fit components were averaged over the 20 model fits:
## 
##   stats linear.predictors
```

```
ihd_1_OR
```

```
##     Effect Lower 0.95 Upper 0.95 
##       4.93       3.56       6.81
```

```
ihd_2_OR <- round(summary(fit.mult.impute(
  logistic_inpatient ~ ihd_retrospective,
  lrm,
  imputation_outcomes,
  data = df_outcome %>% filter(excluded == "Included"))
)[2, c(4,6,7)], 2)
```

```
## 
## Variance Inflation Factors Due to Imputation:
## 
##         Intercept ihd_retrospective 
##                 1                 1 
## 
## Rate of Missing Information:
## 
##         Intercept ihd_retrospective 
##                 0                 0 
## 
## d.f. for t-distribution for Tests of Single Coefficients:
## 
##         Intercept ihd_retrospective 
##      8.923598e+24               Inf 
## 
## The following fit components were averaged over the 20 model fits:
## 
##   stats linear.predictors
```

```
ihd_2_OR
```

```
##     Effect Lower 0.95 Upper 0.95 
##       8.24       5.67      11.98
```

```
ihd_3_OR <- round(summary(fit.mult.impute(
  logistic_icu ~ ihd_retrospective,
  lrm,
  imputation_outcomes,
  data = df_outcome %>% filter(excluded == "Included"))
)[2, c(4,6,7)], 2)
```

```
## 
## Variance Inflation Factors Due to Imputation:
## 
##         Intercept ihd_retrospective 
##                 1                 1 
## 
## Rate of Missing Information:
## 
##         Intercept ihd_retrospective 
##                 0                 0 
## 
## d.f. for t-distribution for Tests of Single Coefficients:
## 
##         Intercept ihd_retrospective 
##      3.088186e+25      2.975754e+30 
## 
## The following fit components were averaged over the 20 model fits:
## 
##   stats linear.predictors
```

```
ihd_3_OR
```

```
##     Effect Lower 0.95 Upper 0.95 
##       5.52       2.55      11.95
```

```
ihd_1_aOR <- round(summary(fit.mult.impute(
  logistic_outpatient ~ ihd_retrospective + rcs(age, 4) + sex + rcs(bmi, 4) +
    htn_retrospective + diabetes_retrospective + smoking_prior,
  lrm,
  imputation_outcomes,
  data = df_outcome %>% filter(excluded == "Included"))
)[2, c(4,6,7)], 2)
```

```
## 
## Variance Inflation Factors Due to Imputation:
## 
##              Intercept      ihd_retrospective                    age 
##                   1.23                   1.01                   1.01 
##                   age'                  age''                    sex 
##                   1.00                   1.00                   1.03 
##                    bmi                   bmi'                  bmi'' 
##                   1.30                   1.27                   1.26 
##      htn_retrospective diabetes_retrospective          smoking_prior 
##                   1.02                   1.01                   1.58 
## 
## Rate of Missing Information:
## 
##              Intercept      ihd_retrospective                    age 
##                   0.19                   0.01                   0.01 
##                   age'                  age''                    sex 
##                   0.00                   0.00                   0.02 
##                    bmi                   bmi'                  bmi'' 
##                   0.23                   0.21                   0.21 
##      htn_retrospective diabetes_retrospective          smoking_prior 
##                   0.02                   0.01                   0.37 
## 
## d.f. for t-distribution for Tests of Single Coefficients:
## 
##              Intercept      ihd_retrospective                    age 
##                 538.44              331955.70              143044.32 
##                   age'                  age''                    sex 
##              815479.11             1215708.91               30528.89 
##                    bmi                   bmi'                  bmi'' 
##                 350.06                 411.78                 436.33 
##      htn_retrospective diabetes_retrospective          smoking_prior 
##               45868.08              133482.38                 141.44 
## 
## The following fit components were averaged over the 20 model fits:
## 
##   stats linear.predictors
```

```
ihd_1_aOR
```

```
##     Effect Lower 0.95 Upper 0.95 
##       1.41       0.96       2.08
```

```
ihd_2_aOR <- round(summary(fit.mult.impute(
  logistic_inpatient ~ ihd_retrospective + rcs(age, 4) + sex + rcs(bmi, 4) +
    htn_retrospective + diabetes_retrospective + smoking_prior,
  lrm,
  imputation_outcomes,
  data = df_outcome %>% filter(excluded == "Included"))
)[2, c(4,6,7)], 2)
```

```
## 
## Variance Inflation Factors Due to Imputation:
## 
##              Intercept      ihd_retrospective                    age 
##                   1.40                   1.02                   1.01 
##                   age'                  age''                    sex 
##                   1.00                   1.00                   1.03 
##                    bmi                   bmi'                  bmi'' 
##                   1.53                   1.53                   1.52 
##      htn_retrospective diabetes_retrospective          smoking_prior 
##                   1.01                   1.02                   2.04 
## 
## Rate of Missing Information:
## 
##              Intercept      ihd_retrospective                    age 
##                   0.29                   0.02                   0.01 
##                   age'                  age''                    sex 
##                   0.00                   0.00                   0.03 
##                    bmi                   bmi'                  bmi'' 
##                   0.35                   0.35                   0.34 
##      htn_retrospective diabetes_retrospective          smoking_prior 
##                   0.01                   0.02                   0.51 
## 
## d.f. for t-distribution for Tests of Single Coefficients:
## 
##              Intercept      ihd_retrospective                    age 
##                 230.27               33366.22              320940.10 
##                   age'                  age''                    sex 
##             1041445.95             1148447.59               27980.77 
##                    bmi                   bmi'                  bmi'' 
##                 158.14                 159.30                 163.16 
##      htn_retrospective diabetes_retrospective          smoking_prior 
##              111596.43               44298.76                  72.91 
## 
## The following fit components were averaged over the 20 model fits:
## 
##   stats linear.predictors
```

```
ihd_2_aOR
```

```
##     Effect Lower 0.95 Upper 0.95 
##       1.11       0.69       1.77
```

```
ihd_3_aOR <- round(summary(fit.mult.impute(
  logistic_icu ~ ihd_retrospective + rcs(age, 4) + sex + rcs(bmi, 4) +
    htn_retrospective + diabetes_retrospective + smoking_prior,
  lrm,
  imputation_outcomes,
  data = df_outcome %>% filter(excluded == "Included"))
)[2, c(4,6,7)], 2)
```

```
## 
## Variance Inflation Factors Due to Imputation:
## 
##              Intercept      ihd_retrospective                    age 
##                   1.12                   1.07                   1.00 
##                   age'                  age''                    sex 
##                   1.00                   1.01                   1.02 
##                    bmi                   bmi'                  bmi'' 
##                   1.29                   1.37                   1.40 
##      htn_retrospective diabetes_retrospective          smoking_prior 
##                   1.03                   1.03                   3.00 
## 
## Rate of Missing Information:
## 
##              Intercept      ihd_retrospective                    age 
##                   0.11                   0.07                   0.00 
##                   age'                  age''                    sex 
##                   0.00                   0.01                   0.02 
##                    bmi                   bmi'                  bmi'' 
##                   0.22                   0.27                   0.29 
##      htn_retrospective diabetes_retrospective          smoking_prior 
##                   0.03                   0.03                   0.67 
## 
## d.f. for t-distribution for Tests of Single Coefficients:
## 
##              Intercept      ihd_retrospective                    age 
##                1703.99                4237.51             2209931.24 
##                   age'                  age''                    sex 
##              851363.67              718310.71               41683.55 
##                    bmi                   bmi'                  bmi'' 
##                 382.62                 256.11                 229.52 
##      htn_retrospective diabetes_retrospective          smoking_prior 
##               24484.81               20502.89                  42.74 
## 
## The following fit components were averaged over the 20 model fits:
## 
##   stats linear.predictors
```

```
ihd_3_aOR
```

```
##     Effect Lower 0.95 Upper 0.95 
##       0.81       0.33       2.01
```

#### 12.9 Risk factor: Chronic kidney disease

```
ckd_1_OR <- round(summary(fit.mult.impute(
  logistic_outpatient ~ ckd_retrospective,
  lrm,
  imputation_outcomes,
  data = df_outcome %>% filter(excluded == "Included"))
)[2, c(4,6,7)], 2)
```

```
## 
## Variance Inflation Factors Due to Imputation:
## 
##         Intercept ckd_retrospective 
##                 1                 1 
## 
## Rate of Missing Information:
## 
##         Intercept ckd_retrospective 
##                 0                 0 
## 
## d.f. for t-distribution for Tests of Single Coefficients:
## 
##         Intercept ckd_retrospective 
##      3.328833e+25      1.785144e+27 
## 
## The following fit components were averaged over the 20 model fits:
## 
##   stats linear.predictors
```

```
ckd_1_OR
```

```
##     Effect Lower 0.95 Upper 0.95 
##      10.87       7.42      15.94
```

```
ckd_2_OR <- round(summary(fit.mult.impute(
  logistic_inpatient ~ ckd_retrospective,
  lrm,
  imputation_outcomes,
  data = df_outcome %>% filter(excluded == "Included"))
)[2, c(4,6,7)], 2)
```

```
## 
## Variance Inflation Factors Due to Imputation:
## 
##         Intercept ckd_retrospective 
##                 1                 1 
## 
## Rate of Missing Information:
## 
##         Intercept ckd_retrospective 
##                 0                 0 
## 
## d.f. for t-distribution for Tests of Single Coefficients:
## 
##         Intercept ckd_retrospective 
##      9.866066e+24      5.070790e+26 
## 
## The following fit components were averaged over the 20 model fits:
## 
##   stats linear.predictors
```

```
ckd_2_OR
```

```
##     Effect Lower 0.95 Upper 0.95 
##      21.44      14.45      31.82
```

```
ckd_3_OR <- round(summary(fit.mult.impute(
  logistic_icu ~ ckd_retrospective,
  lrm,
  imputation_outcomes,
  data = df_outcome %>% filter(excluded == "Included"))
)[2, c(4,6,7)], 2)
```

```
## 
## Variance Inflation Factors Due to Imputation:
## 
##         Intercept ckd_retrospective 
##                 1                 1 
## 
## Rate of Missing Information:
## 
##         Intercept ckd_retrospective 
##                 0                 0 
## 
## d.f. for t-distribution for Tests of Single Coefficients:
## 
##         Intercept ckd_retrospective 
##      1.224392e+27      2.237187e+28 
## 
## The following fit components were averaged over the 20 model fits:
## 
##   stats linear.predictors
```

```
ckd_3_OR
```

```
##     Effect Lower 0.95 Upper 0.95 
##      10.95       5.34      22.49
```

```
ckd_1_aOR <- round(summary(fit.mult.impute(
  logistic_outpatient ~ ckd_retrospective + rcs(age, 4) + rcs(bmi, 4) +
    htn_retrospective + diabetes_retrospective + smoking_prior,
  lrm,
  imputation_outcomes,
  data = df_outcome %>% filter(excluded == "Included"))
)[2, c(4,6,7)], 2)
```

```
## 
## Variance Inflation Factors Due to Imputation:
## 
##              Intercept      ckd_retrospective                    age 
##                   1.23                   1.00                   1.01 
##                   age'                  age''                    bmi 
##                   1.00                   1.00                   1.30 
##                   bmi'                  bmi''      htn_retrospective 
##                   1.27                   1.26                   1.02 
## diabetes_retrospective          smoking_prior 
##                   1.01                   1.59 
## 
## Rate of Missing Information:
## 
##              Intercept      ckd_retrospective                    age 
##                   0.19                   0.00                   0.01 
##                   age'                  age''                    bmi 
##                   0.00                   0.00                   0.23 
##                   bmi'                  bmi''      htn_retrospective 
##                   0.21                   0.21                   0.02 
## diabetes_retrospective          smoking_prior 
##                   0.01                   0.37 
## 
## d.f. for t-distribution for Tests of Single Coefficients:
## 
##              Intercept      ckd_retrospective                    age 
##                 545.41             1402747.18              139127.22 
##                   age'                  age''                    bmi 
##              895867.52             1368110.86                 353.33 
##                   bmi'                  bmi''      htn_retrospective 
##                 412.17                 438.41               46071.57 
## diabetes_retrospective          smoking_prior 
##              152824.85                 137.51 
## 
## The following fit components were averaged over the 20 model fits:
## 
##   stats linear.predictors
```

```
ckd_1_aOR
```

```
##     Effect Lower 0.95 Upper 0.95 
##       2.96       1.89       4.64
```

```
ckd_2_aOR <- round(summary(fit.mult.impute(
  logistic_inpatient ~ ckd_retrospective + rcs(age, 4) + rcs(bmi, 4) +
    htn_retrospective + diabetes_retrospective + smoking_prior,
  lrm,
  imputation_outcomes,
  data = df_outcome %>% filter(excluded == "Included"))
)[2, c(4,6,7)], 2)
```

```
## 
## Variance Inflation Factors Due to Imputation:
## 
##              Intercept      ckd_retrospective                    age 
##                   1.40                   1.01                   1.01 
##                   age'                  age''                    bmi 
##                   1.00                   1.00                   1.52 
##                   bmi'                  bmi''      htn_retrospective 
##                   1.52                   1.51                   1.01 
## diabetes_retrospective          smoking_prior 
##                   1.02                   2.05 
## 
## Rate of Missing Information:
## 
##              Intercept      ckd_retrospective                    age 
##                   0.29                   0.01                   0.01 
##                   age'                  age''                    bmi 
##                   0.00                   0.00                   0.34 
##                   bmi'                  bmi''      htn_retrospective 
##                   0.34                   0.34                   0.01 
## diabetes_retrospective          smoking_prior 
##                   0.02                   0.51 
## 
## d.f. for t-distribution for Tests of Single Coefficients:
## 
##              Intercept      ckd_retrospective                    age 
##                 232.61              272029.35              363228.78 
##                   age'                  age''                    bmi 
##             1075640.48             1146044.82                 161.02 
##                   bmi'                  bmi''      htn_retrospective 
##                 162.83                 166.52              119682.68 
## diabetes_retrospective          smoking_prior 
##               50566.87                  72.23 
## 
## The following fit components were averaged over the 20 model fits:
## 
##   stats linear.predictors
```

```
ckd_2_aOR
```

```
##     Effect Lower 0.95 Upper 0.95 
##       2.89       1.75       4.77
```

```
ckd_3_aOR <- round(summary(fit.mult.impute(
  logistic_icu ~ ckd_retrospective + rcs(age, 4) + rcs(bmi, 4) +
    htn_retrospective + diabetes_retrospective + smoking_prior,
  lrm,
  imputation_outcomes,
  data = df_outcome %>% filter(excluded == "Included"))
)[2, c(4,6,7)], 2)
```

```
## 
## Variance Inflation Factors Due to Imputation:
## 
##              Intercept      ckd_retrospective                    age 
##                   1.13                   1.04                   1.00 
##                   age'                  age''                    bmi 
##                   1.01                   1.01                   1.29 
##                   bmi'                  bmi''      htn_retrospective 
##                   1.39                   1.43                   1.02 
## diabetes_retrospective          smoking_prior 
##                   1.04                   3.02 
## 
## Rate of Missing Information:
## 
##              Intercept      ckd_retrospective                    age 
##                   0.11                   0.03                   0.00 
##                   age'                  age''                    bmi 
##                   0.01                   0.01                   0.23 
##                   bmi'                  bmi''      htn_retrospective 
##                   0.28                   0.30                   0.02 
## diabetes_retrospective          smoking_prior 
##                   0.04                   0.67 
## 
## d.f. for t-distribution for Tests of Single Coefficients:
## 
##              Intercept      ckd_retrospective                    age 
##                1473.01               15522.00              931112.33 
##                   age'                  age''                    bmi 
##              332147.21              277044.18                 372.94 
##                   bmi'                  bmi''      htn_retrospective 
##                 239.79                 213.20               32570.02 
## diabetes_retrospective          smoking_prior 
##               14829.78                  42.49 
## 
## The following fit components were averaged over the 20 model fits:
## 
##   stats linear.predictors
```

```
ckd_3_aOR
```

```
##     Effect Lower 0.95 Upper 0.95 
##       1.65       0.64       4.23
```

#### 12.10 Risk factor: Chronic obstructive pulmonary disease

Defined as the presence of an ICD-10 diagnosis of J40 - J44 AND A prescription within the last year for drugs with ATC numbers R03 (Drugs for obstructive airway diseases).

```
copd_1_OR <- round(summary(fit.mult.impute(
  logistic_outpatient ~ copd_retrospective,
  lrm,
  imputation_outcomes,
  data = df_outcome %>% filter(excluded == "Included"))
)[2, c(4,6,7)], 2)
```

```
## 
## Variance Inflation Factors Due to Imputation:
## 
##          Intercept copd_retrospective 
##                  1                  1 
## 
## Rate of Missing Information:
## 
##          Intercept copd_retrospective 
##                  0                  0 
## 
## d.f. for t-distribution for Tests of Single Coefficients:
## 
##          Intercept copd_retrospective 
##       1.178556e+26       3.032992e+28 
## 
## The following fit components were averaged over the 20 model fits:
## 
##   stats linear.predictors
```

```
copd_1_OR
```

```
##     Effect Lower 0.95 Upper 0.95 
##       5.36       3.28       8.76
```

```
copd_2_OR <- round(summary(fit.mult.impute(
  logistic_inpatient ~ copd_retrospective,
  lrm,
  imputation_outcomes,
  data = df_outcome %>% filter(excluded == "Included"))
)[2, c(4,6,7)], 2)
```

```
## 
## Variance Inflation Factors Due to Imputation:
## 
##          Intercept copd_retrospective 
##                  1                  1 
## 
## Rate of Missing Information:
## 
##          Intercept copd_retrospective 
##                  0                  0 
## 
## d.f. for t-distribution for Tests of Single Coefficients:
## 
##          Intercept copd_retrospective 
##                Inf       2.151621e+29 
## 
## The following fit components were averaged over the 20 model fits:
## 
##   stats linear.predictors
```

```
copd_2_OR
```

```
##     Effect Lower 0.95 Upper 0.95 
##       6.87       3.90      12.11
```

```
copd_3_OR <- round(summary(fit.mult.impute(
  logistic_icu ~ copd_retrospective,
  lrm,
  imputation_outcomes,
  data = df_outcome %>% filter(excluded == "Included"))
)[2, c(4,6,7)], 2)
```

```
## 
## Variance Inflation Factors Due to Imputation:
## 
##          Intercept copd_retrospective 
##                  1                  1 
## 
## Rate of Missing Information:
## 
##          Intercept copd_retrospective 
##                  0                  0 
## 
## d.f. for t-distribution for Tests of Single Coefficients:
## 
##          Intercept copd_retrospective 
##       2.267996e+25       2.357321e+28 
## 
## The following fit components were averaged over the 20 model fits:
## 
##   stats linear.predictors
```

```
copd_3_OR
```

```
##     Effect Lower 0.95 Upper 0.95 
##      15.00       6.75      33.34
```

```
copd_1_aOR <- round(summary(fit.mult.impute(
  logistic_outpatient ~ copd_retrospective + rcs(age, 4) + smoking_prior,
  lrm,
  imputation_outcomes,
  data = df_outcome %>% filter(excluded == "Included"))
)[2, c(4,6,7)], 2)
```

```
## 
## Variance Inflation Factors Due to Imputation:
## 
##          Intercept copd_retrospective                age               age' 
##               1.00               1.01               1.00               1.00 
##              age''      smoking_prior 
##               1.00               1.52 
## 
## Rate of Missing Information:
## 
##          Intercept copd_retrospective                age               age' 
##               0.00               0.01               0.00               0.00 
##              age''      smoking_prior 
##               0.00               0.34 
## 
## d.f. for t-distribution for Tests of Single Coefficients:
## 
##          Intercept copd_retrospective                age               age' 
##       129908611.09          374246.31         4255872.44        20158564.22 
##              age''      smoking_prior 
##        18041731.42             160.73 
## 
## The following fit components were averaged over the 20 model fits:
## 
##   stats linear.predictors
```

```
copd_1_aOR
```

```
##     Effect Lower 0.95 Upper 0.95 
##       1.91       1.12       3.25
```

```
copd_2_aOR <- round(summary(fit.mult.impute(
  logistic_inpatient ~ copd_retrospective + rcs(age, 4) + smoking_prior,
  lrm,
  imputation_outcomes,
  data = df_outcome %>% filter(excluded == "Included"))
)[2, c(4,6,7)], 2)
```

```
## 
## Variance Inflation Factors Due to Imputation:
## 
##          Intercept copd_retrospective                age               age' 
##               1.00               1.03               1.00               1.00 
##              age''      smoking_prior 
##               1.00               2.02 
## 
## Rate of Missing Information:
## 
##          Intercept copd_retrospective                age               age' 
##               0.00               0.03               0.00               0.00 
##              age''      smoking_prior 
##               0.00               0.50 
## 
## d.f. for t-distribution for Tests of Single Coefficients:
## 
##          Intercept copd_retrospective                age               age' 
##       1.517373e+09       2.982273e+04       6.318020e+06       1.647967e+08 
##              age''      smoking_prior 
##       5.158141e+07       7.486000e+01 
## 
## The following fit components were averaged over the 20 model fits:
## 
##   stats linear.predictors
```

```
copd_2_aOR
```

```
##     Effect Lower 0.95 Upper 0.95 
##       1.56       0.81       3.01
```

```
copd_3_aOR <- round(summary(fit.mult.impute(
  logistic_icu ~ copd_retrospective + rcs(age, 4) + smoking_prior,
  lrm,
  imputation_outcomes,
  data = df_outcome %>% filter(excluded == "Included"))
)[2, c(4,6,7)], 2)
```

```
## 
## Variance Inflation Factors Due to Imputation:
## 
##          Intercept copd_retrospective                age               age' 
##               1.00               1.16               1.00               1.00 
##              age''      smoking_prior 
##               1.00               2.92 
## 
## Rate of Missing Information:
## 
##          Intercept copd_retrospective                age               age' 
##               0.00               0.14               0.00               0.00 
##              age''      smoking_prior 
##               0.00               0.66 
## 
## d.f. for t-distribution for Tests of Single Coefficients:
## 
##          Intercept copd_retrospective                age               age' 
##         6799451.75             989.94        10514211.90         1434419.50 
##              age''      smoking_prior 
##         1024439.37              43.90 
## 
## The following fit components were averaged over the 20 model fits:
## 
##   stats linear.predictors
```

```
copd_3_aOR
```

```
##     Effect Lower 0.95 Upper 0.95 
##       5.32       2.05      13.83
```

#### 12.11 Risk factor: History of cancer

```
cancer_1_OR <- round(summary(fit.mult.impute(
  logistic_outpatient ~ icd10_cancer,
  lrm,
  imputation_outcomes,
  data = df_outcome %>% filter(excluded == "Included"))
)[2, c(4,6,7)], 2)
```

```
## 
## Variance Inflation Factors Due to Imputation:
## 
##    Intercept icd10_cancer 
##            1            1 
## 
## Rate of Missing Information:
## 
##    Intercept icd10_cancer 
##            0            0 
## 
## d.f. for t-distribution for Tests of Single Coefficients:
## 
##    Intercept icd10_cancer 
## 3.193026e+25 3.746785e+29 
## 
## The following fit components were averaged over the 20 model fits:
## 
##   stats linear.predictors
```

```
cancer_1_OR
```

```
##     Effect Lower 0.95 Upper 0.95 
##       2.52       1.82       3.49
```

```
cancer_2_OR <- round(summary(fit.mult.impute(
  logistic_inpatient ~ icd10_cancer,
  lrm,
  imputation_outcomes,
  data = df_outcome %>% filter(excluded == "Included"))
)[2, c(4,6,7)], 2)
```

```
## 
## Variance Inflation Factors Due to Imputation:
## 
##    Intercept icd10_cancer 
##            1            1 
## 
## Rate of Missing Information:
## 
##    Intercept icd10_cancer 
##            0            0 
## 
## d.f. for t-distribution for Tests of Single Coefficients:
## 
##    Intercept icd10_cancer 
## 1.221662e+25 9.608446e+27 
## 
## The following fit components were averaged over the 20 model fits:
## 
##   stats linear.predictors
```

```
cancer_2_OR
```

```
##     Effect Lower 0.95 Upper 0.95 
##       3.62       2.40       5.47
```

```
cancer_3_OR <- round(summary(fit.mult.impute(
  logistic_icu ~ icd10_cancer,
  lrm,
  imputation_outcomes,
  data = df_outcome %>% filter(excluded == "Included"))
)[2, c(4,6,7)], 2)
```

```
## 
## Variance Inflation Factors Due to Imputation:
## 
##    Intercept icd10_cancer 
##            1            1 
## 
## Rate of Missing Information:
## 
##    Intercept icd10_cancer 
##            0            0 
## 
## d.f. for t-distribution for Tests of Single Coefficients:
## 
##    Intercept icd10_cancer 
##          Inf 7.321193e+29 
## 
## The following fit components were averaged over the 20 model fits:
## 
##   stats linear.predictors
```

```
cancer_3_OR
```

```
##     Effect Lower 0.95 Upper 0.95 
##       4.41       2.04       9.51
```

```
cancer_1_aOR <- round(summary(fit.mult.impute(
  logistic_outpatient ~ icd10_cancer + rcs(age, 4) + sex + smoking_prior,
  lrm,
  imputation_outcomes,
  data = df_outcome %>% filter(excluded == "Included"))
)[2, c(4,6,7)], 2)
```

```
## 
## Variance Inflation Factors Due to Imputation:
## 
##     Intercept  icd10_cancer           age          age'         age'' 
##          1.00          1.00          1.00          1.00          1.00 
##           sex smoking_prior 
##          1.01          1.55 
## 
## Rate of Missing Information:
## 
##     Intercept  icd10_cancer           age          age'         age'' 
##          0.00          0.00          0.00          0.00          0.00 
##           sex smoking_prior 
##          0.01          0.35 
## 
## d.f. for t-distribution for Tests of Single Coefficients:
## 
##     Intercept  icd10_cancer           age          age'         age'' 
##   38056155.63    4744114.21    2930233.65    8592038.90    7497200.94 
##           sex smoking_prior 
##     231702.55        152.63 
## 
## The following fit components were averaged over the 20 model fits:
## 
##   stats linear.predictors
```

```
cancer_1_aOR
```

```
##     Effect Lower 0.95 Upper 0.95 
##       0.99       0.68       1.42
```

```
cancer_2_aOR <- round(summary(fit.mult.impute(
  logistic_inpatient ~ icd10_cancer + rcs(age, 4) + sex + smoking_prior,
  lrm,
  imputation_outcomes,
  data = df_outcome %>% filter(excluded == "Included"))
)[2, c(4,6,7)], 2)
```

```
## 
## Variance Inflation Factors Due to Imputation:
## 
##     Intercept  icd10_cancer           age          age'         age'' 
##          1.00          1.00          1.00          1.00          1.00 
##           sex smoking_prior 
##          1.01          2.02 
## 
## Rate of Missing Information:
## 
##     Intercept  icd10_cancer           age          age'         age'' 
##          0.00          0.00          0.00          0.00          0.00 
##           sex smoking_prior 
##          0.01          0.50 
## 
## d.f. for t-distribution for Tests of Single Coefficients:
## 
##     Intercept  icd10_cancer           age          age'         age'' 
##  277371249.02    4164651.87    4843898.84  154984397.60   57191819.50 
##           sex smoking_prior 
##     113773.51         74.81 
## 
## The following fit components were averaged over the 20 model fits:
## 
##   stats linear.predictors
```

```
cancer_2_aOR
```

```
##     Effect Lower 0.95 Upper 0.95 
##       0.84       0.52       1.37
```

```
cancer_3_aOR <- round(summary(fit.mult.impute(
  logistic_icu ~ icd10_cancer + rcs(age, 4) + sex + smoking_prior,
  lrm,
  imputation_outcomes,
  data = df_outcome %>% filter(excluded == "Included"))
)[2, c(4,6,7)], 2)
```

```
## 
## Variance Inflation Factors Due to Imputation:
## 
##     Intercept  icd10_cancer           age          age'         age'' 
##          1.00          1.01          1.00          1.00          1.00 
##           sex smoking_prior 
##          1.03          2.85 
## 
## Rate of Missing Information:
## 
##     Intercept  icd10_cancer           age          age'         age'' 
##          0.00          0.01          0.00          0.00          0.00 
##           sex smoking_prior 
##          0.03          0.65 
## 
## d.f. for t-distribution for Tests of Single Coefficients:
## 
##     Intercept  icd10_cancer           age          age'         age'' 
##   30408779.14     616815.58   46714997.42    4278426.54    3040985.66 
##           sex smoking_prior 
##      28975.98         45.08 
## 
## The following fit components were averaged over the 20 model fits:
## 
##   stats linear.predictors
```

```
cancer_3_aOR
```

```
##     Effect Lower 0.95 Upper 0.95 
##       1.25       0.54       2.86
```

### 13 Performance metrics

```
df_perforrmance <- df_val_validation%>%
  mutate(
    logistic_outpatient = factor(outpatient_fct),
    logistic_inpatient = factor(inpatient_fct),
    logistic_icu_death = factor(icu_death_fct)
  ) %>%
  select(logistic_outpatient,
         logistic_inpatient,
         logistic_icu_death,
         `y>=1`,
         `y>=2`,
         `y>=3`)

df_threshold <- threshperf(
  df = df_perforrmance,
  outcome = "logistic_outpatient",
  prediction = "y>=1"
) %>%
  mutate(outcome = "Outpatient or worse") %>%
  union_all(
    threshperf(
      df = df_perforrmance,
      outcome = "logistic_inpatient",
      prediction = "y>=2"
    ) %>%
      mutate(outcome = "Hospital admission or worse")
  ) %>%
  union_all(
    threshperf(
      df = df_perforrmance,
      outcome = "logistic_icu_death",
      prediction = "y>=3"
    ) %>%
      mutate(outcome = "ICU admission or death")
  )

figure_performance <- df_threshold %>%
  mutate(
    .metric = fct_relevel(case_when(
      .metric == "sens" ~ "Sensitivity",
      .metric == "spec" ~ "Specificity",
      .metric == "ppv" ~ "PPV",
      .metric == "npv" ~ "NPV"
    ), "Sensitivity", "Specificity", "PPV", "NPV"),
    outcome = fct_relevel(
      outcome,
      "Outpatient or worse",
      "Hospital admission or worse",
      "ICU admission or death"
    )
  ) %>%
  ggplot(aes(
    x = .threshold,
    y = PointEst,
    ymax = ul,
    ymin = ll,
    color = outcome,
    fill = outcome
  )) +
  geom_line() + geom_ribbon(aes(color = NULL), alpha = 0.5) +
  facet_wrap(~.metric, ncol = 1, strip.position = "right") +
  scale_y_continuous(label = scales::percent) +
  scale_color_manual(values = nejm_palette[c(1, 2, 4)]) +
  scale_fill_manual(values = nejm_palette[c(1, 2, 4)]) +
  labs(x = "Threshold", y = "Performance (%)") +
  theme_bw() +
  theme(
    legend.position = c(0.8, 0.9),
    legend.title = element_blank(),
    legend.text = element_text(size = 11),
    legend.key = element_rect(fill = "transparent", color = "transparent"),
    legend.background = element_rect(fill = "transparent", color = "transparent")
  )

figure_performance
```

### 14 Case study of model

```
df_n <- df_phone %>%
  right_join(
    df_validation %>%
      filter(excluded == "Included") %>%
      select(kt)
  ) %>%
  select(kt, call_nr, date) %>%
  distinct()

df_benefit <- df_validation %>%
  filter(excluded == "Included") %>%
  left_join(
    df_n %>%
      count(kt, name = "obs_n")
  ) %>%
  select(kt, outcome_fct, obs_n) %>%
  bind_cols(
    tbl_df(
      predict(
        mod_multi_derivation,
        df_validation,
        type = "fitted")
    )) %>%
  select(kt, outcome_fct, obs_n, prob = `y>=1`)

sequence_benefits <- seq(from = 0, to = 1, by = 0.001)
output_benefits <- matrix(ncol = 7, nrow = length(sequence_benefits))

for(i in 1:length(sequence_benefits)){
  threshold <- sequence_benefits[i]
  total_observed <- sum(df_benefit$obs_n)

  total_pred <- df_benefit %>%
    mutate(pred_n = case_when(
      prob <= threshold ~ 2L,
      TRUE ~ obs_n
    )) %>%
    pull(pred_n) %>%
    sum()

  nr_prevented_phonecalls <- total_observed - total_pred

  outpatient_nr_missed <- df_benefit %>%
    filter(prob <= threshold & outcome_fct == 1) %>%
    nrow()

  inpatient_nr_missed <- df_benefit %>%
    filter(prob <= threshold & outcome_fct == 2) %>%
    nrow()

  critical_nr_missed <- df_benefit %>%
    filter(prob <= threshold & outcome_fct == 3) %>%
    nrow()

  output_benefits[i, 1] <- threshold
  output_benefits[i, 2] <- total_observed
  output_benefits[i, 3] <- total_pred
  output_benefits[i, 4] <- nr_prevented_phonecalls
  output_benefits[i, 5] <- outpatient_nr_missed
  output_benefits[i, 6] <- inpatient_nr_missed
  output_benefits[i, 7] <- critical_nr_missed
}

df_output_benefits <- tbl_df(as.data.frame(output_benefits)) %>%
  mutate(V1 = 1-V1)

names(df_output_benefits) <- c("threshold", "total_observed", "total_pred", "total_prevented",
                               "missed_outpatient", "missed_inpatient", "missed_critical")

plot_prevented <-  df_output_benefits %>%
  mutate(
    threshold = rev(threshold),
    percent_prevented = total_prevented/total_observed
  ) %>%
  filter(threshold <= 0.12) %>%
  ggplot(aes(x = threshold, y = percent_prevented)) +
  geom_line(size = 0.7) +
  geom_vline(xintercept = 0.041, lty = 2) +
  scale_y_continuous(
    limits = c(NA, 0.48),
    labels = scales::percent_format(accuracy = 1L)
  ) +
  scale_x_continuous(
    breaks = seq(0, 0.12, 0.01),
    limits = c(0, 0.12),
    labels = scales::percent_format(accuracy = 1L)
  ) +
  labs(x = NULL, y = "Prevented \ntelehealth interviews") +
  theme_bw()

plot_prevented
```

```
plot_missed_cases <- df_output_benefits %>%
  mutate(
    threshold = rev(threshold)) %>%
  filter(threshold <= 0.12) %>%
  pivot_longer(
    cols = starts_with("missed"),
    names_to = "type",
    values_to = "n"
  ) %>%
  mutate(
    type = fct_relevel(
      type,
      "missed_outpatient",
      "missed_inpatient",
      "missed_critical"
    )) %>%
  left_join(
    tibble(
      type = as_factor(c(
        "missed_outpatient",
        "missed_inpatient",
        "missed_critical"
      )),
      total_case = c(
        df_validation %>% filter(outcome_fct == "1") %>% nrow(),
        df_validation %>% filter(outcome_fct == "2") %>% nrow(),
        df_validation %>% filter(outcome_fct == "3") %>% nrow()
      )
    )
  ) %>%
  mutate(percent_prevented = n/total_case) %>%
  ggplot(aes(x = threshold, y = percent_prevented, color = type)) +
  geom_line(size = 0.7, position = position_dodge(width = 0.0001)) +
  geom_vline(xintercept = 0.041, lty = 2) +
  scale_y_continuous(
    limits = c(NA, 0.48),
    labels = scales::percent_format(accuracy = 1L)
  ) +
  scale_color_manual(
    name = element_blank(),
    values = nejm_palette[c(1, 2, 4)],
    labels = c("Outpatient", "Hospital admission", "Intensive care or death")
  ) +
  scale_x_continuous(
    breaks = seq(0, 0.12, 0.01),
    limits = c(0, 0.12),
    labels = scales::percent_format(accuracy = 1L)
  ) +
  labs(x = NULL, y = "Cases not in \nactive monitoring") +
  theme_bw() +
  theme(
    legend.position = c(0.18, 0.6),
    legend.text = element_text(size = 12),
    legend.key = element_rect(fill = "transparent", color = "transparent"),
    legend.background = element_rect(fill = "transparent", color = "transparent")
  )

plot_missed_cases
```

```
plot_ratio <- df_output_benefits %>%
  mutate(threshold = rev(threshold)) %>%
  filter(threshold <= 0.12) %>%
  pivot_longer(
    cols = starts_with("missed"),
    names_to = "type",
    values_to = "n"
  ) %>%
  mutate(
    type = fct_relevel(type, "missed_outpatient", "missed_inpatient", "missed_critical"),
    ratio = total_prevented/n,
    ratio = if_else(ratio %in% c(NaN, Inf, -Inf), total_prevented, ratio)
  ) %>%
  filter(ratio > 0) %>%
  ggplot(aes(x = threshold, y = ratio, color = type)) +
  geom_line(size = 0.7, position = position_dodge(width = 0.0005)) +
  geom_vline(xintercept = 0.041, lty = 2) +
  scale_y_log10(
    breaks = c(20, 50, 200, 500, 1000, 4000, 7000),
    minor_break = NULL,
    limits = c(20, 7000),
    labels = scales::comma
  ) +
  annotation_logticks(side = "l") +
  scale_color_manual(
    name = element_blank(),
    values = nejm_palette[c(1, 2, 4)],
    labels = c("Outpatient", "Hospital admission", "Intensive care or death")
  ) +
  scale_x_continuous(
    breaks = seq(0, 0.12, 0.01),
    limits = c(0, 0.12),
    labels = scales::percent_format(accuracy = 1L)
  ) +
  labs(
    x = "Threshold of predicted probability for active monitoring",
    y = "Prevented interviews per case \nnot in active monitoring") +
  theme_bw() +
  theme(legend.position = "none")

plot_ratio
```

```
figure_combined_example <- plot_grid(
  plot_prevented,
  plot_missed_cases,
  plot_ratio,
  align = "hv",
  ncol = 1
)

figure_combined_example
```
